# Mainly Heterosexual, Bisexual, or Other?: the measurement of sexual minority status and its impact on analytic sample, demographic distribution and health outcomes

**DOI:** 10.1101/2023.03.06.23286850

**Authors:** Evangeline Tabor, Dylan Kneale, Praveetha Patalay

**Affiliations:** Social Research Institute, University College London, London; MRC Unit for Lifelong Health and Ageing, University College London, London

**Author notes:** **Corresponding author:** Tabor, Evangeline.

## Abstract

**Background:** Sexual orientation has been measured in a wide variety of ways which reflect both theoretical and practical considerations. However, choice of sexual orientation measure and recoding strategy can impact analytic sample, as well as demographic and health profiles, in analyses of sexual minority populations. We aimed to examine how choice of sexual orientation dimension and recoding decisions impact estimates in the sexual minority population in two population-based studies in the UK.

**Methods:** We used data collected at age 17 (2018) in the UK Millennium Cohort Study and at wave six (2012-13) and eight (2017-18) of the English Longitudinal Study of Ageing. Descriptive statistics were used to examine the impact of choice of sexual orientation dimension (i.e. identity, attraction and experience) and recoding decisions on achieved analytic sample and composition by selected demographic and health measures within and between datasets.

**Results:** Dimension choice and recoding decisions resulted in variation in analytic sample. For example, more respondents reported some same-sex sexual attraction than reported a non-heterosexual identity (adolescents: 20.77% vs 8.97%, older adults: 4.77% vs 1.04%). Demographic distributions varied, but not substantially by dimension choice or recoding strategy. Overall, in both datasets sexual minority respondents were more likely to be White and in the highest quintiles for income and education than heterosexual respondents. Health status did not vary substantially by dimension choice or recoding strategy, however sexual minority respondents reported worse health than their heterosexual peers.

**Conclusions:** This study explores a range of practical and theoretical considerations when analysing sexual minority respondents using survey data. We highlight the impact recoding decisions may have on the numbers of sexual minority respondents identified within a dataset and demographic and health distributions in this understudied population. We also demonstrate the benefits of including multiple dimensions for capturing mechanisms of interest in elucidating ambiguous responses and exploring sexual diversity.

## Background

Recent research using large observational datasets has shown sexual minority people in the UK have poorer mental and physical health outcomes than heterosexual people (1–3). This research has been facilitated by the growth in inclusion of relevant measures to population health and demographic datasets, exemplified by the recent introduction of routine collection of sexual identity by the NHS and the inclusion of a sexual identity question in the 2021 England and Wales Census (4,5). However, the dimension of sexuality captured by the included measures and therefore used by researchers varies. For example, studies have used respondent sexual identity, sexual attraction, and sexual behaviour (6), each collected through different questions and allowing for different response options. These different measures represent the result of a range of theoretical and practical considerations, including survey design, queer and sexuality theory, and public policy priorities (see Sell, 2007; Savin-Williams, 2009; Wolff *et al.*, 2017; Matsuno *et al.*, 2020, Russell *et al*., 2023 for more information), but have important consequences for research into inequities by sexual minority status. For example, health outcomes may vary by which dimension of sexual orientation is selected for measurement, and focusing on only one dimension may miss important health disparities and limit the effectiveness of policy recommendations (6). Similarly, researchers are encouraged to use the dimension of sexual orientation which is most relevant to their research question, however this is constrained by which dimensions have been captured (10).

In the following sections we will examine four ways sexual orientation has been measured and analysed: sexual attraction, sexual behaviour, sexual identity, and through composite measures. We provide a brief overview of theoretical and practical considerations for each, as well as limitations of their operationalisation in quantitative survey research. Finally, we will discuss how researcher choices may further influence how sexual minority populations are analysed.

### Attraction and Behaviour

Sexual attraction and behaviour have been used to identify sexual minority individuals who may not all identify with the LGBTQ+ community or a specific identity (9). For some researchers, attraction is the most important dimension of sexual orientation as it can be considered the primary source of other dimensions such as behaviour and identity (7,8). Some have argued it is also the most consistent dimension across time and context, while others have argued it may be stable over many years but fluid across a lifetime (8,11,12). Likewise, measuring sexual behaviour has been an important tool for monitoring sexual health (13). For example, terms in the public health literature such as ‘men who have sex with men’ (MSM) have been used to identify individuals who may be at risk of sexual transmitted infections such as HIV and as a way of avoiding the social and cultural implications of identity labels (14).

However, measuring attraction and behaviour has several challenges. Firstly, while there is considerable overlap between reporting same-sex attraction and behaviour and sexual identity, those who report any same-sex attraction or behaviour are a much larger proportion of the population (13). Many of those who report same-sex attraction or behaviour do not report an LGB+ identity, and reported attraction and behaviour do not overlap neatly (13). Secondly, the timescale on which the behaviour and attraction is measured has important implications for inferring sexual orientation, and varies widely between measures (6,13). For example, measuring lifetime prevalence of same-sex behaviour may reflect fluidity over time and experimentation during development, while recent behaviour may map more onto current sexual identity (6,13). Additionally, questions on sexual behaviour may be perceived as too invasive or sensitive by respondents and irrelevant or inappropriate to ask of adolescents by their guardians which may result in survey attrition or low response rates (13). Finally, what is meant by sexual attraction and behaviour are often left underdefined in questions with variation in understanding of the term between individuals (6,8,15). Likewise, sexual behaviour and attraction measures are often predicated on the existence of binary genders or sexes which may not match respondents own understanding (6). Overall, these measures have an advantage in identifying those who do not align themselves with the LGBTQ+ community and therefore may not be reached by existing support systems. However, alone neither can tell researchers much about each other, or the identity and experiences of respondents.

### Sexual identity

Sexual identity has been repeatedly recommended as a key measure for population health surveys (6,16,17). Sexual identity is not just a proxy marker for sexual attraction and behaviour, but also a powerful expression of self-identity, community, and political alignment (14,16,18). Naming is a way of exercising power and control, and for marginalised groups the right to determine their own name is often hard won (14). Terms such as ‘gay’, ‘lesbian’, and ‘LGBT’ form part of the language of global political movements for rights and freedoms, as well as a global LGBTQ+ community, in ways that public health terms such as ‘men who have sex with men’ do not (14,18). Sexual identities can invoke a wider conversation about discrimination and disadvantage in a way that measures of attraction and behaviour may obscure (16).

However, as with attraction and behaviour, measuring sexual identity has a number of limitations and considerations. Firstly, identity categories are historically and culturally specific (14,18). While terms like gay and bisexual have global salience in rights and equalities landscapes, there are many culturally, temporally, or culturally specific terms which may not map neatly onto these more dominant terms (14). As a result, options presented in quantitative research may capture only the closest approximation of a response or fail to be comprehensible to respondents at all. As such, important nuance and distinctions may be lost, and those who don’t consider themselves as sitting within such terms may be obscured (14). Given that those most likely to give responses outside of common terms such as heterosexual, gay/lesbian and bisexual are also generally older, of lower socioeconomic status and more likely to belong to an ethnic minority group, this may only exacerbate existing inequalities (19). As we can see from this example, sexual identity intersects with other identities such as ethnicity, age, and gender in ways which both materially impact an individual’s experience of the world and their response to measures of sexual identity (6,13,20,21). Secondly, as with behaviour and attraction, it is unclear how stable identity is across time (22,23). For example, in Diamond’s studies of sexual identity, nearly two-thirds of young women in the study changed their identity label at least once in a ten year period (8,23). Similarly, Hu & Denier (2023)’s recent study using a UK longitudinal study found that 6.6% of their respondents changed their identity label over a six year timeframe (24). Heterosexual and gay/lesbian respondents were largely stable while those who reported their identity as “Other” or did not disclose had the highest mobility between time points (24). As a result, researchers should be mindful of the temporal and social variability of sexual identity.

### Composite methods

As discussed above, there is often a false assumption of interchangeability or perfect alignment between the different dimensions of sexuality (6,14). For example, a lesbian identified woman is assumed to only experience attraction to and engage in sexual behaviours with women, and consider themselves part of a community of lesbian-identified women (6). However, sexuality can be better understood as a matrix which can be fluid, dynamic and multifaceted (6,14).

While the incorporation of more complex models of sexual orientation are theoretically grounded, translating these models into quantitative measures and empirical analysis has been limited (6). There have been many attempts to incorporate multiple dimensions of sexuality or develop complex measures however they have not found widespread use or acceptance, not been properly validated, or have been cumbersome to use empirically (6,7). As a result of these issues, and pressure for single-question measures due to limited time and resources in large quantitative studies, researchers are largely restricted to relying on measures of single dimension (6,7). However, some studies capture more than one dimension allowing for comparison between dimensions and the potential to differentiate the impact of identity, attraction, and behaviour on outcomes of interest. In addition, the collection of measures at multiple time points allows for greater understanding of the development of sexuality over the lifecourse, changes across age, cohort and life events, and, in longitudinal studies, reduces the likelihood of missing data (23,24).

### Sexual orientation data

As of the 2021 England and Wales Census, 3.2% of the England and Wales population identified as gay, lesbian, bisexual or “other sexual orientation” (25). As a result, even in representative population surveys, sample size remains a persistent issue in analyses of sexual minorities. One common solution is recoding - a data management practice of re-categorising variable responses in new ways, often to combine and reduce possible responses, before data analysis is undertaken. While it is often a necessary practical step to ensure statistical analysis can take place, it can obscure differences within the sexual minority population, including significant differences in health outcomes (6,26). The asymmetric nature of heterosexual and sexual minority population sizes means that choices about who is or isn’t included becomes particularly meaningful, yet the inclusion or exclusion of certain responses is often left unjustified by researchers.

Savin-Williams (2009) argues these decisions raises the question of ‘how much’ of a given dimension is needed for inclusion as sexual minority, especially as it interacts with methodological concerns around sample size and representativeness (8). For example, sexual attraction or behaviour measures do not require association with a social identity nor repeated experience, and as a result have relatively low ‘barrier to entry’ to reporting. As a result, who should be included in a ‘sexual minority’ category using attraction and experience data is not immediately obvious. However, generating a cut-off for ‘significant’ same-sex attraction or experience creates arbitrary categories of those who are considered to have ‘enough’ same-sex attraction or experience to be considered sexual minority. The question of ‘enough’ has been used to police queer individuals’ participation in the LGBTQ+ community, erase or invalidate identities such as bisexuality, and been used to deny refugee status to LGBTQ+ asylum seekers (27,28). As a result, the decisions of researchers post data collection may influence the outcomes of interest as much as the measures of sexuality themselves. However, there is little in the literature discussing this important but often obscured element of quantitative research on sexual minority communities.

### The current analysis

Measurement of sexual orientation is not straightforward and, while there is concordance, dimensions of sexuality are not interchangeable. Much of the work of preparing the responses to measures of sexuality are rarely disclosed in detail despite the impact measures and recoding may have on the demographic makeup of the analytic sample and outcomes such as health and wellbeing.

This analysis critically examines the implications of using three different measures, and two illustrative examples of coding response patterns, using data from two population-based cohort studies from the UK which have captured multiple dimensions of sexuality and represent populations of particular interest to researchers (i.e. adolescents and older adults). We aim to illustrate the impact of dimension choice and researcher coding decisions on analytic sample and demographic and health distributions. We hope this analysis will serve as a resource for researchers who work with these studies or plan to use or collect sexual minority respondent data in the future.

## Methods

### Study Samples

The study incorporates data from two longitudinal population-based studies: the Millennium Cohort Study (MCS (adolescents)) and the English Longitudinal Study of Ageing (ELSA (older adults)). These datasets were selected as they measured multiple dimensions of sexuality, collected information on both demographic distributions and health outcomes, and were sampled using probabilistic and representative methods. The two datasets also allow for the comparison of two populations representing distinct age groups and generations.

MCS is a large longitudinal birth cohort study of children born between September 2000 and January 2002 (29). There have been seven data collection sweeps at ages 9 months, 3, 5, 7, 11, 14, and 17 years to date (29). Children living in disadvantaged areas, and children from ethnic minority backgrounds were deliberately oversampled to allow analysis of these populations (29). Further detail is available elsewhere (29,30).

ELSA is a longitudinal panel study of people aged 50 and over in England (31). The original sample of respondents was selected from households who had previously responded to the Health Surveys for England, a cross-sectional nationally representative household survey, in 1998, 1999 or 2001, with the sample replenished periodically thereafter (30–32).

### Variables

Sexuality is measured in the datasets in three main ways: sexual identity, same-sex attraction, and same-sex behaviour. The measures of sexuality captured in MCS and ELSA are described in Table 1. The following demographic variables were used to describe the composition of samples: sex and gender identity, age, income quintile, education to degree level, ethnicity, and urbanity. Health outcome variables included are life satisfaction, psychological distress and self-rated general health. Each measure is described below.

**Table 1:** Sexuality variables in adolescents and older adults (% excluding missing)

|  | Adolescents | Older adults |  |
| --- | --- | --- | --- |
| **N=** | 10,103 | 7,130 |  |
| **Study & Collection sweep** | MCS (2018) | ELSA (2012-13) | ELSA (2017-18) |
| **Identity** |  |  |  |
| Which of the following options best describes how you currently think of yourself? N (%) |  |  |  |
| Completely Heterosexual | 7888 (76.3) | - | - |
| Mainly Heterosexual | 1101 (10.6) | - | - |
| Bisexual | 656 (6.3) | - | - |
| Mainly Gay or Lesbian | 90 (0.9) | - | - |
| Completely Gay or Lesbian | 160 (1.6) | - | - |
| Other | 157 (1.5) | - | - |
| Don't Know | 16 (0.2) | - | - |
| PNS | 35 (0.3) | - | - |
| Missing | - | - | - |
| Which of the following options best describes how you think of yourself? N (%) |  |  |  |
| Heterosexual | - | - | 4,513 (94.7) |
| Gay or Lesbian | - | - | 31 (0.7) |
| Bisexual | - | - | 43 (0.9) |
| Other | - | - | 27 (0.6) |
| PNS | - | - | 152 (3.2) |
| Missing | - | - | 2,364 |
| **Attraction** |  |  |  |
| I have felt sexually attracted... (Adolescents) N (%) |  |  |  |
| Which statement best describes your sexual desires over your lifetime? Please include being interested in sex, fantasising about sex or wanting to have sex? (Older adults) N (%) |  |  |  |
| Only to opposite | 7656 (75.8) | 5,666 (93.0) | - |
| Mostly to opposite | 1382 (13.7) | 218 (3.6) | - |
| Equally | 385 (3.8) | 57 (0.9) | - |
| Mostly to same | 194 (1.9) | 19 (0.3) | - |
| Only to same | 137 (1.4) | 46(0.8) | - |
| No attraction/desires | 291 (2.9) | 87 (1.4) | - |
| Don't Know | 11 (0.1) | - | - |
| PNS | 46 (0.5) | - | - |
| Missing | 1 | 1,037 | - |
| **Behaviour** |  |  |  |
| Finally, we would like to know a little about your lifetime sexual experiences and desires. Which statement best describes your sexual experiences over your lifetime? Please include all sexual experiences including sexual intercourse, fondling and petting.N (%) |  |  |  |
| Only w. opposite | - | 5,790 (94.8) | - |
| Mostly w. opposite | - | 158 (2.6) | - |
| Equally | - | 23 (0.4) | - |
| Mostly w. same | - | 27(0.4) | - |
| Only w. same | - | 33 (0.5) | - |
| No experience | - | 79 (1.3) | - |
| Don't Know | - | - | - |
| PNS | - | - | - |
| Missing | - | 1,020 | - |

#### Sex and Gender Identity

Gender identity in MCS is collected at age 17 with following question: “Which of the following describes how you think of yourself?”. The response options are: “Male”, “Female”, “In another way”, and “Prefer not to say”. Those who selected “In another way” were asked to write their response in a text box. In this analysis, those who were categorized as “Other”, “Androgynous / male and female”, “Gender fluid”, “Non-binary” respondents were recoded as “Non-Binary/Other”. Those who were categorized as “Vague irrelevant answer” were recoded as missing, and those who responded “Don’t know” or “Prefer Not to Say” were recoded as “Don’t Know/PNS”.

A further sex variable was included for MCS which was derived from information collected at birth. Options are “Male” or “Female”.

In ELSA, no gender identity variable was available so sex was used for this analysis. Respondents are asked for their sex on entering the study. Options are “Male” or “Female”. This answer is confirmed at each wave.

#### Age

As MCS is a birth cohort study, there is little to no difference in age at participation at each wave, so all participants were aged between 16-18 at the age 17 sweep, with the majority assessed at age 17. However, for ELSA age varies between participants at each wave. For ELSA, respondents will be grouped into the following intervals: “50-59”, “60-69”, “70-79”, “80-89”, and “90+”. A categorical variable rather than a continuous variable is included in this analysis as those over the age of 90 are collapsed in the dataset. For ELSA, age at sexual identity collection was used (wave 8 (2016-17)).

#### Socio-economic status

In both datasets quintiles of household income with OECD equivalence scaling and an eight-category education variable were used to indicate socioeconomic status. In addition, in ELSA, quintiles of net household wealth were also used. All variables used were collected at wave 8 (2016-17) in ELSA, and at age 14 for MCS (2015).

#### Ethnicity

For both datasets ethnicity will be presented as a binary “White” and “Non-White”.

#### Government Office Region and Urbanity

ELSA includes a Government Office Region variable. Government Office Region is a twelve-item category: “North East”, “North West”, “Yorkshire and the Humber”, “East Midlands”, “West Midlands”, “East of England”, “London”, “South East”, “South West”, “Wales”, “Scotland”, and “Northern Ireland”. In MCS a binary urban/rural variable was used using 2005 ONS Rural/Urban Classification.

### Health and wellbeing measures

A measure of life satisfaction, general mental health, and general physical health was selected for each dataset based on their suitability for cross-cohort comparison, and relation to health outcomes of interest.

#### Life satisfaction

The following self-reported life satisfaction measures were used:

MCS at age 17 (2018): “How much do you agree or disagree with the following statements about you?: On the whole, I am satisfied with myself”. Options include: “Strongly agree”, “Agree”, “Disagree”, “Strongly disagree”. Respondents could also answer “Do not know” and “I do not wish to answer”.

ELSA at wave 8 (2016-17): “Please say how much you agree or disagree with the following statements: I am satisfied with my life”. Options include: “Strongly agree”, “Agree”, “Slightly Agree”, Neither agree nor disagree”, “Slightly disagree”, “Disagree”, and “Strongly disagree”.

#### Mental Health

The K-6 is a measure of psychological distress measured in MCS at age 17 (2018). K-6 is a validated six-item measure of psychological distress. Referring to the last 30 days, respondents are asked to respond to items with five response options (ranging from none of the time to all of the time) (33). Scores ranged from 0 to 24 with higher scores indicating greater distress. The scale has moderate and severe (≥13) thresholds which are indicative of moderate and serious mental illness (33,34). K-6 questions include: “During the last 30 days, about how often did you feel so depressed that nothing could cheer you up?” and “During the last 30 days, about how often did you feel hopeless?”.

The Center for Epidemiological Studies Depression Scale or CES-D is a validated eight-item measure of depression (35,36). Referring to the past week, respondents are asked to respond to items with a binary “Yes” or “No”. Scores range from 0 (no symptoms) to 8 (all symptoms) with a score of 3 or more indicative of depression ‘caseness’ (36). CES-D questions include: “Much of the time during the past week, you felt depressed?” and “Much of the time during the past week, you felt lonely?”. Responses from ELSA wave 8 (2016-17) are used in this analysis.

#### Physical Health

The following self-rated general health measures were used:

MCS at age 17: “How would you describe your health generally? Would you say it is…”. Options included: “Excellent”, “Very Good”, “Good”, “Fair”, “Poor”, “Don’t know”, and “I do not wish to answer”.

ELSA at wave 8 (2016-17): “Would you say your health is…”. Options included: “Excellent”, “Very Good”, “Good”, “Fair”, and “Poor”.

### Analysis

This study is composed of three descriptive analyses:

#### Analysis 1: Describe the dimensions of sexuality captured and investigate how they overlap in young people and older adults

We generated descriptive statistics of the distributions and missingness of sexual identity, attraction, and behaviour in each dataset. Using a series of cross-tabulations, we then generated descriptive statistics of the overlap between these dimensions and presented as Venn diagrams.

#### Analysis 2: Describe how choice of sexuality dimension impacts demographic distributions (i.e. sex/gender, SES, and ethnicity), and health outcomes

We present descriptive statistics of sex/gender, age, income, education, ethnicity, and urbanity to examine variation in reporting by different dimensions of sexuality. We also present descriptive statistics of life satisfaction, self-rated general health, and a measure of psychological distress to examine variation in reporting by different dimensions of sexuality.

#### Analysis 3: Describe how recoding sexuality dimensions for analysis impacts analytic sample, demographic distributions (i.e. sex/gender, SES, and ethnicity), and health outcomes

We present descriptive statistics of the demographic and health variables as above. However, in this analysis we present variation resulting from researcher choice regarding sexuality dimension recoding. In this paper we present two illustrative examples which sit along a continuum of potential approaches and are described in Box 2.

##### Box 2.

Box 2:
| Approach | Broad | Restricted |
| --- | --- | --- |
| **Description** | Approach which prioritizes an expansive definition of sexual minority. For example, including those who report their identity as Other and those who do not know or do not want to say. Likewise, it may encompass those who report no attraction or heterosexuals who report some same sex attraction. | Approach which prioritises respondents who sit clearly within the sexual minority category and those with coincident identity and attraction/experience. For example, those who identify as lesbian, gay, and bisexual, have “significant” same-sex attraction, and whose identity is coincident with reported attraction etc. |
| **Pros** | - Inclusive of the diversity of the LGBTQ+ and sexual minority population.<br>- Increases potential analytic sample | - Conceptually clearly defined.<br>- A clear match between decision and population of interest |
| **Cons** | - May be difficult to know exactly what mechanism is under investigation.<br>- May not reflect common understandings of LGBTQ+ or sexual minority populations. | - May be overly restrictive and not reflect real-world diversity.<br>- Restricts potential analytic sample numbers. |
| **Strategy in Coding Identity Data** | Respondents who reported their sexual identity as “Heterosexual” or as “Completely Heterosexual” are recoded as “Non-Sexual Minority”.<br>Respondents who reported their sexual identity as “Lesbian or Gay”, “Bisexual”, “Other”, “Prefer Not To Say” and “Don’t Know” or as “Completely Lesbian/Gay”, “Mainly Gay/Lesbian”, and “Mainly Heterosexual” are recoded as “Sexual Minority”. | Respondents who reported their sexual identity as “Heterosexual”, “Completely Heterosexual” or “Mainly Heterosexual” are recoded as “Non-Sexual Minority”.<br>Respondents who reported their sexual identity as “Lesbian or Gay” and “Bisexual” or as “Completely Lesbian/Gay” and “Mainly Gay/Lesbian” are recoded as “Sexual Minority”. |
| **Strategy in Coding Attraction/ Behaviour Data** | Respondents who reported their attraction or experience as “Only opposite” are recoded as “Non-Sexual Minority”.<br>Respondents who reported their attraction or experience as “Only same”, “Mostly same”, “Equally”, “Mostly Opposite”, “No attraction/experience”, “Prefer Not To Say” and “Don’t Know” are recoded as “Sexual Minority”. | Respondents who reported their attraction or experience as “Only opposite” or “Mostly Opposite” are recoded as “Non-Sexual Minority”.<br>Respondents who reported their attraction or experience as “Only same”, “Mostly same” and “Equally” are recoded as “Sexual Minority”. |
| **Strategy in Coding Data where multiple measures are available** | Respondents who reported their sexual identity as “Heterosexual” or as “Completely Heterosexual” AND reported their attraction and experience as “Only opposite” are recoded as “Non-Sexual Minority”.<br>Respondents who reported their sexual identity “Lesbian or Gay”, “Bisexual”, “Other”, “Prefer Not To Say and Don’t Know or as “Completely Lesbian/Gay” and “Mainly Gay/Lesbian” OR reported their attraction and experience as “Only same”, “Mostly same”, “Equally”, “Mostly Opposite” and “No attraction/experience” are recoded as “Sexual Minority”. | Respondents who reported their sexual identity as “Heterosexual”, “Completely Heterosexual” or “Mainly Heterosexual” OR reported their attraction or experience as “Only opposite” or “Mostly Opposite” are recoded as “Non-Sexual Minority”.<br>Respondents who reported their sexual identity as “Lesbian or Gay” and “Bisexual” or as “Completely Lesbian/Gay” and “Mainly Gay/Lesbian” AND reported their attraction and experience as “Only same”, “Mostly same” and “Equally” are recoded as “Sexual Minority”. |

## Results

This analysis includes study members who reported their sexual identity, attraction and/or behaviour in at least one wave. The study sample is 10,103 adolescents (MCS) and 7,130 older adults (ELSA).

### Non-response and analytic sample

The derivation of the analytic sample in each dataset is described in the flow diagram below:

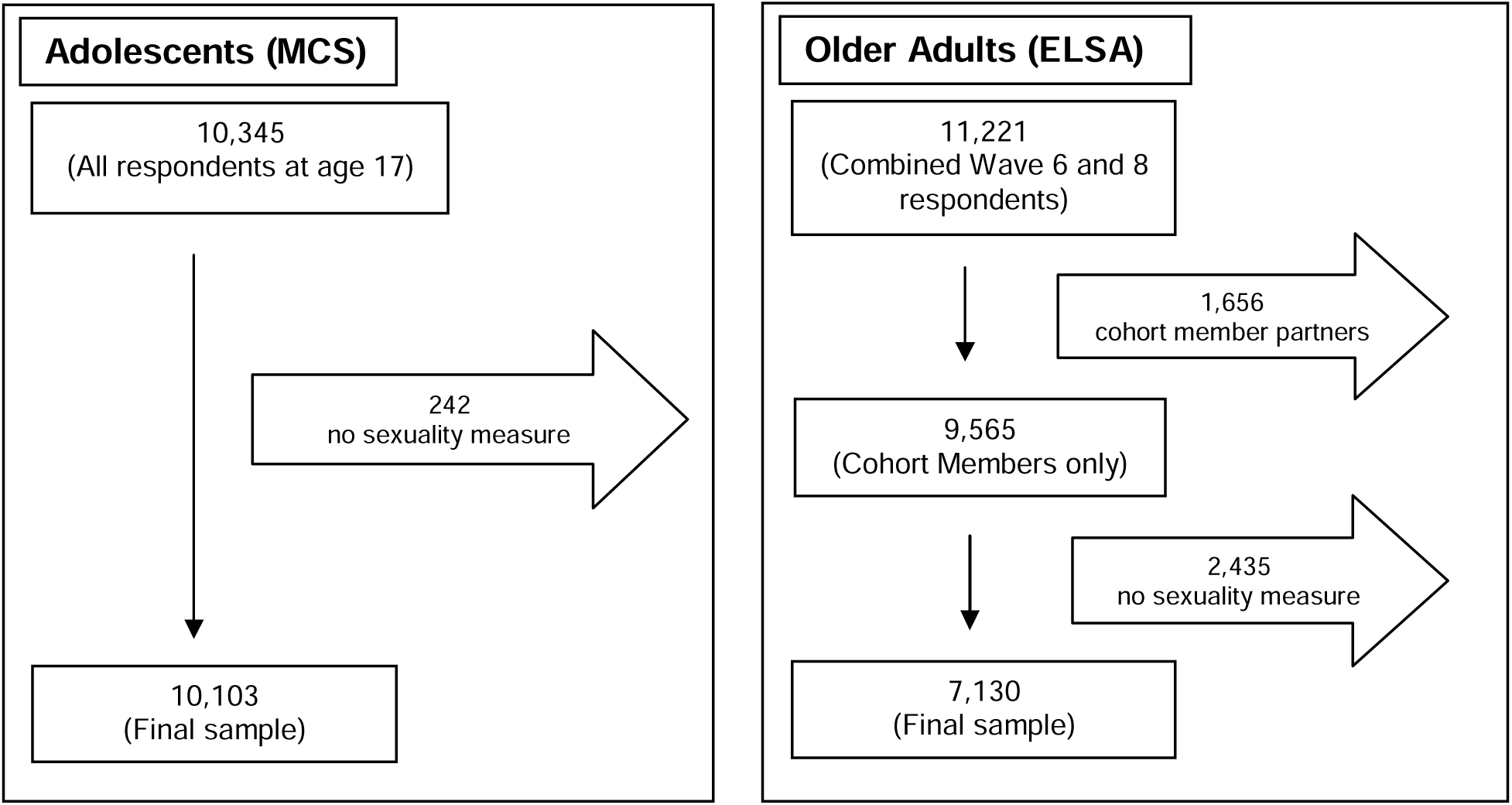

There was no difference in non-response between sexuality measures in MCS. However, sexual identity had a higher missingness than attraction or experience in ELSA. For example, 33.2% of the sample was missing a sexual identity response, compared to 14.5% missing an attraction response and 14.3% missing an experience response. This difference in missingness between measure is largely driven by attrition between wave 6 and 8 with 1,460 participants (20.5% of the sample) only responding at one wave. Those missing an identity response at wave 8 were more likely to be female, older (>70 years), and have lower socioeconomic status. Those missing an attraction or experience response at wave 6 were more likely to be female, younger (<70 years) and non-White.

## Analysis 1: Describe the dimensions of sexuality captured and investigate how they overlap in young people and older adults

We have measures of sexual identity and sexual attraction in adolescents captured at age 17. We have measures of these same dimensions in older adults (aged 50+), plus a measure of sexual experience. While the attraction measures are very similar between the datasets, the sexual identity measures differ in important ways which will be examined in more detail in the Discussion.

All measures, including the question and options used, are described in the variable section above and in Table 1.

### Descriptives for different dimensions of sexuality

The majority of participants in both adolescent and older adult samples report their sexual identity as “Completely Heterosexual” or “Heterosexual”, and their sexual attraction and experience as exclusively with the opposite sex. For example, 94.7% of older adult respondents identify as “Heterosexual”, 93.0% report attraction only to the opposite sex, and 94.8% report sexual experience with opposite sex. Likewise, 76.3% of adolescent respondents identified as “Completely Heterosexual”, rising to 86.9% if “Mainly Heterosexual” respondents are included, and 75.8% report attraction only to the opposite sex.

By contrast, less than 1% of older adults reported their sexual identity as either Gay/Lesbian or Bisexual. A larger proportion of adolescents identified as Gay/Lesbian (2.4%) or Bisexual (6.3%), but also a slightly higher percentage identified as “Other” (1.5% vs 0.6% in older adults). While adolescent respondents are more likely to report non-Heterosexual identity and same-sex attraction than older adults, it was still the minority (Table 1).

### Dimension Overlap

Heterosexual identified adolescents and older adults reported high coincidence between their reported identity and attraction/behaviour. For example, 93.0% of “Completely Heterosexual” adolescents and 94.5% of “Heterosexual” older adults reported only opposite-sex attraction (see Figure 1A & 1C).

**Figure 1:**
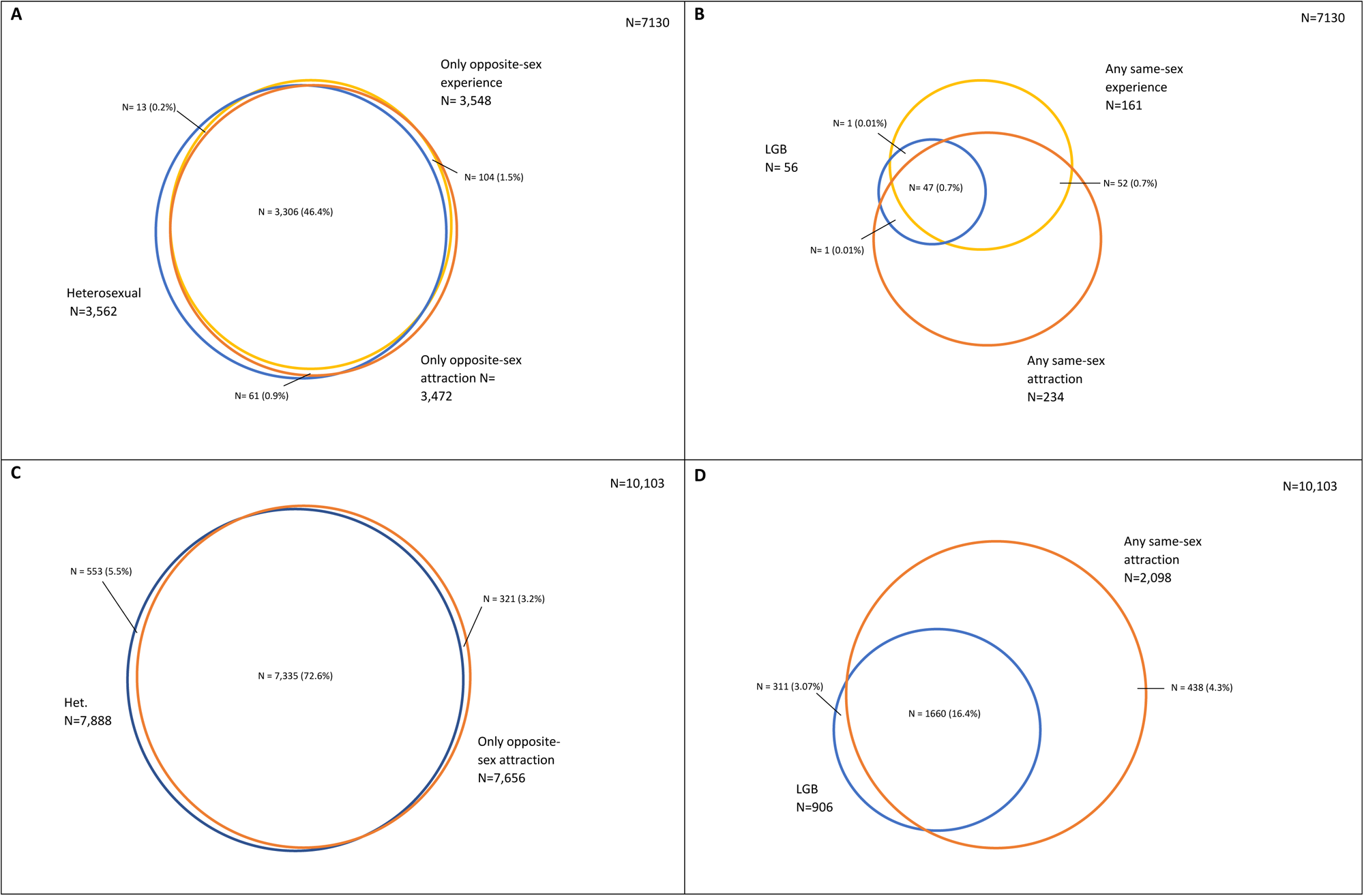
Overlap between A) Heterosexual identified older adults and those who report only opposite-sex attraction and experience, B) LGB identified older adults and those who report any opposite-sex attraction and experience, C) Heterosexual identified adolescents and those who report only opposite-sex attraction, and D) LGB identified adolescents and those who report any same-sex attraction. Blue corresponds to sexual identity measures, orange to sexual attraction measures, and yellow to sexual experience measures. All complete case.

By contrast, for those who reported “Gay/Lesbian” or “Bisexual” identities or same-sex attraction or experiences, coincidence was less the norm. For example, only 75.6% of “Completely Gay or Lesbian” adolescents reported “Only to same sex” attraction and 66.7% of “Gay or Lesbian” older adults reported only same-sex attraction and 47.1% reported only same-sex experience. Likewise, of those who reported at least some same-sex attraction, 74.0% of older adults also identified as “Heterosexual” and 17.7% of adolescents identified as “Completely Heterosexual” (see Figure 1B & 1D).

The collection of multiple dimensions also provides further context for respondents that might otherwise be dismissed as ambiguous (see Box 4). For example, older adult respondents who identified as “Other” were more likely to report their sexual attraction (65.2%) and experience (70.8%) as entirely with the opposite sex, whereas the majority of “Other” adolescents were evenly split between “Never sexually attracted” (40.8%) and reporting some same-sex attraction (39.5%). More detail is available in the supplementary.

### Analysis 2: Describe how choice of sexuality dimension impacts demographic distributions and health outcomes

#### Demographic distributions

##### Identity

A larger proportion of “Gay or Lesbian” and “Completely Gay or Lesbian” adolescents and older adults identified as male than the sample as a whole. This was also true of “Bisexual” older adults, while by contrast, a higher proportion of “Bisexual” adolescents identified as female than the sample as a whole. This is particular significant considering in the adolescent sample there are roughly equal proportions of male and female identified participants in the cohort, while the older adult sample was 55.3% female (Table 2).

**Table 2:**
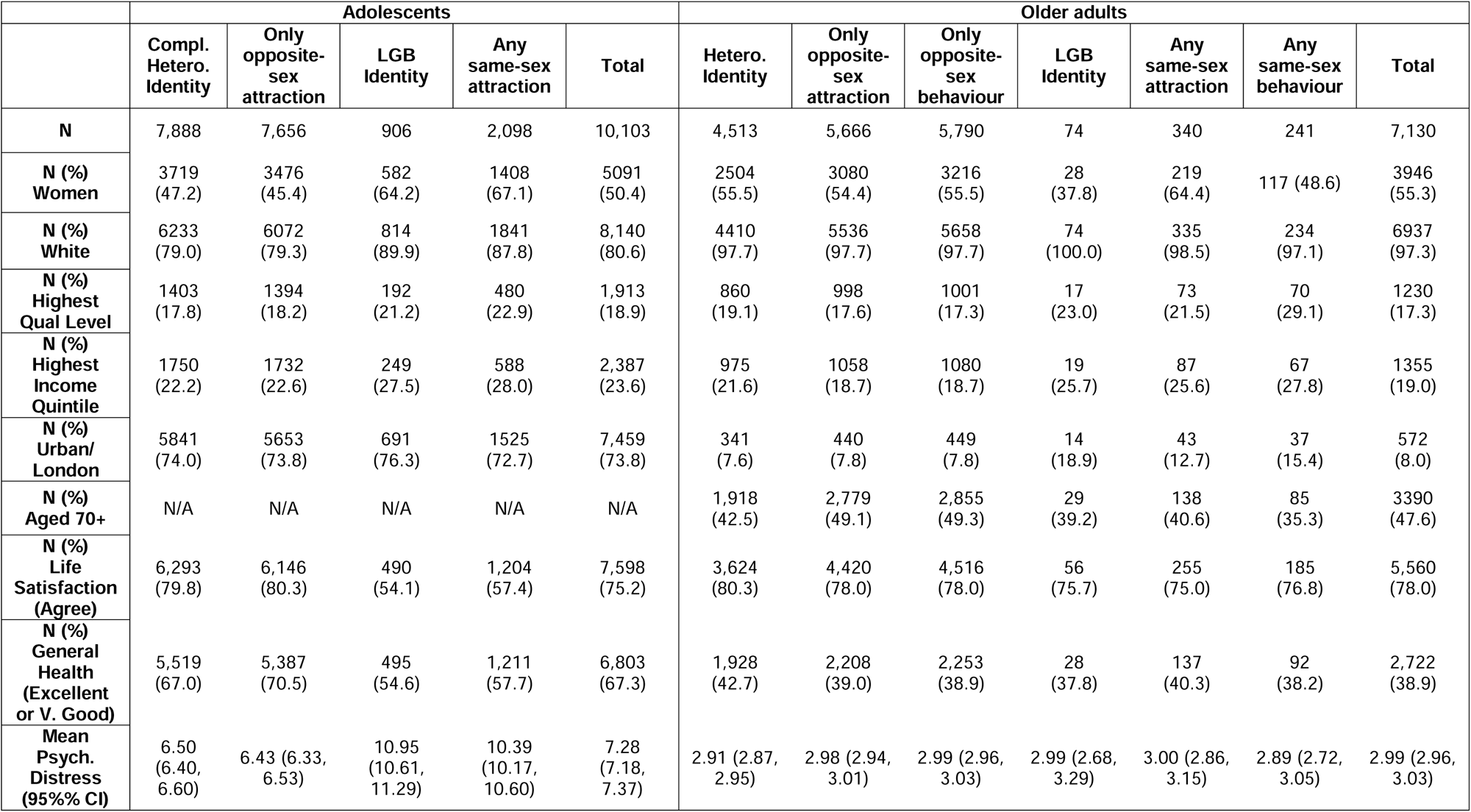
Demographic and health and wellbeing variables by sexuality in older adults and adolescents.

Regardless of age group, a higher proportion of “Bisexual” respondents were in the highest quintile for income and had a degree than the sample as a whole. This also held for “Gay/Lesbian” older adults. By contrast, the proportion of “Completely Gay or Lesbian” adolescents in the highest income quintile or with a parent with a degree was similar or lower than “Completely Heterosexual” adolescents and the sample as a whole. In both age groups, the proportion of those who reported their identity as “Other” or did not give an answer in the highest quintile for income or had a degree were lower than the sample as a whole (Figure 2).

**Figure 2:**
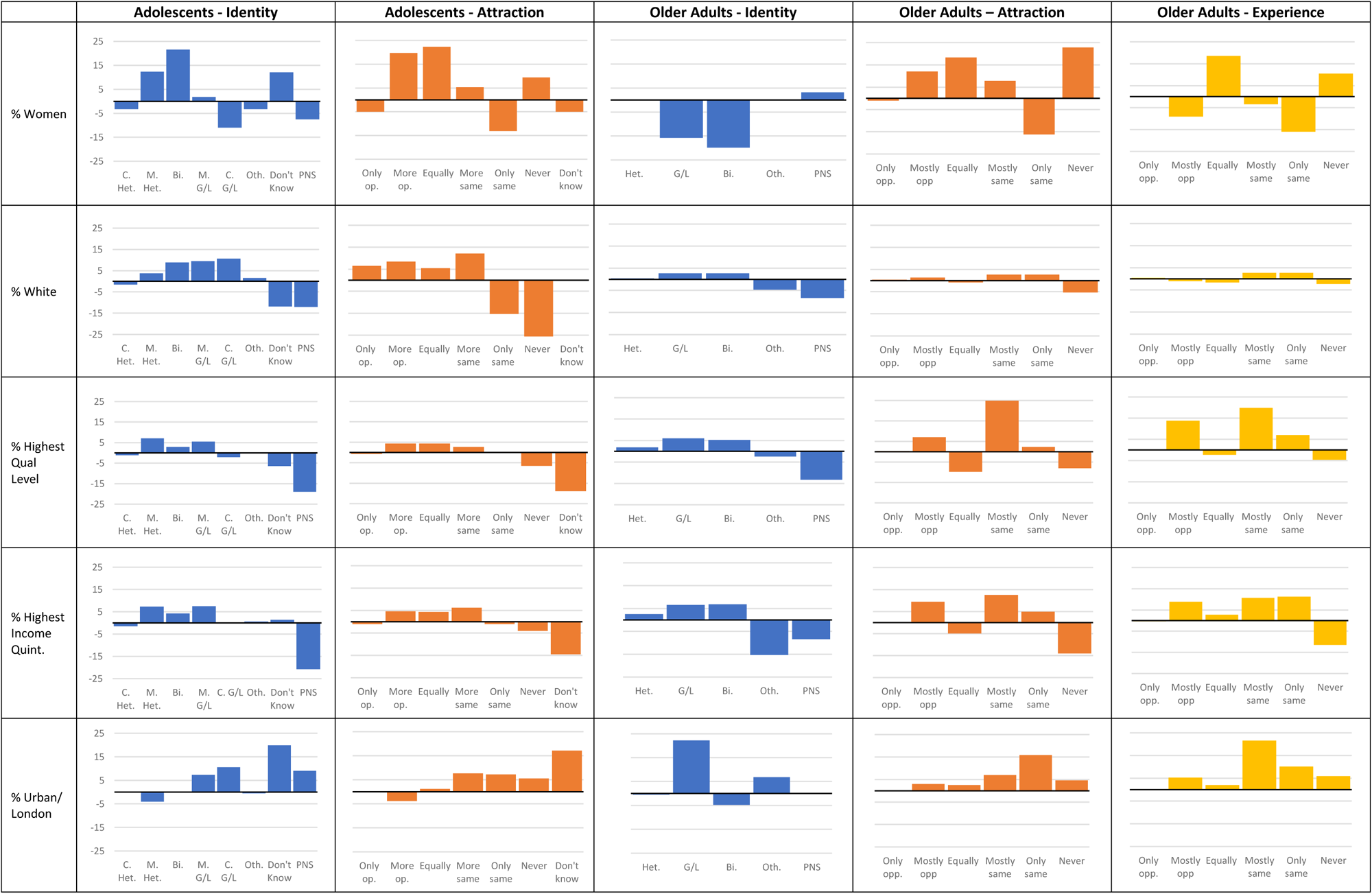
Demographic distributions by sexual identity, sexual attraction and behaviour in adolescents and older adults.

Finally, in both adolescents and older adults, a higher proportion of Lesbian/Gay participants resided in London or an urban area than their Heterosexual peers and the sample as a whole. However, while a similar proportion of Bisexual and “Mostly Heterosexual/Gay or Lesbian” adolescents resided in an urban area, a lower proportion of Bisexual older adults resided in London than Heterosexual older adults and the sample as a whole (Figure 2).

#### Attraction

In both adolescents and older adults, a larger proportion of those who reported exclusively opposite- or same-sex attraction identified as male than those who reported non-exclusive attraction (Figure 2).

In adolescents, those who reported exclusive attraction had similar income and education status, with those who reported non-exclusive attraction reporting slightly higher income and education than the exclusive attraction respondents. However, in older adults, those who reported any same-sex attraction reported higher income and education than those who reported only experiencing opposite-sex attraction. The exception was those who reported equal opposite- and same-sex attraction who reported lower education and income outcomes than the attraction categories (excluding never attracted) and the sample as a whole. Regardless of age group, those who reported never experiencing attraction or that they didn’t know, reported lower education and income than the sample as a whole (Figure 2).

Finally, a higher proportion of older adults who reported any same-sex attraction or no attraction resided in London than those who reported exclusively opposite-sex attraction. Likewise, a higher proportion of adolescents who reported “More often to same”, “Only to same” and “Never” attraction resided in an urban area than those who reported mostly opposite-sex attraction or the sample as a whole (Figure 2).

#### Behaviour

Excluding those who reported their behaviour as “equally” with the opposite- and same-sex, a higher proportion of older adults who report any same-sex behaviour identify as male, report higher income and education, and reside in London than those who report only opposite-sex experience and the sample as a whole (Figure 2). Respondents who reported no sexual experience followed a similar pattern to those who reported equal experience, reporting lower income and education, and a higher proportion of women than the other sexual experience categories (Figure 2).

#### Identity vs Attraction vs Behaviour

In general, regardless of dimension, a higher proportion of those who reported a non-“Heterosexual” identity or any same-sex attraction or experience also reported higher education and income, and resided in an urban area (Figure 2). In adolescents, a higher proportion of this group identified as female than the sample as a whole, while in older adults only a higher proportion of those with same-sex attraction or experience (but not LGB identity) identified as female.

Finally, across the dimensions, a lower proportion of those reporting “Other” identity, no attraction or experience and those who did not answer were also in the higher income and education categories, identified as “White”, or lived in a non-urban environment than the sample as a whole (Table 2). More detail is available in the supplementary.

#### Health and wellbeing

The proportion of adolescents who reported good life satisfaction or self-rated health was much lower among those who did not identify as “Completely Heterosexual” or who reported any attraction other than “Only to Opposite” (Figure 3). Similarly, mean psychological distress was worse for those same groups, compared to those who identified as “Completely Heterosexual” or reporting “Only to Opposite” attraction.

**Figure 3:**
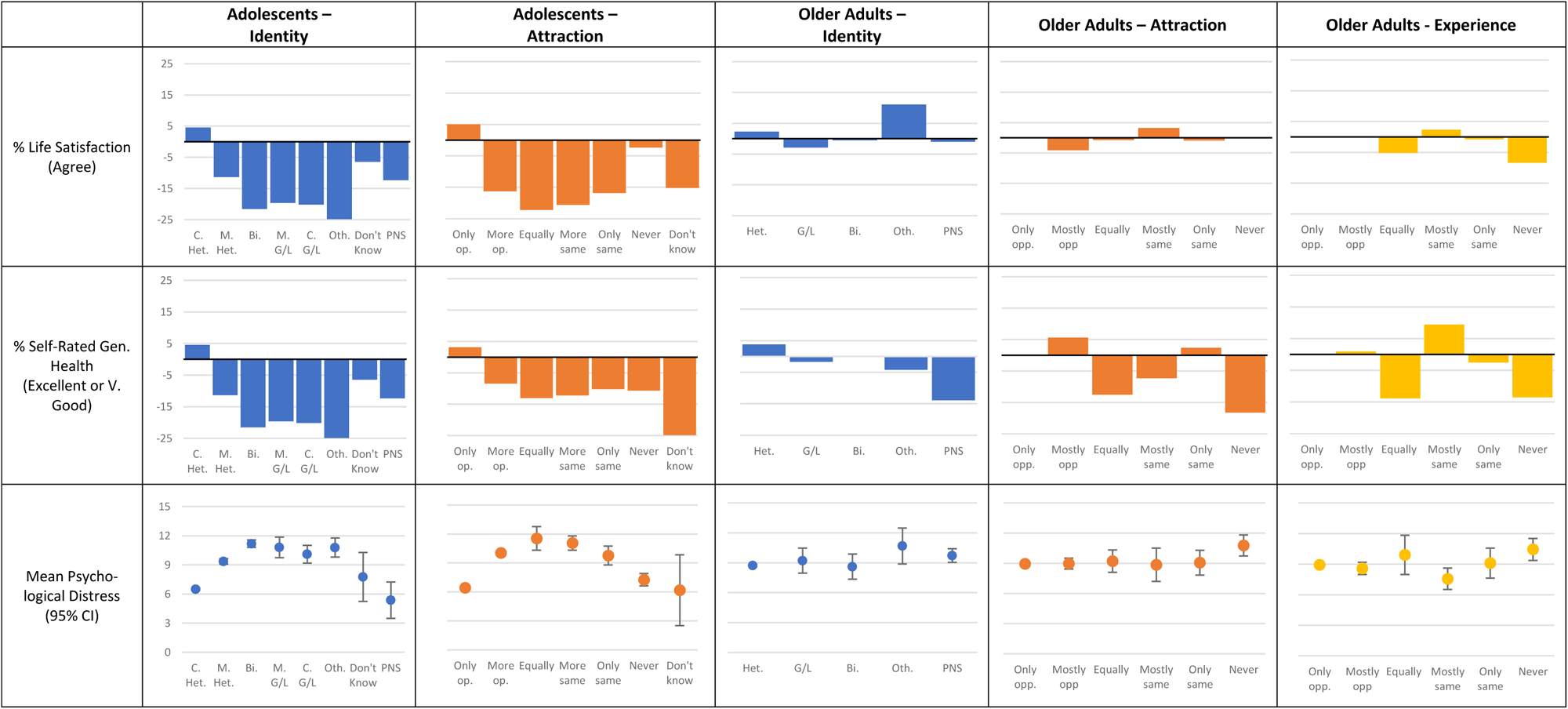
Health outcomes by sexual identity, sexual attraction and behaviour in adolescents and older adults

In older adults, psychological distress and the proportion who reported good life satisfaction was similar regardless of identity, attraction or experience (Figure 3). Likewise, self-rated health did not differ substantially by sexual identity in older adults. However, the proportion of older adults reporting equal or no attraction/experience who report good self-rated health was lower than all of the other groups and the sample as a whole (Figure 3).

##### Identity vs Attraction vs Behaviour

Regardless of dimension choice, sexual minority adolescents had worse outcomes on life satisfaction, self-rated general health, and psychological distress than the heterosexual sample (Table 2). However, this trend is mostly only present in the adolescent sample, with outcomes by identity, attraction, and experience similar or better in older adults. More detail is available in the supplementary.

### Analysis 3: Describe how recoding sexuality dimensions for analysis impacts analytic sample, demographic distributions (i.e. sex/gender, SES, and ethnicity), and health outcomes

As expected, the Broad recoding strategy resulted in much larger analytic samples than a restricted recoding, regardless of age group or dimension. For example, older adults categorized as sexual minority rose from 74 to 253 and adolescents rose from 906 to 2215 as a result of the Broad recoding of sexual identity. Likewise, Broad recoding of sexual attraction led to the sexual minority analytic sample size rising from 122 to 427 older adults (83 to 320 older adults using experience) and from 716 to 2446 adolescents. Finally, a Broad recoding of the combined dimensions resulted in an increased sexual minority analytic sample from 84 to 704 older adults and 480 to 2746 adolescents. See the supplementary for more detail.

#### Demographic distributions

##### Identity

In adolescents, using the Broad coding scheme results in a sexual minority sample in which a largely similar proportion identify as female (61.9% vs 64.2%) or white (86.1% vs 89.9%), are in the highest quintile for household income (28.6% vs 27.5%) or have a parent with a degree (23.0% vs 21.2%), and live in an urban area (73.1% vs 76.3%) than that produced using the Restricted coding scheme (Figure 4). However, regardless of coding scheme, a higher proportion of sexual minority adolescents likely to identify as female or White, and are in the highest quintile for household income or have a parent with a degree than non-sexual minority adolescents and the sample as a whole.

**Figure 4:**
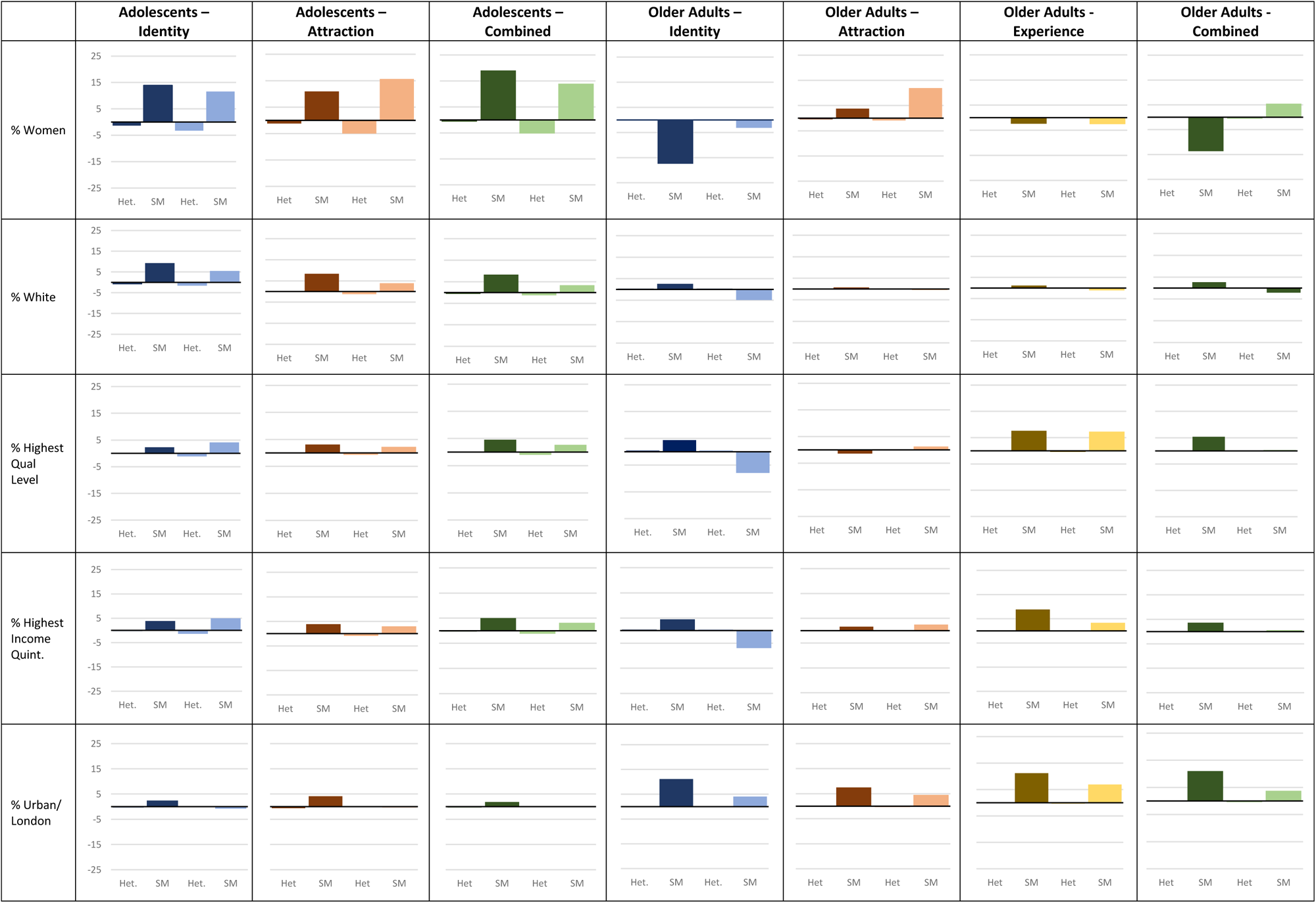
Demographic distributions by Restricted and Broad coding schemes in adolescents and older adults

In older adults, using the Broad coding scheme results in a sexual minority sample where a larger proportion identify as female (37.8% vs 52.2%) and live in London (11.9% vs 18.9%) but fewer are in the highest quintile for income (14.2% vs 25.7%) and have a degree or equivalent (10.7% vs 23.0%) (Figure 4). In fact, the Broad coding scheme resulted in a sexual minority sample where fewer are in the highest quintile for income and have a degree or equivalent than the sample as a whole, whereas a greater proportion of the Restricted sexual minority sample was in the highest quintile for income and had a degree or equivalent than the sample as a whole. There was no difference in the percentage identifying as White. However, regardless of coding scheme, a lower proportion of sexual minority older adults identified as female and a higher proportion lived in London than non-sexual minority older adults and the sample as a whole.

#### Attraction

In adolescents, using a Broad coding scheme results in a sexual minority sample a similar proportion identifying as female (66.0% vs 61.3%), and White (84.5% vs 89.0%) or living in an urban area (73.8% vs 77.9%) than that produced using a Restricted coding scheme (Figure 4). There was little to no difference in the proportion in the highest quintile for household income or having a parent with a degree by recoding scheme. However, regardless of coding scheme, sexual minority respondents are still more likely to identify as female or White, and more likely to be in the highest quintile for household income or have a parent with a degree than non-sexual minority adolescents and the sample as a whole.

In older adults, using a Broad coding scheme results in a sexual minority sample more likely to identify as female (67.2% vs 59.0%) and slightly less likely to live in London (12.7% vs 15.6%) than that produced using a Restricted coding scheme (Figure 4). There was little to no difference in likelihood of identifying as White, being in the highest quintile for income or having a degree or equivalent by recoding scheme. However, regardless of coding scheme, sexual minority older adults were less likely to identify as female and more likely to live in London than non-sexual minority older adults and the sample as a whole.

#### Experience

In older adults, using a Broad coding scheme results in a lower proportion of the sexual minority sample in the highest quintile of income (22.2% vs 27.7%) or living in London (15.0% vs 19.3%) than that produced using a Restricted coding scheme (Figure 4). There was little to no difference in the proportion identifying as Female or White, in the highest quintile for income or having a degree or equivalent by recoding scheme. However, regardless of coding scheme, sexual minority older adults were more likely to have a degree or equivalent, be in the highest quintile of income, and live in London and less likely to identify as female than non-sexual minority older adults and the sample as a whole.

#### Combination of Dimensions

In adolescents, using a Broad coding scheme results in a sexual minority sample in which a slightly smaller but similar proportion identify as female (64.3% vs 69.4%) or white (84.0% vs 89.0%), are in the highest quintile for household income (26.8% vs 28.8%) or have a parent with a degree (21.6% vs 23.5%), or live in an urban area (73.9% vs 75.6%) than that produced using a Restricted coding scheme (Figure 4). However, regardless of coding scheme, sexual minority respondents are still more likely to identify as female or White, and more likely to be in the highest quintile for household income or have a parent with a degree than non-sexual minority adolescents and the sample as a whole.

In older adults, using a Broad coding scheme results in a larger proportion of the sexual minority sample identifying as female (60.8% vs 41.7%), but a smaller proportion in the highest income quintile (19.6% vs 22.6%), with a degree or equivalent (17.6% vs 22.6%) and living in London (11.8% vs 19.1%) than that produced using a Restricted coding scheme (Figure 4). In fact, the Broad coding scheme resulted in a sexual minority sample more likely to identify as female than the sample as a whole, whereas the Restricted sample was less likely to identify as female than the sample as a whole. However, regardless of coding scheme, sexual minority respondents are more likely to live in London than non-sexual minority adolescents and the sample as a whole.

#### Health and wellbeing

##### Identity

In adolescents, using a Broad coding scheme results in a similar proportion of the sexual minority sample agreeing they are satisfied with life (59.9% vs 54.1%), rate their health as Excellent or Very Good (58.0% vs 54.6%), and report similar psychological distress (10.05 [95% 9.84-10.27] vs 10.95 [95% 10.61-11.29]) than that produced using a Restricted coding scheme (Figure 5). However, regardless of coding scheme, sexual minority adolescents report higher psychological distress and a greater proportion agree they are satisfied with their life or rate their health as Excellent or Very Good than non-sexual minority adolescents and the sample as a whole.

**Figure 5:**
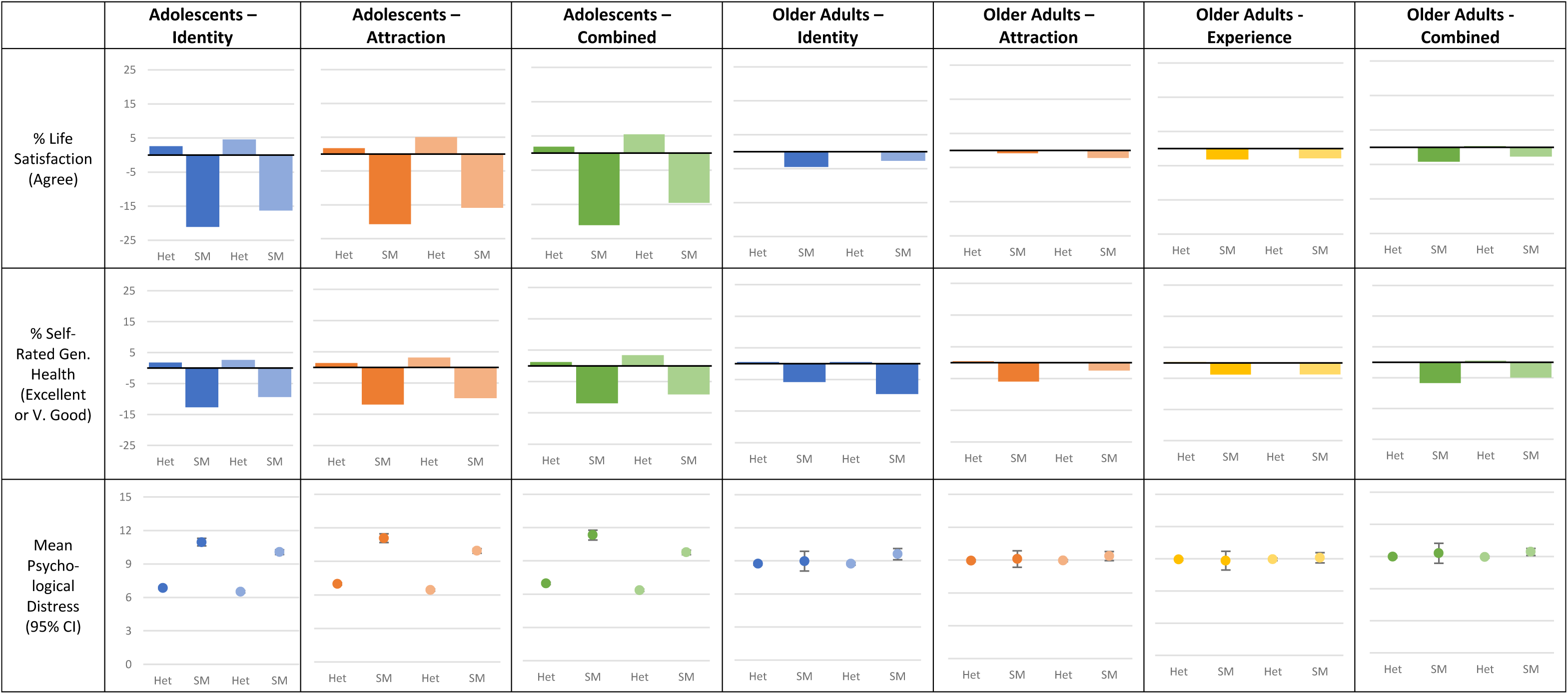
Health and wellbeing by Restricted and Broad coding schemes in adolescents and older adults

In older adults, using a Broad coding scheme results in a sexual minority sample of which a similar proportion are satisfied with life (77.5% vs 75.7%), rate their health as Excellent or Very Good (53.0% vs 56.8%), and who report similar psychological distress (3.2 [95% 3.02-3.37] vs 2.99 [95% 2.69-3.29]) than that produced using a Restricted coding scheme (Figure 5). Regardless of coding scheme sexual minority older adults are less likely to agree they are satisfied with their life or rate their health as Excellent or Very Good than non-sexual minority older adults and the sample as a whole.

#### Attraction

In adolescents, using a Broad coding scheme results in a sexual minority sample in which a higher but similar proportion are satisfied with life (59.4% vs 54.5%), rate their health as Excellent or Very Good (57.5% vs 55.5%), and report better psychological distress (9.93 [95% 9.72-10.13] vs 11.07 [95% 10.68-11.46]) than that produced using a Restricted coding scheme (Figure 5). Regardless of coding scheme, sexual minority adolescents report higher psychological distress and a lower proportion agree they are satisfied with their life or rate their health as Excellent or Very Good than non-sexual minority adolescents and the sample as a whole.

In older adults, there was little to no difference in life satisfaction, self-rated health, or psychological distress by recoding scheme by attraction or experience (Figure 5). However, regardless of coding scheme, sexual minority older adults are less likely to agree they are satisfied with their life or rate their health as Excellent or Very Good than non-sexual minority older adults and the sample as a whole.

#### Combination of Dimensions

In adolescents, using a Broad coding scheme results in a higher proportion of the sexual minority sample agreeing they are satisfied with life (60.7% vs 54.2%), rating their health as Excellent or Very Good (58.3% vs 55.4 %), and they report better psychological distress (9.76 [95% 9.56-9.95] vs 11.32 [95% 10.88-11.76]) than that produced using a Restricted coding scheme (Figure 5). However, regardless of coding scheme, sexual minority adolescents report higher psychological distress and are less likely to agree they are satisfied with their life or rate their health as Excellent or Very Good than non-sexual minority adolescents and the sample as a whole.

In older adults, there was little to no difference in life satisfaction, self-rated health, or psychological distress by Broad or Restricted recoding scheme (Figure 5). However, regardless of coding scheme, sexual minority older adults were less likely to agree they are satisfied with their life or rate their health as Excellent or Very Good than non-sexual minority older adults and the sample as a whole.

#### Sensitivity analyses

A sensitivity analysis was also performed using sex-assigned-at-birth rather than gender identity in MCS and using wealth rather than income in the older adult sample but found no difference in outcomes.

## Discussion

In this analysis, we provide the example of two longitudinal studies from the UK which have captured multiple dimensions (identity, attraction and experience) of sexuality. We aimed to demonstrate the impact of dimension choice and researcher coding decisions on analytic sample, demographic distributions and health outcomes. Firstly, we will discuss patterns of response coincidence between dimensions. Secondly, we will discuss the impact of dimension choice and coding decisions on analytic sample, demographic distributions and health outcomes. Finally, we reflect on the measures used in this analysis and how they inform interpretation.

### Overlap between dimensions

We found that Heterosexual identified adolescents and older adults were characterized by concordance between sexual orientation dimensions with almost all reporting only attraction to and experience with people of the opposite sex. Likewise, the majority of those who did not identify as Heterosexual/Completely Heterosexual reported same-sex attraction/experience or no attraction/experience. However, reporting of attraction and experience with same- and opposite-sex people was more common among Gay and Lesbian identified adolescents and older adults indicating a lower level of concordance between the dimensions in this group.

Likewise, a significant proportion of those reporting same-sex attraction or experience also identified as Heterosexual, particularly in older adults. These results demonstrate the non-equivalence between different dimensions of sexual orientation observed elsewhere, especially among sexual minorities (13). They also highlight the capture of multiple dimensions can provide a greater understanding of sexual diversity in the general population (see Box 3 for more).

### Impact of dimension and coding decisions

Not only do we see diversity in how sexual orientation is expressed in the datasets, we also see that demographics and health outcomes vary by the category under examination and dimension and recoding can substantially impact who is grouped under the sexual minority umbrella.

#### Analytic sample

As discussed above, the proportion of adolescents and older adults who report some same-sex attraction or experience is higher than those who identify as LGBTQ+ (13). While demonstrating the different dimensions of sexual orientation captured by these measures, this has important implications for the size of the analytic sample available to researchers. The larger analytic sample afforded by the attraction measure is appealing for statistical power, however the operationalisation of attraction is challenging as it rarely sits in a wider conversation about discrimination and disadvantage (16). Attraction measures inform on sexual diversity and development, but they may be too imprecise and removed from the mechanisms of disadvantage to estimate a meaningful sexual minority population. Similarly, the proportion of older adults reporting same-sex experience is higher than those identifying as LGBTQ+ in our analysis and the measure has clear importance in sexual health research. However, defining the sexual minority population exclusively by sexual experience/behaviour can be reductive and, like attraction, fail to capture wider social and political contexts. As a result, sexual identity has been repeatedly recommended as a key measure for population surveys (6,16,17). While resulting in a smaller analytic sample, sexual identity allows for a more legible estimate and speaks to wider political landscape and community. Nonetheless, the use of reported sexual identity limits the estimated population solely to those who have made themselves visible and positioned themselves in a way that maps on to the idea of a “LGBTQ+ community”. As such, those alienated from such ideas, or who are excluded or vulnerable in other ways, are left out.

This same tension between the complex and multi-dimensional nature of sexual orientation and the need to make quantitative research clear, testable, and legible to wider stakeholders and policy makers is present in our recoding analysis. As discussed in Box 2, the Broad and Restricted strategies were designed to illustrate approaches at two ends of a continuum with the Broad strategy including a wide range of responses across the three dimensions in the analysis while the Restricted strategy prioritised identification with a lesbian, gay or bisexual identity and concordant attraction/experience. As expected, the Broad strategy resulted in a larger analytic sample than the Restricted strategy, however as discussed above, it’s inclusive framework may limit its usefulness.

Overall, dimension choice and recoding can have significant impact on the analytic sample, particularly of sexual minority categories. As a result, careful consideration by researchers is essential.

#### Demographic distributions

Little substantial variation in demographic distribution by dimension or recoding was observed in either study with a higher proportion of sexual minority respondents being younger, more urban, high income, and high education. However, some differences in gender, age, and socioeconomic outcomes were observed between and within cohorts.

In adolescents, a higher proportion of sexual minority respondents identified as female than their heterosexual peers, whereas a higher proportion of sexual minority older adults identified as male. One potential cause of this difference between the age groups is the difference in the proportion of sexual minority respondents identifying as gay or lesbian or reporting exclusively same-sex attraction/experience. In both adolescents and older adults, monosexual sexual minorities were more likely to identify as male than bisexual respondents or those who report both same- and opposite-sex attraction/experience. Likewise, Geary et al (2018) found a higher proportion of men identified as gay/lesbian than women, while a higher proportion of women identified as bisexual or reported plurisexual experience than men (13). A higher proportion of sexual minority older adults in our analysis identified as gay or lesbian than adolescents which may explain why the older adult sexual minority sample is more likely to be male, despite the higher proportion of female respondents in the sample as whole. However, these changes may instead be due to shifts in sexual identity and attraction responses by age, cohort or time period. That there may be some difference due to the age or generation of the two groups is supported by the shift in sexual orientation outcomes by age. Most notably, a much higher proportion of adolescents identify as gay, lesbian or bisexual and report same-sex attraction than the older adults. In addition, those older adults who did report an LGBTQ+ identity or same-sex attraction/experience were younger than those who reported exclusively opposite-sex attraction/experience and a heterosexual identity. These patterns follow an observed trend of younger adults being more likely or more willing to report sexual minority identity, attraction, and experience reflecting changes in the social and political landscape (25). Additionally, we may be observing a survivor bias in the proportion of sexual minority older adults beyond a difference in disclosure or identification. For example, experience of historic and contemporary structural and interpersonal discrimination, as well as the health and community consequences of the HIV/AIDS epidemic, impacts the health and wellbeing of older LGBTQ+ adults potentially resulting in greater attrition from population studies and mortality (37–39).

In our analysis, sexual minority adolescents and older adults reported higher income and education than non-sexual minority participants and held regardless of dimension or recoding strategy or dimension of socioeconomic status. This matches recent evidence which suggests that some groups, such as older LGBTQ+ women, may have higher educational attainment and income than their heterosexual peers, however this appears to vary by specific sexual identity, generation and gender (40–43). The variation by identity was also present in our analysis with socioeconomic status varying between monosexual and plurisexual respondents. While monosexual sexual minority adolescents reported similar characteristics to their non-sexual minority peers, they reported worse socioeconomic status than plurisexual sexual minority adolescents. By comparison, both monosexual and plurisexual sexual minority older adults had better socioeconomic status than non-sexual minority older adults. These results may reflect differences in likelihood and/or willingness to report a plurisexual identity or attraction by socioeconomic status. It is also worth noting that socioeconomic status for the adolescent sample is that of their parents or household so may reflect their socioeconomic background, rather than the impact of sexual minority status on education or income. By contrast, socioeconomic status in the older adults may reflect more explicitly the impact of sexual minority status over the lifecourse. A high proportion of sexual minority older adults identified as male than the non-sexual minority group, which due to the age profile of the cohort, may results in participants with higher income/wealth and educational qualification characteristics. As well as demographic differences, we also observed differences in health and wellbeing outcomes.

#### Health and wellbeing

Consistent with the literature on health in sexual minority populations (1–3), sexual minority adolescents reported worse outcomes in life satisfaction, and psychological distress than their non-sexual minority peers, while both sexual minority adolescents and older adults reported worse self-rated health. This disparity has been attributed to differences in health behaviours, the impact of structural and interpersonal experiences of discrimination (i.e. minority stress), and poor experiences with healthcare and related services (44,45). Across the three outcomes, the disparity between sexual minority and non-sexual minority participants was more acute in the adolescent study than in the older adult study. It is unclear if this is a product of differences between the datasets in terms of sample sizes, measures and sampling framework, or reflects a narrowing of inequality over the lifecourse. One crucial factor may be the relative youth of sexual minority respondents in the older adult study compared to non-sexual minority respondents and suggests adjusting for age may be important in future studies. While these outcomes did not substantially vary by dimension of sexuality employed or by recoding, the inclusion or exclusion of the “Mostly Heterosexual” category from the sexual minority category did affect the severity of the difference observed in the adolescent sample (see Box 3).

### Reflections on measurement

Finally, in addition to our examination of the impact of dimension and recoding choices on analytic sample, demographics, and health outcomes, we reflect on the measures used in this analysis and how they inform interpretation of the analysis presented.

#### Identity

Both datasets include a measure of sexual identity, however the chosen measures reflect two different conceptual understandings of the dimension.

The identity measure in ELSA is similar to that present in other UK population surveys and is similar to the measure used in the 2021 Census in England and Wales (16,17). As a result, it is useful for cross-dataset analysis and comparison. The categories included are in common use and their salience to the general population has been verified (20). This measure is constructed of strictly nominal categories with identity labels presented as bounded and independent.

By comparison, the measure in MCS tries to transform sexual identity into an ordinal categorical variable ranging from “Completely Heterosexual” to “Completely Gay/Lesbian” with ‘Mainly Gay/Lesbian” and “Mainly Heterosexual” options. This shift, despite the similar categories, represents a fundamental change from considering sexual identity as discrete labels to a continuum or spectrum. Issues introduced due to this change are expanded on in Box 3.

The inclusion of this measure was suggested on the grounds of a more nuanced approach to sexual identity and to allow those who might otherwise select “Other” to expand on their answer (private communication, see Box 3). However, it is unclear what is meant by the qualifiers ‘completely’ or ‘mainly’ in the measure and if understanding is consistent between researcher and respondent, especially as they relate to identities that are employed in real world contexts. In addition, the idea of sexuality as a spectrum with heterosexuality and homosexuality as opposite binary ends has been challenged by some researchers of sexuality (6).

Secondly, the positioning of “Bisexual” as a midpoint between “Completely Heterosexual” and “Completely Gay/Lesbian” potentially reinforces ideas of bisexuality as ‘half-straight/half-gay’, requiring equal attraction to men and women, and/or as a label which inherently supports binary understandings of gender - ideas which have been used by some to erase or discredit bisexuality as an identity (46). Additionally, while there has been increased interest in those that report themselves mostly or mainly heterosexual, it is unclear if equal research interest has been expressed in those identifying as “Mainly Gay or Lesbian” (See Box 2)(47). The measure is difficult to harmonise with other measures of sexual identity making it less useful for cross-dataset analysis and comparison. As demonstrated in this analysis, a dichotomized sexual minority recoding of the measure does allow for some comparability, however the construction of the binary does require careful consideration of the ordinal item structure.

Finally, the timing of collection of sexual identity in the two studies reflect researcher assumptions about the sexual identity of the participants in each study. For example, that the question was first included at age 17 in MCS perhaps reflects concerns around asking about sexual identity at an earlier age, but also the position of MCS as a research tool developed in a UK context which is willing to ask about sexuality and is mindful of an increasingly non-heterosexual identified youth population (48). By contrast, ELSA respondents were only asked about sexual identity in the eighth wave of the study (2017-18), two waves after the first questions on sexual attraction and experience, with no plans to ask again in the near future (private communication). The timing of the inclusion of the question in ELSA reflects the ways in which sexuality, in particular non-heterosexual identities, are deprioritised in research of this age group. For example, sexuality measures are currently missing from key UK studies of ageing such as the 1946 National Study of Health and Development (NSHD) and the 1958 National Child Development Study (NCDS), and older adults are omitted from the recent collection of the British National Surveys of Sexual Attitudes and Lifestyles (NATSAL) (49,50). These trends sit within popular understandings of sexuality as fixed by older age, and perhaps researcher concerns around non-response and question appropriateness for this age group (51). However, these concerns can result in the erasure and understudy of older sexual minorities and the sexuality of older adults in general.

Overall, MCS and ELSA demonstrate two approaches to the measurement of sexual identity and reflect the importance of careful consideration of how identity is conceptualised and how different populations may receive the measure.

#### Attraction and behaviour

The conceptualisation and acceptability of a measure is also an important consideration when measuring sexual attraction and behaviour. Unlike sexual identity, sexual attraction and experience measures do not require individuals to have taken on a specific social identity and may have a lower ‘barrier to entry’ than identity measures. As a result, measuring attraction and behaviour may capture individuals whose sexual identity is developing or flexible, or who may not identify as a member of the LGBTQ+ community or the categorisations presented in identity questions.

While ELSA respondents have been asked about their experience of sex at multiple waves, the orientation of their sexual attraction and experience was only asked once at wave 6 (2012-13). By contrast, MCS respondents have been asked about their sexual attraction at age 14 and again at age 17. Like sexual identity, the choice to repeat this measure or not reflects assumptions about the target population. For example, that sexuality will be changeable at younger ages but fixed at older, the acceptability of the question to different age groups, and the prioritisation of research of sexuality in older and younger cohorts.

A measure of sexual experience is only included in ELSA. While MCS collects information on romantic partners and questions designed to identify sexual behaviour which carries a risk of pregnancy or STIs, the survey does not include questions on the gender of partners. As well as prioritising a conception of sex centred on risk, the selection of measures limits the usefulness of the dataset for understanding the sexuality and sexual behaviour of the target population.

An important element of measures of sexual attraction and behaviour is the time period participants are asked to consider. Although addressing different ends of the lifecourse, MCS and ELSA both ask about lifetime or ‘ever’ attraction and experience rather than a specific time frame. While this framing potentially allows for the consideration of the general pattern of attraction and experience in an individual’s life, it is subject to issues of inaccurate recollection and a social desirability/normative bias in reporting for both cohorts. This means the measure may not effectively capture participant’s ‘normal’ or routine attraction/experience patterns, nor attraction/experience patterns at a specific life stage. As a result, it is important to be clear in analysis about what responses to these measures can and cannot tell us.

As well as the time period participants are asked to consider, the presence of examples or explanatory text is an important element in a measure of sexual attraction/behaviour. Sexual attraction and behaviour experience are subjective and can incorporate a variety of meanings and forms (15). Where no further definition or explanation is given, as in MCS, this can open up inconsistency between researcher and participant understandings of what is being measured. In contrast, the measure in ELSA is phrased as follows: “Which statement best describes your sexual desires over your lifetime? Please include being interested in sex, fantasising about sex or wanting to have sex?” which supports a more consistent understanding between researcher and participant.

Likewise, both ELSA and MCS reinforce a gender binary in the items used to report attraction/experience by not providing other gender options for the objects of attraction (e.g. “only to males, never to females”). ELSA has not yet measured gender identity so uses a self-reported binary sex variable, while MCS respondents at age 17 were able to complete a measure of gender identity immediately before the measure of sexual attraction. However, due to survey routing, a participant who had identified themselves as non-binary but who was assigned female at birth would receive the same question phrasing as someone who reported their gender identity as “Female”. While inconsistent with the MCS survey’s acknowledgement of diverse gender identities, this routing also hampers interpretation in the data release. In the data published by the survey, all attraction responses are recoded into one variable with the terms ‘male’ and ‘female’ replaced with ‘same-sex’ and ‘opposite-sex’. As a result, a non-binary identified participant reporting attraction “only to the opposite-sex” can only be interpreted by referring their sex-assigned-at-birth, which obscures both their gender identity and reinforces a binary they may not identify with.

Overall, MCS and ELSA demonstrate two approaches to the measurement of sexual attraction and behaviour and reflect the importance of careful consideration of how these dimensions are conceptualised and how different populations may receive the measure.

## Conclusion

This study explored a range of practical and theoretical considerations when analysing sexual minority respondents using survey data. We highlight the impact recoding strategy decisions may have on analytic sample and demographic and health distributions. We also draw attention to the value of including multiple dimensions in population surveys, avoiding overzealous grouping or dropping of respondents, and the limits of coincidence between dimensions. However, we have also demonstrated how practical recoding and measurement decisions can be made thoughtfully and carefully We hope this analysis will empower and inform researchers interested in working on sexual minority health and social inequalities, as well as encouraging all researchers to critically examine how they construct and imagine their population of study.

### Limitations

The findings may only be applicable to the populations under analysis, however, the inclusion of two population-based studies covering different age groups we hope to illustrate the potential breath of considerations. Additionally, the analytic samples involved throughout this analysis are small, particularly for older respondents, which may have obscured important differences between the groups. This analysis also cannot discuss age or period effects due to question only being asked once in both cohorts; future examinations in other datasets or different timepoints of the same datasets would be valuable.

#### Box 1

**Key Takeaways**

- Sexual identity, behaviour and attraction measures are not interchangeable, and care should be taken when selecting a measure for inclusion in a survey or analysis.
- Dimension choice and data management of sexuality variables have important consequences for analytic sample and make-up, proposed mechanism of action, and health outcomes.

Including more than one dimension allows for the exploration of sexual diversity and help clarify otherwise ambiguous responses.

#### Box 3

**“Mainly Heterosexual” and “Mostly to Opposite”**

The MCS measure of sexual identity introduces new categories which may not correspond with common sexual identities including “Mainly Heterosexual” and “Mainly Gay/Lesbian”. In sample the “Mainly Heterosexual” identity category is the second largest after the “Completely Heterosexual” category. Evidence from the literature suggests this group is demographically distinct from both heterosexual and bisexual groups, and relatively stable across the lifecourse (47). This is supported by MCS where the “Mainly Heterosexual” group is distinct from the other identity categories, and 71.21% of the category also reported some same-sex attraction. Nonetheless, given their identification with the heterosexual label, they could also be reasonably included under the “Heterosexual” umbrella.

Where items include qualifiers such as ‘mostly’, researchers may then recategorize into fewer groups of ‘true’ gay, lesbians and bisexuals (8). Savin-Williams (2009) argues this raises ‘how much’ of a given dimension is needed for inclusion as non-heterosexual, especially as it interacts with methodological concerns around analytic sample and representativeness (8). For example, using the Broad recoding strategy, including “Mainly Heterosexual” respondents doubles the number of sexual minority participants from 906 to 2007 and as a result justification for inclusion should be carefully considered.

The Broad recoding strategy therefore results in a slightly more male, higher socioeconomic status, and less urban sexual minority sample. This group also reports better life satisfaction, self-rated health, and psychological distress outcomes than non-heterosexual identified respondents and those who report equal or more same-sex attraction, so when combined, health and wellbeing outcomes in the sexual minority group are better than when they are excluded.

While it is highly appealing to include these groups as sexual minorities as they share many qualities in common, it is important researchers are aware of the potential skew that they introduce.

#### Box 4

**“Other”**

Where a sexual identity measure includes the options ‘Other’, “Don’t Know”, and ‘Prefer Not to Say’ those responses are often excluded from analyses or marked as ‘missing’ (e.g. Geary *et al.*, 2018). Their exclusion is justified on two main grounds: small numbers, and uncertainty around what respondents are expressing when selecting these options. However, Booker et al. (2017) have argued strongly for their inclusion and consideration as groups worthy of further examination (19). These options may cover those who do not understand the question, who may not feel they fit within the available options, for example people who identify as asexual or queer, and those who reject labels altogether. (6,16).

Response patterns to these items can be illuminated by attraction and experience measures. For example, in adolescents almost 40% of “Other” respondents reported some same-sex attraction, with a further 40% reporting “Never sexually attracted”. This distribution indicates the “Other” category may be selected by sexual minority adolescents who use sexual identity labels outside of gay/lesbian/bisexual, such as pansexual, queer, or asexual, and reflects the developing sexuality of the cohort. By contrast, the majority of “Other” older adults reported only opposite-sex attraction and experience. Given “Other” older adults were more likely to be over 70, non-white, and lower SES than the sample as a whole, reporting of “Other” identity may reflect poor measure comprehension by respondents who might otherwise identify as heterosexual or straight. This difference observed between adolescent and older adult “Other” respondents points to the value of multiple dimension measurement, and consideration of target population when choosing a measure and data cleaning.

## Declarations

### Ethics approval and consent to participate

ELSA data collection was approved by the London Multicentre Research Ethics Committee (MREC/01/2/91), and informed consent was obtained from all participants.

Ethics approval for the MCS study was obtained from the National Research Ethics Service Committee London— Central (reference 13/LO/1786). All parents gave consent for their children to participate, and young people also provided verbal consent.

Ethics approval for this analysis was received from the Institute of Education, UCL.

### Availability of data and materials

MCS and ELSA data are available to all researchers, free of cost from the UK Data Service (https://www.ukdataservice.ac.uk)(30,52).

## Authors’ Contributions

ET conceptualized the study, developed the methodology, conducted the analyses, prepared the data visualization, and prepared the initial manuscript draft and subsequent revisions. DK and PP supervised the study conceptualization, the development of the methodology and the analysis, and reviewed and commented on the manuscript. All authors approve the final version and consent to its publication.

## Data Availability

MCS and ELSA data are available to all researchers, free of cost from the UK Data Service (https://www.ukdataservice.ac.uk).

## Acknowledgements

Not applicable.

