## Supplementary for "Mainly Heterosexual, Bisexual, or Other?: the measurement of sexual minority status and its impact on analytic sample, demographic distribution and health outcomes"

### Variable Overlap

#### Table S1: Attraction by sexual identity in adolecents

|  | **Completely heterosexual** | **Mainly heterosexual** | **Bisexual** | **Mainly Gay or Lesbian** | **Completely Gay or Lesbian** | **Other** | **Do Not Know** | **Prefer Not to Say** | **Total (%)** |
| --- | --- | --- | --- | --- | --- | --- | --- | --- | --- |
|  | N= 7,888 | N= 1,101 | N= 656 | N= 90 | N= 160 | N= 157 | N= 16 | N = 35 | N= 10,103 |
| **Attraction** |  |  |  |  |  |  |  |  |  |
| **Only to opposite sex (%)** | 7335  (92.99) | 283  (25.70) | 5  (0.76) | 0  (0) | <5  (<0.50) | 27  (17.20) | <5  (<20.00) | <5  (<7.50) | 7656  (75.78) |
| **More often to opposite sex (%)** | 361  (4.58) | 757  (68.76) | 243  (37.04) | <5  (<2.50) | 0  (0) | 17  (10.83) | <5  (<15.00) | <5  (<5.00) | 1382  (13.68) |
| **About equally (%)** | 6  (0.08) | 24  (2.18) | 316  (48.17) | 7  (7.78) | 0  (0) | 30  (19.11) | <5  (<7.50) | <5  (<5.00) | 385  (3.81) |
| **More often to same sex (%)** | 0  (0) | <5  (<0.50) | 72  (10.98) | 74  (82.22) | 33  (20.63) | 13  (8.28) | 0  (0) | 0  (0) | 194  (1.92) |
| **Only ever to same sex (%)** | <5  (<0.50) | <5  (<0.50) | <5  (<0.50) | 7  (7.78) | 121  (75.63) | <5  (<2.50) | 0  (0) | (0) | 137  (1.36) |
| **I have never felt sexual attraction(%)** | 160  (2.03) | 32  (2.91) | 17  (2.59) | <5  (<2.50) | <5  (<2.50) | 64  (40.76) | 7  (43.75) | 6  (17.14) | 291  (2.88) |
| **Do not know (%)** | <5  (<0.50) | 0  (0) | 0  (0) | 0  (0) | <5  (<2.50) | <5  (<2.50) | <5  (<15.00) | <5  (<7.50) | 11  (0.11) |
| **I do not wish to answer (%)** | 18  (0.23) | <5  (<0.50) | <5  (<0.50) | 0  (0) | 0  (0) | <5  (<2.50) | <5  (<7.50) | 23  (65.71) | 46  (0.46) |
| **No answer (%)** | - | - | - | - | - | - | - | - | <5  (<0.50) |

|  | **Only to opposite, never to same** | **More often to opposite, and at least once to a same** | **About equally often to opposite and to opposite** | **More often to same, and at least once to a opposite** | **Only ever to same , never to opposite** | **I have never felt sexually attracted to anyone at all** | **Do not know** | **I do not wish to answer** | **No answer** | **Total** |
| --- | --- | --- | --- | --- | --- | --- | --- | --- | --- | --- |
|  | N=7656 | N=1382 | N=385 | N=194 | N=137 | N=291 | N=11 | N=46 | N=1 | N=10,103 |
| **Completely heterosexual/ straight** | 7335 (95.81) | 361 (26.12) | 6 (1.56) | 0 (0.00) | <5  (<5.00) | 160 (54.98) | <5  (<40.00) | 18 (39.13) | - | 7888 (76.25) |
| **Mainly heterosexual/ straight** | 283 (3.70) | 757 (54.78) | 24 (6.23) | <5  (<2.50) | <5  (<2.50) | 32 (11.00) | 0 (0.00) | <5  (<5.00) | - | 1101 (10.64) |
| **Bisexual** | 5 (0.07) | 243 (17.58) | 316 (82.08) | 72 (37.11) | <5  (<2.50) | 17 (5.84) | 0 (0.00) | <5  (<2.50) | - | 656 (6.34) |
| **Mainly gay or lesbian** | 0 (0.00) | <5  (<0.50) | 7 (1.82) | 74 (38.14) | 7 (5.11) | <5  (<0.50) | 0 (0.00) | 0 (0.00) | - | 90 (0.87) |
| **Completely gay or lesbian** | <5  (<0.50) | 0 (0.00) | 0 (0.00) | 33 (17.01) | 121 (88.32) | <5  (<2.50) | <5  (<10.00) | 0 (0.00) | - | 160 (1.55) |
| **Other** | 27 (0.35) | 17 (1.23) | 30 (7.79) | 13 (6.70) | <5  (<2.50) | 64 (21.99) | <5  (<20.00) | <5  (<2.50) | - | 157 (1.52) |
| **Do not know** | <5  (<0.50) | <5  (<0.50) | <5  (<0.50) | 0 (0.00) | 0 (0.00) | 7 (2.41) | <5  (<20.00) | <5  (<2.50) | - | 16 (0.15) |
| **Prefer not to say** | <5  (<0.50) | <5  (<0.50) | <5  (<0.50) | 0 (0.00) | 0 (0.00) | 6 (2.06) | <5  (<20.00) | 23 (50.00) | - | 35 (0.34) |

#### Table S2: Sexual identity by attraction in adolescents

#### Table S3: Attraction by sexual identity in older adults

|  | **Heterosexual or Straight** | **Gay or Lesbian** | **Bisexual** | **Other** | **Prefer not to say** | **Missing** | **Total** |
| --- | --- | --- | --- | --- | --- | --- | --- |
| N= | 4513 | 43 | 31 | 27 | 152 | 2364 | 7130 |
| **Experience** |  |  |  |  |  |  |  |
| **Entirely with opposite** | 3443 (76.29) | 0 (0.00) | 8 (25.81) | 17 (62.96) | 89 (58.55) | 2233 (94.46) | 5790 (81.21) |
| **Mostly with opposite, but some experience with same** | 98 (2.17) | <5  (<5.00) | 11 (35.48) | <5  (<7.50) | <5  (<2.50) | 40 (1.69) | 158 (2.22) |
| **Equally with opposite and same** | <5  (<0.50) | <5  (<7.50) | <5  (<10.00) | 0 (0.00) | <5  (<2.50) | 13 (0.55) | 23 (0.32) |
| **Mostly with same, but some experience with opposite** | <5  (<0.50) | 11 (25.58) | <5  (<5.00) | 0 (0.00) | <5  (<2.50) | 13 (0.55) | 27 (0.38) |
| **Entirely with same** | <5  (<0.50) | 16 (37.21) | 0 (0.00) | 0 (0.00) | 0 (0.00) | 13 (0.55) | 33 (0.46) |
| **No sexual experience in lifetime** | 22 (0.49) | 0 (0.00) | 0 (0.00) | 5 (18.52) | 7 (4.61) | 45 (1.90) | 79 (1.11) |
| **Missing** | 944 (20.92) | 9 (20.93) | 8 (25.81) | <5  (<15.00) | 49 (32.24) | 7 (0.30) | 2241 (31.03) |

|  | **Heterosexual or Straight** | **Gay or Lesbian** | **Bisexual** | **Other** | **Prefer not to say** | **Missing** | **Total** |
| --- | --- | --- | --- | --- | --- | --- | --- |
| N= | 4513 | 43 | 31 | 27 | 152 | 2364 | 7130 |
| **Attraction** |  |  |  |  |  |  |  |
| **Entirely with opposite** | 3369 (74.65) | <5  (<2.50) | 7 (22.58) | 15 (55.56) | 83 (54.61) | 2191 (92.68) | 5666 (79.47) |
| **Mostly with opposite, but some experience with same** | 147 (3.26) | <5  (<7.50) | 11 (35.48) | <5  (<10.00) | <5  (<5.00) | 51 (2.16) | 218 (3.06) |
| **Equally with opposite and same** | 22 (0.49) | <5  (<7.50) | <5  (<10.00) | 0 (0.00) | <5  (<5.00) | 25 (1.06) | 57 (0.80) |
| **Mostly with same, but some experience with opposite** | <5  (<0.50) | <5  (<10.00) | <5  (<7.50) | 0 (0.00) | <5  (<2.50) | 10 (0.42) | 19 (0.27) |
| **Entirely with same** | <5  (<0.50) | 22 (51.16) | 0 (0.00) | <5  (<7.50) | 0 (0.00) | 19 (0.80) | 46 (0.65) |
| **No sexual experience in lifetime** | 21 (0.47) | 0 (0.00) | 0 (0.00) | <5  (<15.00) | 11 (7.24) | 51 (2.16) | 87 (1.22) |
| **Missing** | 949 (21.03) | 10 (23.26) | 8 (25.81) | <5  (<15.00) | 49 (32.24) | 17 (0.72) | 1037 (14.54) |

#### Table S4: Experience by sexual identity in older adults

#### Table S5: Sexual identity by attraction in older adults

|  | **Entirely with opposite** | **Mostly with opposite, but some experience with same** | **Equally with opposite and same** | **Mostly with same, but some experience with opposite** | **Entirely with same** | **No sexual experience in lifetime** | **Missing** | **Total** |
| --- | --- | --- | --- | --- | --- | --- | --- | --- |
| N= | 5666 | 218 | 57 | 19 | 46 | 87 | 1037 | 7130 |
| **Sexual Identity** |  |  |  |  |  |  |  |  |
| **Heterosexual or Straight** | 3369 (59.46) | 147 (67.43) | 22 (38.60) | <5  (<15.00) | <5  (<7.50) | 21 (24.14) | 949 (91.51) | 4513 (63.30) |
| **Gay or Lesbian** | <5  (<0.50) | <5  (<2.50) | <5  (<7.50) | <5  (<25.00) | 22 (47.83) | 0 (0.00) | 10 (0.96) | 43 (0.60) |
| **Bisexual** | 7 (0.12) | 11 (5.05) | <5  (<7.50) | <5  (<15.00) | 0 (0.00) | 0 (0.00) | 8 (0.77) | 31 (0.43) |
| **Other** | 15 (0.26) | <5  (<2.50) | 0 (0.00) | 0 (0.00) | <5  (<5.00) | <5  (<5.00) | <5  (<0.50) | 27 (0.38) |
| **Prefer not to say** | 83 (1.46) | <5  (<2.50) | <5  (<7.50) | <5  (<7.50) | 0 (0.00) | 11 (12.64) | 49 (4.73) | 152 (2.13) |
| **Missing** | 2191 (38.67) | 51 (23.39) | 25 (43.86) | 10 (52.63) | 19 (41.30) | 51 (58.62) | 17 (1.64) | 2364 (33.16) |

#### Table S6: Experience by attraction in older adults

|  | **Entirely with opposite** | **Mostly with opposite, but some experience with same** | **Equally with opposite and same** | **Mostly with same, but some experience with opposite** | **Entirely with same** | **No sexual experience in lifetime** | **Schedule not applicable** | **Total** |
| --- | --- | --- | --- | --- | --- | --- | --- | --- |
| N= | 5666 | 218 | 57 | 19 | 46 | 87 | 1037 | 7130 |
| **Sexual experience** |  |  |  |  |  |  |  |  |
| **Entirely with opposite** | 5561 (98.15) | 140 (64.22) | 36 (63.16) | <5  (<15.00) | <5  (<5.00) | 27 (31.03) | 22 (2.12) | 5790 (81.21) |
| **Mostly with opposite, but some experience with same** | 78 (1.38) | 72 (33.03) | 5 (8.77) | <5  (<7.50) | 0 (0.00) | <5  (<2.50) | 0 (0.00) | 158 (2.22) |
| **Equally with opposite and same** | <5  (<0.50) | <5  (<2.50) | 11 (19.30) | 3 (15.79) | <5  (<5.00) | <5  (<2.50) | 0 (0.00) | 23 (0.32) |
| **Mostly with same, but some experience with opposite** | 0 (0.00) | 0 (0.00) | <5  (<2.50) | 11 (57.89) | 13 (28.26) | <5  (<2.50) | <5  (<2.50) | 27 (0.38) |
| **Entirely with same** | <5  (<0.50) | 0 (0.00) | 0 (0.00) | <5  (<15.00) | 27 (58.70) | <5  (<2.50) | 0 (0.00) | 33 (0.46) |
| **No sexual experience in lifetime** | 14 (0.25) | <5  (<2.50) | <5  (<5.00) | 0 (0.00) | <5  (<5.00) | 54 (62.07) | 5 (0.48) | 79 (1.11) |
| **Not answered** | 8 (0.14) | 0 (0.00) | <5  (<5.00) | 0 (0.00) | 0 (0.00) | <5  (<2.50) | 1009 (97.30) | 1020 (14.31) |

|  | **Entirely with opposite** | **Mostly with opposite, but some experience with same** | **Equally with opposite and same** | **Mostly with same, but some experience with opposite** | **Entirely with same** | **No sexual experience in lifetime** | **Missing** | **Total** |
| --- | --- | --- | --- | --- | --- | --- | --- | --- |
| N= | 5790 | 158 | 23 | 27 | 33 | 79 | 1020 | 7130 |
| **Sexual Identity** |  |  |  |  |  |  |  |  |
| **Heterosexual or Straight** | 3443 (74.20) | 98 (71.53) | <5  (<7.50) | <5  (<5.00) | <5  (<20.00 | 22 (38.60) | 32 (37.21) | 4513 (63.30) |
| **Gay or Lesbian** | 0 (0.00) | <5  (<5.00) | <5  (<20.00) | 11 (55.00) | 16 (61.54) | 0 (0.00) | 0 (0.00) | 43 (0.60) |
| **Bisexual** | 8 (0.17) | 11 (8.03) | <5  (<20.00 | <5  (<5.00) | 0 (0.00) | 0 (0.00) | 0 (0.00) | 31 (0.43) |
| **Other** | 17 (0.37) | <5  (<2.50) | 0 (0.00) | 0 (0.00) | 0 (0.00) | 5 (8.77) | <5  (<2.50) | 27 (0.38) |
| **Prefer not to say** | 89 (1.92) | <5  (<2.50) | <5  (<20.00 | <5  (<5.00) | 0 (0.00) | 7 (12.28) | 9 (10.47) | 152 (2.13) |
| **Missing** | 2233 (38.57) | 40 (25.32) | 13 (56.52) | 13 (48.15) | 13 (39.39) | 45 (56.96) | 7 (0.69) | 2364 (33.16) |

#### Table S7: Sexual identity by experience in older adults

#### Table S8: Attraction by experience in older adults

|  | **Entirely with opposite** | **Mostly with opposite, but some experience with same** | **Equally with opposite and same** | **Mostly with same, but some experience with opposite** | **Entirely with same** | **No sexual experience in lifetime** | **Missing** | **Total** |
| --- | --- | --- | --- | --- | --- | --- | --- | --- |
| N= | 5790 | 158 | 23 | 27 | 33 | 79 | 1020 | 7130 |
| **Sexual Attraction** |  |  |  |  |  |  |  |  |
| **Entirely with opposite** | 5561 (96.04) | 78 (49.37) | <5  (<10.00) | 0 (0.00) | 3 (9.09) | 14 (17.72) | 8 (0.78) | 5666 (79.47) |
| **Mostly with opposite, but some experience with same** | 140 (2.42) | 72 (45.57) | <5  (<20.00) | 0 (0.00) | 0 (0.00) | <5  (<5.00) | 0 (0.00) | 218 (3.06) |
| **Equally with opposite and same** | 36 (0.62) | 5 (3.16) | 11 (47.83) | <5  (<5.00) | 0 (0.00) | <5  (<5.00) | <5  (<0.50) | 57 (0.80) |
| **Mostly with same, but some experience with opposite** | <5  (<0.50) | <5  (<2.50) | <5  (<15.00) | 11 (40.74) | <5  (<5.00) | 0 (0.00) | 0 (0.00) | 19 (0.27) |
| **Entirely with same** | <5  (<0.50) | 0 (0.00) | <5  (<5.00) | 13 (48.15) | 27 (81.82) | <5  (<5.00) | 0 (0.00) | 46 (0.65) |
| **No sexual experience in lifetime** | 27 (0.47) | <5  (<2.50) | <5  (<5.00) | <5  (<5.00) | <5  (<5.00) | 54 (68.35) | <5  (<0.50) | 87 (1.22) |
| **Missing** | 22 (0.38) | 0 (0.00) | 0 (0.00) | <5  (<5.00) | 0 (0.00) | 5 (6.33) | 1009 (98.92) | 1037 (14.54) |

### Supplementary Results

#### Table S9: Demographic characteristics by sexual identity in adolescents

|  | **Completely heterosexual** | **Mainly heterosexual** | **Bisexual** | **Mainly Gay or Lesbian** | **Completely Gay or Lesbian** | **Other** | **Do Not Know** | **Prefer Not to Say** | **Total** |
| --- | --- | --- | --- | --- | --- | --- | --- | --- | --- |
|  | N= 7,888 | N= 1,101 | N= 656 | N= 90 | N= 160 | N= 157 | N= 16 | N = 35 | N= 10,103 |
| **Gender Identity (Four category)** |  |  |  |  |  |  |  |  |  |
| Male | 4154 (52.66) | 399 (36.24) | 159 (24.24) | 33 (36.67) | 86 (53.75) | 40 (25.48) | 5 (31.25) | 8 (22.86) | 4884 (48.34) |
| Female | 3719 (47.15) | 691 (62.76) | 472 (71.95) | 47 (52.22) | 63 (39.38) | 74 (47.13) | 10 (62.50) | 15 (42.86) | 5091 (50.39) |
| Non-binary/Other | 0 (0.00) | <5 (<0.50) | 14 (2.13) | 7 (7.78) | 10 (6.25) | 25 (15.92) | 0 (0.00) | 0 (0.00) | 58 (0.57) |
| Don’t know / PNS | 9 (0.11) | 5 (0.45) | 11 (1.68) | <5 (<5.00) | <5 (<2.50) | 18 (11.46) | <5 (<7.50) | 11 (31.43) | 59 (0.58) |
| Missing | 6 (0.08) | <5 (<2.50) | 0 (0.00) | 0 (0.00) | 0 (0.00) | 0 (0.00) | 0 (0.00) | <5 (<5.00) | 11 (0.11 |
| **Ethnicity** |  |  |  |  |  |  |  |  |  |
| White | 6233 (79.02) | 929 (84.38) | 587 (89.48) | 81 (90.00) | 146 (91.25) | 129 (82.17) | 11 (68.75) | 24 (68.57) | 8,140 (80.59) |
| **OECD Equivalised income quintiles** |  |  |  |  |  |  |  |  |  |
| Lower quantile | 1325 (16.80) | 108 (9.81) | 83 (12.65) | 8 (8.89) | 25 (15.63) | 26 (16.56) | <5 (<25.00) | 12 (34.29) | 1,591 (15.75) |
| Second quantile | 1236 (15.67) | 145 (13.17) | 100 (15.24) | 12 (13.33) | 36 (22.50) | 31 (19.75) | 5 (31.25) | 9 (25.71) | 1,576 (15.60) |
| Third quantile | 1568 (19.88) | 211 (19.16) | 128 (19.51) | 15 (16.67) | 26 (16.25) | 30 (19.11) | 0 (0.00) | 5 (14.29) | 1,988 (19.68) |
| Fourth quantile | 1831 (23.21) | 271 (24.61) | 147 (22.41) | 26 (28.89) | 31 (19.38) | 31 (19.75) | <5 (<15.50) | 6 (17.14) | 2,346 (23.22) |
| Highest quantile | 1750 (22.19) | 341 (30.97) | 183 (27.90) | 28 (31.11) | 38 (23.75) | 38 (24.20) | <5 (<25.00) | <5 (<5.00) | 2,387 (23.63) |
| Missing | 178 (2.26) | 25 (2.27) | 15 (2.29) | <5 (<2.50) | <5 (<2.50) | <5 (<2.50) | <5 (<7.50) | <5 (<7.50) | 215 (2.13 |
| **Highest parent/guardian qualification (NVQ Level)** |  |  |  |  |  |  |  |  |  |
| NVQ Level 1 | 327 (4.15) | 21 (1.91) | 23 (3.51) | <5 (<2.50) | 6 (3.75) | <5 (<2.50) | <5 (<7.50) | <5 (<15.00) | 388 (3.84) |
| NVQ Level 2 | 1384 (17.55) | 164 (14.90) | 97 (14.79) | 18 (20.00) | 33 (20.63) | 27 (17.20) | <5 (<20.00) | 9 (25.71) | 1,742 (17.42) |
| NVQ Level 3 | 1135 (14.39) | 115 (10.45) | 81 (12.35) | 8 (8.89) | 27 (16.88) | 20 (12.74) | <5 (<25.00) | <5 (<7.50) | 1,392 (13.78) |
| NVQ Level 4 | 2863 (36.30) | 445 (40.42) | 257 (39.18) | 38 (42.22) | 49 (30.63) | 61 (38.85) | <5 (<20.00) | 7 (20.00) | 3,725 (36.87) |
| NVQ Level 5 | 1403 (17.79) | 286 (25.98) | 143 (21.80) | 22 (24.44) | 27 (16.88) | 30 (19.11) | <5 (<15.50) | 0 (0.00) | 1,913 (18.93) |
| None | 371 (4.70) | 27 (2.45) | 18 (2.74) | <5 (<2.50) | 10 (6.25) | 11 (7.01) | <5 (<15.50) | 5 (14.29) | 454 (4.49) |
| Overseas/Other | 166 (2.10) | 12 (1.09) | 16 (2.44) | 0 (0.00) | <5 (<2.50) | <5 (<2.50) | 0 (0.00) | <5 (<10.00) | 206 (2.04) |
| Missing | 239 (3.03) | 31 (2.82) | 21 (3.20) | <5 (<2.50) | <5 (<2.50) | <5 (<2.50) | <5 (<7.50) | 5 (14.29) | 283 (2.80) |
| **ONS Urban/Rural Classification 2005** |  |  |  |  |  |  |  |  |  |
| Rural | 1783 (22.60) | 298 (27.07) | 152 (23.17) | 14 (15.56) | 21 (13.13) | 39 (24.84) | 0 (0.00) | <5 (<10.00) | 2,310 (22.86) |
| Urban | 5841 (74.05) | 768 (69.75) | 483 (73.63) | 73 (81.11) | 135 (84.38) | 115 (73.25) | 15 (93.75) | 29 (82.86) | 7,459 (73.83) |
| Missing | 264 (3.35) | 35 (3.18) | 21 (3.20) | <5 (<5.00) | <5 (<2.50) | <5 (<2.50) | <5 (<7.50) | <5 (<10.00) | 334 (3.31) |

#### Table S10: Demographic characteristics by sexual attraction in adolescents

|  | **Only to opposite** | **More often to opposite** | **About equally** | **More often to same** | **Only to same** | **Never sexually attracted** | **Do not know** | **I do not wish to answer** | **Missing** | **Total** |
| --- | --- | --- | --- | --- | --- | --- | --- | --- | --- | --- |
| **N=10,345)** | N=7,656 | N=1,382 | N=385 | N=194 | N=137 | N=291 | N=11 | N=46 | N=1 | N=10,103 |
| **Gender Identity (Four category)** |  |  |  |  |  |  |  |  |  |  |
| Male | 4155 (54.27) | 399 (28.87) | 81 (21.04) | 67 (34.54) | 75 (54.74) | 96 (32.99) | 2 (18.18) | 9 (19.57) | - | 4884 (48.34) |
| Female | 3476 (45.40) | 969 (70.12) | 280 (72.73) | 108 (55.67) | 51 (37.23) | 174 (59.79) | 5 (45.45) | 28 (60.87) | - | 5091 (50.39) |
| Non-binary/Other | 2 (0.03) | 8 (0.58) | 15 (3.90) | 14 (7.22) | 8 (5.84) | 10 (3.44) | 0 (0.00) | 0 (0.00) | - | 58 (0.57) |
| Don’t know / PNS | 15 (0.20) | <5 (<0.50) | 9 (2.34) | 5 (2.58) | <5 (<2.50) | 11 (3.78) | <5 (<40.00) | 8 (17.39) | - | 59 (0.58) |
| Missing | 8 (0.10) | <5 (<0.50) | 0 (0.00) | 0 (0.00) | 0 (0.00) | 0 (0.00) | 0 (0.00) | <5 (<2.50) | - | 11 (0.11 |
| **Ethnicity** |  |  |  |  |  |  |  |  |  |  |
| White | 6072 (79.31) | 1204 (87.12) | 343 (89.09) | 167 (86.08) | 127 (92.70) | 190 (65.29) | 6 (54.55) | 30 (65.22) | - | 8,140 (80.59) |
| **OECD Equivalised income quintiles** |  |  |  |  |  |  |  |  |  |  |
| Lower quantile | 1265 (16.52) | 126 (9.12) | 52 (13.51) | 17 (8.76) | 25 (18.25) | 84 (28.87) | <5 (<40.00) | 18 (39.13) | - | 1,591 (15.75) |
| Second quantile | 1196 (15.62) | 191 (13.82) | 66 (17.14) | 25 (12.89) | 33 (24.09) | 53 (18.21) | <5 (<30.00) | 7 (15.22) | - | 1,576 (15.60) |
| Third quantile | 1512 (19.75) | 288 (20.84) | 69 (17.92) | 44 (22.68) | 18 (13.14) | 42 (14.43) | <5 (<10.00) | 9 (19.57) | - | 1,988 (19.68) |
| Fourth quantile | 1781 (23.26) | 355 (25.69) | 76 (19.74) | 48 (24.74) | 27 (19.71) | 49 (16.84) | <5 (<20.00) | 7 (15.22) | - | 2,346 (23.22) |
| Highest quantile | 1732 (22.62) | 391 (28.29) | 108 (28.05) | 58 (29.90) | 31 (22.63) | 57 (19.59) | <5 (<10.00) | <5 (<10.00) | - | 2,387 (23.63) |
| Missing | 170 (2.22) | 31 (2.24) | 14 (3.64) | 2 (1.03) | <5 (<2.50) | 6 (2.06) | 0 (0.00) | <5 (<2.50) | - | 215 (2.13 |
| **Highest parent/guardian qualification (NVQ Level)** |  |  |  |  |  |  |  |  |  |  |
| NVQ Level 1 | 305 (3.98) | 34 (2.46) | 12 (3.12) | 5 (2.58) | 5 (3.65) | 21 (7.22) | 2 (18.18) | <5 (<7.50) | - | 388 (3.84) |
| NVQ Level 2 | 1341 (17.52) | 208 (15.05) | 56 (14.55) | 36 (18.56) | 25 (18.25) | 55 (18.90) | <5 (<30.00) | 11 (23.91) | - | 1,742 (17.42) |
| NVQ Level 3 | 1091 (14.25) | 173 (12.52) | 45 (11.69) | 24 (12.37) | 19 (13.87) | 36 (12.37) | 0 (0.00) | 5 (<10.00) | - | 1,392 (13.78) |
| NVQ Level 4 | 2774 (36.23) | 561 (40.59) | 143 (37.14) | 79 (40.72) | 45 (32.85) | 107 (36.77) | <5 (<30.00) | 11 (23.91) | - | 3,725 (36.87) |
| NVQ Level 5 | 1394 (18.21) | 322 (23.30) | 90 (23.38) | 42 (21.65) | 26 (18.98) | 36 (12.37) | 0 (0.00) | <5 (<5.00) | - | 1,913 (18.93) |
| None | 353 (4.61) | 31 (2.24) | 14 (3.64) | <5 (<2.50) | 10 (7.30) | 23 (7.90) | <5 (<30.00) | 8 (17.39) | - | 454 (4.49) |
| Overseas/Other | 170 (2.22) | 9 (0.65) | 10 (2.60) | <5 (<2.50) | <5 (<5.00) | 5 (1.72) | 0 (0.00) | <5 (<7.50) | - | 206 (2.04) |
| Missing | 228 (2.98) | 44 (3.18) | 15 (3.90) | <5 (<2.50) | <5 (<2.50) | 8 (2.75) | 0 (0.00) | 5 (<10.00) | - | 283 (2.80) |
| **ONS Urban/Rural Classification 2005** |  |  |  |  |  |  |  |  |  |  |
| Rural | 1747 (22.82) | 372 (26.92) | 80 (20.78) | 32 (16.49) | 22 (16.06) | 51 (17.53) | <5 (<10.00) | 5 (<10.00) | - | 2,310 (22.86) |
| Urban | 5653 (73.84) | 967 (69.97) | 289 (75.06) | 158 (81.44) | 111 (81.02) | 231 (79.38) | 10 (90.91) | 40 (86.96) | - | 7,459 (73.83) |
| Missing | 256 (3.34) | 43 (3.11) | 16 (4.16) | <5 (<0.50) | 5 (<5.00) | 9 (3.09) | 0 (0.00) | <5 (<5.00) | - | 334 (3.31) |

#### Table S11: Demographic characteristics by sexual identity in older adults

|  | **Heterosexual or Straight** | **Gay or Lesbian** | **Bisexual** | **Other** | **Prefer Not to Say** | **Missing** | **Total** |
| --- | --- | --- | --- | --- | --- | --- | --- |
| N= | 4513 | 43 | 31 | 27 | 152 | 2364 | 7130 |
| **Sex** |  |  |  |  |  |  |  |
| Male | 2009 (44.52) | 26 (60.47) | 20 (64.52) | 12 (44.44) | 63 (41.45) | 1054 (44.59) | 3184 (44.66) |
| Female | 2504 (55.48) | 17 (39.53) | 11 (35.48) | 15 (55.56) | 89 (58.55) | 1310 (55.41) | 3946 (55.34) |
| **Age bands** |  |  |  |  |  |  |  |
| 50-59 | 584 (12.94) | 8 (18.60) | <5 (<15.00) | 0 (0.00) | 10 (6.58) | 310 (13.11) | 916 (12.85) |
| 60-69 | 2011 (44.56) | 19 (44.19) | 14 (45.16) | 9 (33.33) | 48 (31.58) | 723 (30.58) | 2824 (39.61) |
| 70-79 | 1387 (30.73) | 16 (37.21) | 13 (41.94) | 7 (25.93) | 52 (34.21) | 758 (32.06) | 2233 (31.32) |
| 80-89 | 490 (10.86) | 0 (0.00) | 0 (0.00) | 9 (33.33) | 40 (26.32 ) | 476 (20.14) | 1015 (14.24) |
| 90+ | 41 (0.91) | 0 (0.00) | 0 (0.00) | <5 (<7.50) | <5 (<2.50) | 97 (4.10) | 142 (1.99) |
| **Ethnicity** |  |  |  |  |  |  |  |
| White | 4410 (97.72) | 43 (100.00) | 31 (100.00) | 25 (92.59) | 135 (88.82) | 2293 (97.00) | 6937 (97.29) |
| Non-white | 103 (2.28) | 0 (0.00) | 0 (0.00) | <5 (<7.50) | 17 (11.18) | 71 (3.00) | 193 (2.71) |
| **Income quintile (BU net worth)** |  |  |  |  |  |  |  |
| Lower quantile | 665 (14.74) | 6 (13.95) | 6 (19.35) | 8 (29.63) | 39 (25.66) | 440 (18.61) | 1164 (16.33) |
| Second quantile | 835 (18.50) | 8 (18.60) | 8 (25.81) | 5 (18.52) | 35 (23.03) | 511 (21.62) | 1402 (19.66) |
| Third quantile | 938 (20.78) | 8 (18.60) | <5 (<15.00) | 9 (33.33) | 38 (25.00) | 478 (20.22) | 1475 (20.69) |
| Fourth quantile | 1016 (22.51) | 7 (16.28) | 7 (22.58) | 5 (18.52) | 25 (16.45) | 470 (19.88) | 1530 (21.46) |
| Highest quantile | 1037 (22.98) | 14 (32.56) | 6 (19.35) | 0 (0.00) | 15 (9.87) | 434 (18.36) | 1506 (21.12) |
| Missing | 22 (0.49) | 0 (0.00) | 0 (0.00) | 0 (0.00) | 0 (0.00) | 31 (1.31) | 53 (0.74) |
| **Income quintile (BU equiv income)** |  |  |  |  |  |  |  |
| Lower quantile | 776 (17.19) | 7 (16.28) | 7 (22.58) | 6 (22.22) | 44 (28.95) | 507 (21.45) | 1347 (18.89) |
| Second quantile | 851 (18.86) | 5 (11.63) | 5 (16.13) | <5 (<15.00) | 43 (28.29) | 529 (22.38) | 1437 (20.15) |
| Third quantile | 907 (20.10) | 5 (11.63) | <5 (<15.00) | 9 (33.33) | 27 (17.76) | 483 (20.43) | 1435 (20.13) |
| Fourth quantile | 982 (21.76) | 15 (34.88) | 7 (22.58) | 7 (25.93) | 22 (14.47) | 470 (19.88) | 1503 (21.08) |
| Highest quantile | 975 (21.60) | 11 (25.58) | 8 (25.81) | <5 (<5.00) | 16 (10.53) | 344 (14.55) | 1355 (19.00) |
| Missing | 22 (0.49) | 0 (0.00) | 0 (0.00) | 0 (0.00) | 0 (0.00) | 31 (1.31) | 53 (0.74) |
| **Highest qualification** |  |  |  |  |  |  |  |
| Missing | 80 (1.77) | 0 (0.00) | <5 (<7.50) | 0 (0.00) | <5 (<2.50) | 10 (0.42) | 93 (1.30) |
| nvq4/nvq5/degree or equiv | 860 (19.06) | 10 (23.26) | 7 (22.58) | <5 (<15.00) | 6 (3.95) | 343 (14.51) | 1230 (17.25) |
| higher ed below degree | 654 (14.49) | 6 (13.95) | 5 (16.13) | 5 (18.52) | 8 (5.26) | 317 (13.41) | 995 (13.96) |
| nvq3/gce a level equiv | 421 (9.33) | 7 (16.28) | <5 (<10.00) | <5 (<15.00) | 7 (4.61) | 199 (8.42) | 640 (8.98) |
| nvq2/gce o level equiv | 901 (19.96) | 6 (13.95) | 5 (16.13) | <5 (<15.00) | 23 (15.13) | 448 (18.95) | 1387 (19.45) |
| nvq1/cse other grade equiv | 148 (3.28) | <5 (<2.50) | 0 (0.00) | <5 (<5.00) | 5 (3.29) | 104 (4.40) | 259 (3.63) |
| foreign/other | 658 (14.58) | 7 (16.28) | 6 (19.35) | <5 (<15.00) | 22 (14.47) | 283 (11.97) | 979 (13.73) |
| no qualification | 791 (17.53) | 6 (13.95) | <5 (<10.00) | 7 (25.93) | 80 (52.63) | 660 (27.92) | 1547 (21.70) |
| **Government Office Region** |  |  |  |  |  |  |  |
| North East | 319 (7.07) | <5 (<5.00) | <5 (<5.00) | <5 (<5.00) | 8 (5.26) | 100 (4.23) | 431 (6.04) |
| North West | 503 (11.15) | <5 (<7.50) | <5 (<5.00) | <5 (<7.50) | 19 (12.50) | 283 (11.97) | 811 (11.37) |
| Yorkshire and the Humber | 442 (9.79) | <5 (<7.50) | <5 (<5.00) | <5 (<15.00) | 16 (10.53) | 292 (12.35) | 758 (10.63) |
| East Midlands | 500 (11.08) | <5 (<5.00) | 6 (19.35) | <5 (<15.00) | 25 (16.45) | 249 (10.53) | 785 (11.01) |
| West Midlands | 480 (10.64) | 5 (11.63) | 5 (16.13) | <5 (<15.00) | 19 (12.50) | 233 (9.86) | 745 (10.45) |
| East of England | 611 (13.54) | <5 (<10.00) | 7 (22.58) | <5 (<15.00) | 18 (11.84) | 309 (13.07) | 952 (13.35) |
| London | 341 (7.56) | 13 (30.23) | <5 (<5.00) | <5 (<15.00) | 12 (7.89) | 201 (8.50) | 572 (8.02) |
| South East | 772 (17.11) | 5 (11.63) | 6 (19.35) | <5 (<15.00) | 24 (15.79) | 414 (17.51) | 1225 (17.18) |
| South West | 531 (11.77) | 7 (16.28) | <5 (<10.00) | <5 (<7.50) | 11 (7.24) | 275 (11.63) | 829 (11.63) |
| Missing | 14 (0.31) | 0 (0.00) | 0 (0.00) | 0 (0.00) | 0 (0.00) | 8 (0.34) | 22 (0.31) |

#### Table S12: Demographic characteristics by sexual attraction in older adults

|  | **Entirely with opposite** | **Mostly w. opp., but some experience w. same** | **Equally with opposite and same** | **Mostly w. same, but some experience w. opp.** | **Entirely with same** | **No sexual experience in lifetime** | **Missing** | **Total** |
| --- | --- | --- | --- | --- | --- | --- | --- | --- |
| N= | 5666 | 218 | 57 | 19 | 46 | 87 | 1037 | 7130 |
| **Sex** |  |  |  |  |  |  |  |  |
| Male | 2586 (45.64) | 71 (32.57) | 15 (26.32) | 7 (36.84) | 28 (60.87) | 19 (21.84) | 458 (44.17) | 3184 (44.66) |
| Female | 3080 (54.36) | 147 (67.43) | 42 (73.68) | 12 (63.16) | 18 (39.13) | 68 (78.16) | 579 (55.83) | 3946 (55.34) |
| **Age bands** |  |  |  |  |  |  |  |  |
| 50-59 | 594 (10.48) | 43 (19.72) | <5 (<7.50) | <5 (<20.00) | 10 (21.74) | <5 (<5.00) | 260 (25.07) | 916 (12.85) |
| 60-69 | 2293 (40.47) | 101 (46.33) | 19 (33.33) | 8 (42.11) | 15 (32.61) | 20 (22.99) | 368 (35.49) | 2824 (39.61) |
| 70-79 | 1826 (32.23) | 60 (27.52) | 26 (45.61) | 6 (31.58) | 17 (36.96) | 31 (35.63) | 267 (25.75) | 2233 (31.32) |
| 80-89 | 832 (14.68) | 14 (6.42) | 9 (15.79) | <5 (<15.00) | <5 (<10.00) | 25 (28.74) | 129 (12.44) | 1015 (14.24) |
| 90+ | 121 (2.14) | 0 (0.00) | 0 (0.00) | 0 (0.00) | 0 (0.00) | 8 (9.20) | 13 (1.25) | 142 (1.99) |
| **Ethnicity** |  |  |  |  |  |  |  |  |
| White | 5536 (97.71) | 215 (98.62) | 55 (96.49) | 19 (100.00) | 46 (100.00) | 80 (91.95) | 986 (95.08) | 6937 (97.29) |
| Non-white | 130 (2.29) | <5 (<2.50) | <5 (<5.00) | 0 (0.00) | 0 (0.00) | 7 (8.05) | 51 (4.92) | 193 (2.71) |
| **Income quintile (BU net worth)** |  |  |  |  |  |  |  |  |
| Lower quantile | 864 (15.25) | 32 (14.68) | 12 (21.05) | <5 (<20.00) | 10 (21.74) | 30 (34.48) | 213 (20.54) | 1164 (16.33) |
| Second quantile | 1085 (19.15) | 43 (19.72) | 12 (21.05) | <5 (<15.00) | 5 (10.87) | 28 (32.18) | 227 (21.89) | 1402 (19.66) |
| Third quantile | 1187 (20.95) | 36 (16.51) | 19 (33.33) | <5 (<15.00) | 9 (19.57) | 19 (21.84) | 203 (19.58) | 1475 (20.69) |
| Fourth quantile | 1272 (22.45) | 51 (23.39) | 8 (14.04) | 6 (31.58) | 11 (23.91) | 8 (9.20) | 174 (16.78) | 1530 (21.46) |
| Highest quantile | 1217 (21.48) | 56 (25.69) | 6 (10.53) | 6 (31.58) | 11 (23.91) | <5 (<2.50) | 208 (20.06) | 1506 (21.12) |
| Missing | 41 (0.72) | 0 (0.00) | 0 (0.00) | 0 (0.00) | 0 (0.00) | 0 (0.00) | 12 (1.16) | 53 (0.74) |
| **Income quintile (BU equiv income)** |  |  |  |  |  |  |  |  |
| Lower quantile | 1030 (18.18) | 46 (21.10) | 17 (29.82) | <5 (<20.00) | 10 (21.74) | 34 (39.08) | 207 (19.96) | 1347 (18.89) |
| Second quantile | 1163 (20.53) | 31 (14.22) | 15 (26.32) | <5 (<20.00) | 7 (15.22) | 24 (27.59) | 194 (18.71) | 1437 (20.15) |
| Third quantile | 1155 (20.38) | 35 (16.06) | 10 (17.54) | <5 (<15.00) | 5 (10.87) | 17 (19.54) | 211 (20.35) | 1435 (20.13) |
| Fourth quantile | 1219 (21.51) | 44 (20.18) | 7 (12.28) | 5 (26.32) | 13 (28.26) | 8 (9.20) | 207 (19.96) | 1503 (21.08) |
| Highest quantile | 1058 (18.67) | 62 (28.44) | 8 (14.04) | 6 (31.58) | 11 (23.91) | <5 (<5.00) | 206 (19.86) | 1355 (19.00) |
| Missing | 41 (0.72) | 0 (0.00) | 0 (0.00) | 0 (0.00) | 0 (0.00) | 0 (0.00) | 12 (1.16) | 53 (0.74) |
| **Highest qualification** |  |  |  |  |  |  |  |  |
| Missing | 18 (0.32) | <5 (<0.50) | 0 (0.00) | <5 (<7.50) | 0 (0.00) | 0 (0.00) | 73 (7.04) | 93 (1.30) |
| nvq4/nvq5/degree or equiv | 998 (17.61) | 53 (24.31) | <5 (<7.50) | 8 (42.11) | 9 (19.57) | 8 (9.20) | 151 (14.56) | 1230 (17.25) |
| higher ed below degree | 818 (14.44) | 28 (12.84) | 13 (22.81) | <5 (<15.00) | 5 (10.87) | 9 (10.34) | 120 (11.57) | 995 (13.96) |
| nvq3/gce a level equiv | 504 (8.90) | 33 (15.14) | <5 (<7.50) | <5 (<7.50) | 6 (13.04) | <5 (<5.00) | 88 (8.49) | 640 (8.98) |
| nvq2/gce o level equiv | 1124 (19.84) | 39 (17.89) | 9 (15.79) | <5 (<7.50) | 7 (15.22) | 10 (11.49) | 197 (19.00) | 1387 (19.45) |
| nvq1/cse other grade equiv | 215 (3.79) | 8 (3.67) | <5 (<7.50) | 0 (0.00) | <5 (<5.00) | <5 (<5.00) | 28 (2.70) | 259 (3.63) |
| foreign/other | 783 (13.82) | 35 (16.06) | 8 (14.04) | <5 (<20.00) | 7 (15.22) | 9 (10.34) | 134 (12.92) | 979 (13.73) |
| no qualification | 1206 (21.28) | 21 (9.63) | 17 (29.82) | <5 (<20.00) | 10 (21.74) | 44 (50.57) | 246 (23.72) | 1547 (21.70) |
| **Government Office Region** |  |  |  |  |  |  |  |  |
| North East | 314 (5.54) | 11 (5.05) | <5 (<7.50) | 0 (0.00) | <5 (<5.00) | 5 (5.75) | 96 (9.26) | 431 (6.04) |
| North West | 634 (11.19) | 16 (7.34) | <5 (<7.50) | <5 (<25.00) | <5 (<10.00) | 9 (10.34) | 140 (13.50) | 811 (11.37) |
| Yorkshire and the Humber | 615 (10.85) | 24 (11.01) | 10 (17.54) | <5 (<7.50) | 6 (13.04) | 10 (11.49) | 92 (8.87) | 758 (10.63) |
| East Midlands | 631 (11.14) | 19 (8.72) | 6 (10.53) | 0 (0.00) | <5 (<10.00) | 11 (12.64) | 114 (10.99) | 785 (11.01) |
| West Midlands | 549 (9.69) | 28 (12.84) | 6 (10.53) | <5 (<15.00) | <5 (<10.00) | 9 (10.34) | 147 (14.18) | 745 (10.45) |
| East of England | 814 (14.37) | 28 (12.84) | 8 (14.04) | <5 (<25.00) | <5 (<5.00) | 10 (11.49) | 86 (8.29) | 952 (13.35) |
| London | 440 (7.77) | 24 (11.01) | 6 (10.53) | <5 (<15.00) | 11 (23.91) | 11 (12.64) | 78 (7.52) | 572 (8.02) |
| South East | 1011 (17.84) | 41 (18.81) | 8 (14.04) | <5 (<25.00) | <5 (<10.00) | 16 (18.39) | 141 (13.60) | 1225 (17.18) |
| South West | 640 (11.30) | 27 (12.39) | 6 (10.53) | <5 (<15.00) | 9 (19.57) | 6 (6.90) | 139 (13.40) | 829 (11.63) |
| Missing | 18 (0.32) | 0 (0.00) | 0 (0.00) | 0 (0.00) | 0 (0.00) | 0 (0.00) | <5 (<0.50) | 22 (0.31) |

#### Table S13: Demographic characteristics by sexual experience in older adults

|  | **Entirely with opposite** | **Mostly w. opp., but some experience w. same** | **Equally with opposite and same** | **Mostly w. same, but some experience w. opp.** | **Entirely with same** | **No sexual experience in lifetime** | **Missing** | **Total** |
| --- | --- | --- | --- | --- | --- | --- | --- | --- |
| N= | 5790 | 158 | 23 | 27 | 33 | 79 | 1020 | 7130 |
| **Sex** |  |  |  |  |  |  |  |  |
| Male | 2574 (44.46) | 85 (53.80) | 6 (26.09) | 13 (48.15) | 20 (60.61) | 27 (34.18) | 459 (45.00) | 3184 (44.66) |
| Female | 3216 (55.54) | 73 (46.20) | 17 (73.91) | 14 (51.85) | 13 (39.39) | 52 (65.82) | 561 (55.00) | 3946 (55.34) |
| **Age bands** |  |  |  |  |  |  |  |  |
| 50-59 | 616 (10.64) | 24 (15.19) | <5 (<10.00) | 6 (22.22) | 8 (24.24) | <5 (<5.00) | 258 (25.29) | 916 (12.85) |
| 60-69 | 2319 (40.05) | 85 (53.80) | 9 (39.13) | 11 (40.74) | 11 (33.33) | 20 (25.32) | 369 (36.18) | 2824 (39.61) |
| 70-79 | 1878 (32.44) | 38 (24.05) | 8 (34.78) | 9 (33.33) | 12 (36.36) | 28 (35.44) | 260 (25.49) | 2233 (31.32) |
| 80-89 | 854 (14.75) | 11 (6.96) | <5 (<20.00) | <5 (<5.00) | <5 (<7.50) | 21 (26.58) | 122 (11.96) | 1015 (14.24) |
| 90+ | 123 (2.12) | 0 (0.00) | 0 (0.00) | 0 (0.00) | 0 (0.00) | 8 (10.13) | 11 (1.08) | 142 (1.99) |
| **Ethnicity** |  |  |  |  |  |  |  |  |
| White | 5658 (97.72) | 152 (96.20) | 22 (95.65) | 27 (100.00) | 33 (100.00) | 75 (94.94) | 970 (95.10) | 6937 (97.29) |
| Non-white | 132 (2.28) | 6 (3.80) | <5 (<5.00) | 0 (0.00) | 0 (0.00) | <5 (<7.50) | 50 (4.90) | 193 (2.71) |
| **Income quintile (BU net worth)** |  |  |  |  |  |  |  |  |
| Lower quantile | 887 (15.32) | 28 (17.72) | <5 (<15.00) | <5 (<15.00) | 5 (15.15) | 36 (45.57) | 201 (19.71) | 1164 (16.33) |
| Second quantile | 1127 (19.46) | 19 (12.03) | <5 (<15.00) | 5 (18.52) | 5 (15.15) | 17 (21.52) | 225 (22.06) | 1402 (19.66) |
| Third quantile | 1204 (20.79) | 33 (20.89) | 6 (26.09) | 5 (18.52) | <5 (<15.00) | 18 (22.78) | 205 (20.10) | 1475 (20.69) |
| Fourth quantile | 1292 (22.31) | 40 (25.32) | 6 (26.09) | 6 (22.22) | 9 (27.27) | 6 (7.59) | 171 (16.76) | 1530 (21.46) |
| Highest quantile | 1240 (21.42) | 37 (23.42) | <5 (<20.00) | 7 (25.93) | 10 (30.30) | <5 (<5.00) | 206 (20.20) | 1506 (21.12) |
| Missing | 40 (0.69) | <5 (<2.50) | 0 (0.00) | 0 (0.00) | 0 (0.00) | 0 (0.00) | 12 (1.18) | 53 (0.74) |
| **Income quintile (BU equiv income)** |  |  |  |  |  |  |  |  |
| Lower quantile | 1070 (18.48) | 33 (20.89) | 6 (26.09) | 5 (18.52) | 7 (21.21) | 29 (36.71) | 197 (19.31) | 1347 (18.89) |
| Second quantile | 1188 (20.52) | 24 (15.19) | 6 (26.09) | <5 (<15.00) | 5 (15.15) | 18 (22.78) | 193 (18.92) | 1437 (20.15) |
| Third quantile | 1173 (20.26) | 26 (16.46) | <5 (<20.00) | <5 (<15.00) | <5 (<7.50) | 20 (25.32) | 206 (20.20) | 1435 (20.13) |
| Fourth quantile | 1239 (21.40) | 30 (18.99) | <5 (<10.00) | 7 (25.93) | 9 (27.27) | 8 (10.13) | 208 (20.39) | 1503 (21.08) |
| Highest quantile | 1080 (18.65) | 44 (27.85) | 5 (21.74) | 8 (29.63) | 10 (30.30) | <5 (<7.50) | 204 (20.00) | 1355 (19.00) |
| Missing | 40 (0.69) | <5 (<2.50) | 0 (0.00) | 0 (0.00) | 0 (0.00) | 0 (0.00) | 12 (1.18) | 53 (0.74) |
| **Highest qualification** |  |  |  |  |  |  |  |  |
| Missing | 18 (0.31) | <5 (<2.50) | 0 (0.00) | <5 (<5.00) | 0 (0.00) | 0 (0.00) | 73 (7.16) | 93 (1.30) |
| nvq4/nvq5/degree or equiv | 1001 (17.29) | 49 (31.01) | <5 (<15.00) | 10 (37.04) | 8 (24.24) | 10 (12.66) | 149 (14.61) | 1230 (17.25) |
| higher ed below degree | 833 (14.39) | 23 (14.56) | 5 (21.74) | 5 (18.52) | <5 (<5.00) | 7 (8.86) | 121 (11.86) | 995 (13.96) |
| nvq3/gce a level equiv | 516 (8.91) | 26 (16.46) | <5 (<10.00) | <5 (<7.50) | <5 (<10.00) | <5 (<5.00) | 88 (8.63) | 640 (8.98) |
| nvq2/gce o level equiv | 1148 (19.83) | 27 (17.09) | <5 (<15.00) | <5 (<5.00) | 7 (21.21) | 7 (8.86) | 194 (19.02) | 1387 (19.45) |
| nvq1/cse other grade equiv | 227 (3.92) | 0 (0.00) | 0 (0.00) | 0 (0.00) | <5 (<7.50) | <5 (<5.00) | 28 (2.75) | 259 (3.63) |
| foreign/other | 813 (14.04) | 19 (12.03) | <5 (<15.00) | <5 (<15.00) | 5 (15.15) | 5 (6.33) | 130 (12.75) | 979 (13.73) |
| no qualification | 1234 (21.31) | 13 (8.23) | 7 (30.43) | <5 (<15.00) | 7 (21.21) | 45 (56.96) | 237 (23.24) | 1547 (21.70) |
| **Government Office Region** |  |  |  |  |  |  |  |  |
| North East | 322 (5.56) | 5 (3.16) | <5 (<5.00) | 0 (0.00) | <5 (<7.50) | 7 (8.86) | 94 (9.22) | 431 (6.04) |
| North West | 639 (11.04) | 16 (10.13) | <5 (<15.00) | <5 (<7.50) | <5 (<15.00) | 10 (12.66) | 137 (13.43) | 811 (11.37) |
| Yorkshire and the Humber | 635 (10.97) | 15 (9.49) | <5 (<20.00) | 5 (18.52) | 0 (0.00) | 11 (13.92) | 88 (8.63) | 758 (10.63) |
| East Midlands | 646 (11.16) | 12 (7.59) | 0 (0.00) | 0 (0.00) | <5 (<10.00) | 8 (10.13) | 116 (11.37) | 785 (11.01) |
| West Midlands | 565 (9.76) | 17 (10.76) | <5 (<10.00) | <5 (<7.50) | <5 (<15.00) | 8 (10.13) | 147 (14.41) | 745 (10.45) |
| East of England | 830 (14.34) | 24 (15.19) | <5 (<10.00) | <5 (<15.00) | <5 (<7.50) | 7 (8.86) | 84 (8.24) | 952 (13.35) |
| London | 449 (7.75) | 21 (13.29) | <5 (<10.00) | 8 (29.63) | 6 (18.18) | 11 (13.92) | 75 (7.35) | 572 (8.02) |
| South East | 1032 (17.82) | 30 (18.99) | 5 (21.74) | <5 (<15.00) | 6 (18.18) | 11 (13.92) | 138 (13.53) | 1225 (17.18) |
| South West | 654 (11.30) | 18 (11.39) | <5 (<20.00) | <5 (<15.00) | 6 (18.18) | 6 (7.59) | 137 (13.43) | 829 (11.63) |
| Missing | 18 (0.31) | 0 (0.00) | 0 (0.00) | 0 (0.00) | 0 (0.00) | 0 (0.00) | <5 (<0.50) | 22 (0.31) |

#### Table S14: Health outcomes by sexual identity in adolescents

|  | **Completely heterosexual** | **Mainly heterosexual** | **Bisexual** | **Mainly Gay or Lesbian** | **Completely Gay or Lesbian** | **Other** | **Do Not Know** | **Prefer Not to Say** | **Total** |
| --- | --- | --- | --- | --- | --- | --- | --- | --- | --- |
|  | N= 7,888 | N= 1,101 | N= 656 | N= 90 | N= 160 | N= 157 | N= 16 | N = 35 | N= 10,103 |
| **On the whole, I am satisfied with myself** |  |  |  |  |  |  |  |  |  |
| Strongly agree | 1787  (22.65) | 124  (11.26) | 51  (7.77) | 7  (7.78) | 29  (18.13) | 14  (8.92) | 0  (0) | 8  (22.86) | 2020  (19.99) |
| Agree | 4506  (57.12) | 579  (52.59) | 301  (45.88) | 43  (47.78) | 59  (36.88) | 65  (41.40) | 11  (68.75) | 14  (40.00) | 5578  (55.21) |
| Disagree | 1181  (14.97) | 304  (27.61) | 227  (34.60) | 24  (26.67) | 50  (31.25) | 42  (26.75) | <5 (<20.00) | 5  (14.29) | 1836  (18.17) |
| Strongly disagree | 401  (5.08) | 92  (8.36) | 76  (11.59) | 16  (17.78) | 22  (13.75) | 33  (21.02) | <5  (<7.50) | <5  (<7.50) | 643  (6.36) |
| Do Not Know | 5  (0.06) | <5  (<0.50) | <5  (<0.50) | 0  (0) | 0  (0) | <5  (<2.50) | <5  (<7.50) | <5  (<7.50) | 14  (0.14) |
| I do not wish to answer | 8  (0.10) | 0  (0) | 0  (0) | 0  (0) | 0  (0) | 0  (0) | 0  (0) | <5  (<15.00) | 12  (0.12) |
| **How would you describe your health generally?** |  |  |  |  |  |  |  |  |  |
| Excellent | 2395  (30.36) | 265  (24.07) | 118  (17.99) | 15  (16.67) | 34  (21.25) | 27  (17.20) | <5  (<7.50) | 11  (31.43) | 2866  (28.37) |
| Very good | 3,124  (39.60) | 427  (38.78) | 237  (36.13) | 36  (40.00) | 5  (34.38) | 47  (29.94) | <5  (<15.00) | 9  (25.71) | 3937  (38.97) |
| Good | 1822  (23.10) | 293  (26.61) | 215  (32.77) | 26  (28.89) | 42  (26.25) | 59  (37.58) | 7  (43.75) | 10  (28.57) | 2474  (24.49) |
| Fair | 391  (4.96) | 85  (7.72) | 58  (8.84) | 11  (12.22) | 19  (11.88) | 18  (11.46) | 5  (31.25) | <5  (<10.00) | 590  (5.84) |
| Poor | 77  (0.95) | 17  (1.54) | 20  (3.05) | <5  (<2.50) | <5  (<2.50) | 5  (3.18) | 0  (0) | 0  (0) | 122  (1.21) |
| No answer | 81  (1.03) | 14  (1.27) | 8  (1.22) | 0  (0.00) | 7  (4.38) | <5  (<2.50) | <5  (<7.50) | <5  (<7.50) | 114  (1.13) |
| **KESSLER** |  |  |  |  |  |  |  |  |  |
|  | N=7880 | N=1100 | N=656 | N=90 | N=160 | N=157 | N=16 | N=28 | N=10,092 |
| Mean KESSLER (95% CI) | 6.50  (6.40, 6.60) | 9.36  (9.08, 9.64) | 11.18  (10.79, 11.56) | 10.79  (9.73, 11.85) | 10.09  (9.17, 11.01) | 10.78  (9.80, 11.76) | 7.75  (5.23, 10.27) | 5.36  (3.49, 7.22) | 7.28  (7.18, 7.37) |

Table S15: Health outcomes by sexual attraction in adolescents

|  | **Only to opposite** | **More often to opposite** | **About equally** | **More often to same** | **Only to same** | **Never sexually attracted** | **Do not know** | **I do not wish to answer** | **No answer** | **Total** |
| --- | --- | --- | --- | --- | --- | --- | --- | --- | --- | --- |
|  | N=7,656 | N=1,382 | N=385 | N=194 | N=137 | N=291 | N=11 | N=46 | N=1 | N=10,103 |
| **On the whole, I am satisfied with myself** | | | | | | | | | |  |
| Strongly agree | 1734 (22.65) | 150 (10.85) | 24 (6.23) | 14 (7.22) | 26 (18.98) | 57 (19.59) | <5 (<30.00) | 12 (26.09) | - | 2020 (19.99) |
| Agree | 4412 (57.63) | 664 (48.05) | 180 (46.75) | 92 (47.42) | 54 (39.42) | 155 (53.26) | <5 (<30.00) | 18 (39.13) | - | 5580 (55.21) |
| Disagree | 1127 (14.72) | 422 (30.54) | 130 (33.77) | 53 (27.32) | 40 (29.20) | 54 (18.56) | <5 (<30.00) | 7 (15.22) | - | 1836 (18.17) |
| Strongly disagree | 373 (4.87) | 143 (10.35) | 50 (12.99) | 35 (18.04) | 17 (12.41) | 21 (7.22) | 0 (0.00) | <5 (<7.50) | - | 643 (6.36) |
| Do Not Know | 5 (0.07) | <5 (<0.50) | <5 (<0.50) | 0 (0.00) | 0 (0.00) | <5 (<2.50) | <5 (<20.00) | <5 (<2.50) | - | 14 (0.14) |
| I do not wish to answer | 5 (0.07) | <5 (<0.50) | 0 (0.00) | 0 (0.00) | 0 (0.00) | <5 (<0.50) | 0 (0.00) | 5 (10.87) | - | 12 (0.12) |
| **How would you describe your health generally?** | | | | | | | | | |  |
| Excellent | 2340 (30.69) | 305 (22.07) | 68 (17.66) | 29 (14.95) | 33 (24.09) | 68 (23.37) | <5 (<2.50) | 12 (26.09) | - | 2,866 (28.37) |
| Very good | 3047 (39.80) | 509 (36.83) | 141 (36.62) | 78 (40.21) | 48 (33.04) | 97 (33.33) | <5 (<40.00) | 13 (28.26) | - | 3,937 (38.97) |
| Good | 1747 (22.82) | 395 (28.52) | 136 (35.32) | 58 (29.90) | 34 (24.82) | 86 (29.55) | <5 (<40.00) | 14 (30.43) | - | 2,474 (24.49) |
| Fair | 363 (4.74) | 126 (9.12) | 24 (6.23) | 21 (10.82) | 15 (10.95) | 32 (11.00) | <5 (<20.00) | 6 (13.04) | - | 590 (5.94) |
| Poor | 73 (0.95) | 27 (1.95) | 8 (2.08) | 5 (2.58) | <5 (<2.50) | 6 (2.06) | 0 (0.00) | <5 (<2.50) | - | 122 (1.21) |
| No answer | 76 (0.99) | 20 (1.45) | 8 (2.08) | <5 (<2.50) | 5 (3.65) | <5 (<2.50) | 0 (0.00) | 0 (0.00) | - | 144 (1.13 |
| **KESSLER** | | | | | | | | | |  |
|  | N=7652 | N=1382 | N=385 | N=194 | N=137 | N=290 | N=11 | N=35 | - | N=10092 |
| Mean KESSLER (95% CI) | 6.43 (6.33, 6.53) | 10.03 (9.78, 10.29) | 11.54 (11.03, 12.04) | 11.06 (10.32, 11.80) | 9.77 (8.79, 10.76) | 7.27 (6.63, 7.90) | 6.18 (2.51, 9.85) | 5.60 (3.91, 7.29) | - | 7.28 (7.18, 7.37) |

Table S16: Health outcomes by sexual identity in older adults

|  | **Heterosexual or Straight** | **Gay or Lesbian** | **Bisexual** | **Other** | **Prefer Not to Say** | **Missing** | **Total** |
| --- | --- | --- | --- | --- | --- | --- | --- |
| N= | 4513 | 43 | 31 | 27 | 152 | 2364 | 7130 |
| **Life Satisfaction** |  |  |  |  |  |  |  |
| Missing | 9 (0.20) | 0 (0.00) | 0 (0.00) | 0 (0.00) | 0 (0.00) | 46 (1.95) | 55 (0.77) |
| Strongly agree | 693 (15.36) | 7 (16.28) | 6 (19.35) | 8 (29.63) | 26 (17.11) | 329 (13.92) | 1069 (14.99) |
| Agree | 2221 (49.21) | 21 (48.84) | 13 (41.94) | 12 (44.44) | 70 (46.05) | 1004 (42.47) | 3341 (46.86) |
| Slightly agree | 710 (15.73) | <5 (<10.00) | 5 (16.13) | <5 (<15.00) | 21 (13.82) | 407 (17.22) | 1150 (16.13) |
| Neither agree nor disagree | 353 (7.82) | <5 (<10.00) | <5 (<15.00) | 0 (0.00) | 20 (13.16) | 211 (8.93) | 591 (8.29) |
| Slightly disagree | 283 (6.27) | <5 (<5.00) | <5 (<10.00) | <5 (<15.00) | 8 (5.26) | 159 (6.73) | 458 (6.42) |
| Disagree | 188 (4.17) | <5 (<10.00) | 0 (0.00) | <5 (<5.00) | 7 (4.61) | 135 (5.71) | 335 (4.70) |
| Strongly disagree | 56 (1.24) | <5 (<5.00) | 0 (0.00) | 0 (0.00) | 0 (0.00) | 73 (3.09) | 131 (1.84) |
| **Self-rated General Health** |  |  |  |  |  |  |  |
| Missing | 0 (0.00) | 0 (0.00) | 0 (0.00) | 0 (0.00) | 0 (0.00) | <5 (<0.50) | <5 (<0.50) |
| excellent | 566 (12.54) | 5 (11.63) | 6 (19.35) | <5 (<5.00) | 8 (5.26) | 187 (7.91) | 774 (10.86) |
| very good | 1362 (30.18) | 11 (25.58) | 6 (19.35) | 8 (29.63) | 30 (19.74) | 581 (24.58) | 1998 (28.02) |
| Good | 1487 (32.95) | 13 (30.23) | 12 (38.71) | 6 (22.22) | 48 (31.58) | 778 (32.91) | 2344 (32.88) |
| Fair | 797 (17.66) | 7 (16.28) | <5 (<10.00) | 7 (25.93) | 50 (32.89) | 522 (22.08) | 1386 (19.44) |
| Poor | 301 (6.67) | 7 (16.28) | 4 (12.90) | <5 (<15.00) | 16 (10.53) | 295 (12.48) | 627 (8.79) |
| **CES-D** |  |  |  |  |  |  |  |
| N= | 4513 | 43 | 31 | 27 | 152 | 2,357 | 7130 |
| **Mean CES-D (95% CI)** | 2.91 (2.87, 2.95) | 3.07 (2.65, 3.49) | 2.87 (2.45, 3.29) | 3.56 (2.96, 4.15) | 3.24 (3.01, 3.47) | 3.13 (3.08, 3.19) | 2.99 (2.96, 3.03) |

Table S17: Health outcomes by sexual attraction in older adults

|  | **Entirely with opposite** | **Mostly w. opp., but some experience w. same** | **Equally with opposite and same** | **Mostly w. same, but some experience w. opp.** | **Entirely with same** | **No sexual experience in lifetime** | **Missing** | **Total** |
| --- | --- | --- | --- | --- | --- | --- | --- | --- |
| N= | 5666 | 218 | 57 | 19 | 46 | 87 | 1037 | 7130 |
| **Life Satisfaction** |  |  |  |  |  |  |  |  |
| Missing | 43 (0.76) | 0 (0.00) | 0 (0.00) | 0 (0.00) | <5 (<2.50) | 1 (1.15) | 10 (0.96) | 55 (0.77) |
| Strongly agree | 849 (14.98) | 26 (11.93) | 11 (19.30) | 5 (26.32) | 11 (23.91) | 17 (19.54) | 150 (14.46) | 1069 (14.99) |
| Agree | 2640 (46.59) | 108 (49.54) | 28 (49.12) | 9 (47.37) | 21 (45.65) | 38 (43.68) | 497 (47.93) | 3341 (46.86) |
| Slightly agree | 931 (16.43) | 27 (12.39) | 5 (8.77) | <5 (<7.50) | <5 (<7.50) | 13 (14.94) | 170 (16.39) | 1150 (16.13) |
| Neither agree nor disagree | 478 (8.44) | 21 (9.63) | <5 (<7.50) | 0 (0.00) | <5 (<7.50) | <5 (<5.00) | 82 (7.91) | 591 (8.29) |
| Slightly disagree | 357 (6.30) | 21 (9.63) | 6 (10.53) | <5 (<7.50) | <5 (<5.00) | 8 (9.20) | 63 (6.08) | 458 (6.42) |
| Disagree | 264 (4.66) | 10 (4.59) | <5 (<7.50) | <5 (<20.00) | <5 (<5.00) | <5 (<5.00) | 50 (4.82) | 335 (4.70) |
| Strongly disagree | 104 (1.84) | 5 (2.29) | <5 (<2.50) | 0 (0.00) | <5 (<7.50) | <5 (<10.00) | 15 (1.45) | 131 (1.84) |
| **Self-rated General Health** |  |  |  |  |  |  |  |  |
| Missing | 0 (0.00) | <5 (<0.50) | 0 (0.00) | 0 (0.00) | 0 (0.00) | 0 (0.00) | 0 (0.00) | <5 (<0.50) |
| excellent | 603 (10.64) | 26 (11.93) | 6 (10.53) | 0 (0.00) | 9 (19.57) | 9 (10.34) | 121 (11.67) | 774 (10.86) |
| very good | 1605 (28.33) | 71 (32.57) | 9 (15.79) | 6 (31.58) | 10 (21.74) | 9 (10.34) | 288 (27.77) | 1998 (28.02) |
| Good | 1857 (32.77) | 72 (33.03) | 20 (35.09) | 10 (52.63) | 11 (23.91) | 25 (28.74) | 349 (33.65) | 2344 (32.88) |
| Fair | 1118 (19.73) | 35 (16.06) | 16 (28.07) | <5 (<15.00) | 8 (17.39) | 20 (22.99) | 187 (18.03) | 1386 (19.44) |
| Poor | 483 (8.52) | 13 (5.96) | 6 (10.53) | <5 (<7.50) | 8 (17.39) | 24 (27.59) | 92 (8.87) | 627 (8.79) |
| **CES-D** |  |  |  |  |  |  |  |  |
| N= | 5,659 | 218 | 57 | 19 | 46 | 87 | 1037 | 7,123 |
| **Mean CES-D (95% CI)** | 2.98 (2.94, 3.01) | 2.99 (2.81, 3.17) | 3.07 (2.70, 3.44) | 2.95 (2.40, 3.50) | 3.02 (2.61, 3.43) | 3.59 (3.24, 3.93) | 3.05 (2.96, 3.13) | 2.99 (2.96, 3.03) |

Table S18: Health outcomes by sexual experience in older adults

|  | **Entirely with opposite** | **Mostly w. opp., but some experience w. same** | **Equally with opposite and same** | **Mostly w. same, but some experience w. opp.** | **Entirely with same** | **No sexual experience in lifetime** | **Not answered** | **Total** |
| --- | --- | --- | --- | --- | --- | --- | --- | --- |
| N= | 5790 | 158 | 23 | 27 | 33 | 79 | 1020 | 7130 |
| **Life Satisfaction** |  |  |  |  |  |  |  |  |
| Missing | 41 (0.71) | 0 (0.00) | <5 (<10.00) | 0 (0.00) | <5 (<7.50) | <5 (<5.00) | 8 (0.78) | 55 (0.77) |
| Strongly agree | 870 (15.03) | 18 (11.39) | <5 (<15.00) | 6 (22.22) | 6 (18.18) | 21 (26.58) | 145 (14.22) | 1069 (14.99) |
| Agree | 2702 (46.67) | 83 (52.53) | 11 (47.83) | 13 (48.15) | 17 (51.52) | 25 (31.65) | 490 (48.04) | 3341 (46.86) |
| Slightly agree | 944 (16.30) | 22 (13.92) | <5 (<10.00) | <5 (<10.00) | <5 (<7.50) | 9 (11.39) | 169 (16.57) | 1150 (16.13) |
| Neither agree nor disagree | 488 (8.43) | 8 (5.06) | <5 (<10.00) | 0 (0.00) | <5 (<7.50) | 10 (12.66) | 81 (7.94) | 591 (8.29) |
| Slightly disagree | 375 (6.48) | 10 (6.33) | <5 (<15.00) | <5 (<5.00) | 0 (0.00) | 7 (8.86) | 62 (6.08) | 458 (6.42) |
| Disagree | 269 (4.65) | 9 (5.70) | 0 (0.00) | 5 (18.52) | <5 (<5.00) | <5 (<2.50) | 50 (4.90) | 335 (4.70) |
| Strongly disagree | 101 (1.74) | 8 (5.06) | 0 (0.00) | 0 (0.00) | <5 (<10.00) | <5 (<7.50) | 15 (1.47) | 131 (1.84) |
| **Self-rated General Health** |  |  |  |  |  |  |  |  |
| Missing | 0 (0.00) | <5 (<2.50) | 0 (0.00) | 0 (0.00) | 0 (0.00) | 0 (0.00) | 0 (0.00) | <5 (<0.50) |
| excellent | 610 (10.54) | 24 (15.19) | <5 (<5.00) | <5 (<15.00) | 7 (21.21) | 8 (10.13) | 121 (11.86) | 774 (10.86) |
| very good | 1643 (28.38) | 39 (24.68) | <5 (<20.00) | 9 (33.33) | 5 (15.15) | 12 (15.19) | 286 (28.04) | 1998 (28.02) |
| Good | 1896 (32.75) | 57 (36.08) | 11 (47.83) | 8 (29.63) | 10 (30.30) | 20 (25.32) | 342 (33.53) | 2344 (32.88) |
| Fair | 1147 (19.81) | 23 (14.56) | 6 (26.09) | <5 (<15.00) | <5 (<15.00) | 19 (24.05) | 183 (17.94) | 1386 (19.44) |
| Poor | 494 (8.53) | 14 (8.86) | <5 (<5.00) | <5 (<15.00) | 7 (21.21) | 20 (25.32) | 88 (8.63) | 627 (8.79) |
| **CES-D** |  |  |  |  |  |  |  |  |
| N= | 5783 | 158 | 23 | 27 | 33 | 79 | 1020 | 7123 |
| **Mean CES-D (95% CI)** | 2.98 (2.95, 3.02) | 2.86 (2.66, 3.06) | 3.30 (2.66, 3.95) | 2.52 (2.17, 2.87) | 3.03 (2.54, 3.52) | 3.48 (3.12, 3.84) | 3.05 (2.97, 3.14) | 2.99 (2.96, 3.03) |

Table S19: Demographic characteristics by sexual identity recoding strategy in older adults

|  | Heterosexual | SM | Total | Heterosexual | SM | Total |
| --- | --- | --- | --- | --- | --- | --- |
| **Recoding strategy** | **R** | **R** | **R** | **B** | **B** | **B** |
| N= | 4513 | 74 | 4587 | 4513 | 253 | 4766 |
| **Sex** |  |  |  |  |  |  |
| Male | 2009 (44.52) | 46 (62.16) | 2055 (44.80) | 2009 (44.52) | 121 (47.83) | 2130 (44.69) |
| Female | 2504 (55.48) | 28 (37.84) | 2532 (55.20) | 2504 (55.48) | 132 (52.17) | 2636 (55.31) |
| **Age bands** |  |  |  |  |  |  |
| 50-59 | 584 (12.94) | 12 (16.22) | 596 (12.99) | 584 (12.94) | 22 (8.70) | 606 (12.72) |
| 60-69 | 2011 (44.56) | 33 (44.59) | 2044 (44.56) | 2011 (44.56) | 90 (35.57) | 2101 (44.08) |
| 70-79 | 1387 (30.73) | 29 (39.19) | 1416 (30.87) | 1387 (30.73) | 88 (34.78) | 1475 (30.95) |
| 80-89 | 490 (10.86) | 0 (0.00) | 490 (10.68) | 490 (10.86) | 49 (19.37) | 539 (11.31) |
| 90+ | 41 (0.91) | 0 (0.00) | 41 (0.89) | 41 (0.91) | 4 (1.58) | 45 (0.94) |
| **Ethnicity** |  |  |  |  |  |  |
| White | 4410 (97.72) | 74 (100.00) | 4484 (97.75) | 4410 (97.72) | 234 (92.49) | 4644 (97.44) |
| Non-white | 103 (2.28) | 0 (0.00) | 103 (2.25) | 103 (2.28) | 19 (7.51) | 122 (2.56) |
| **Income quintile (BU net worth)** |  |  |  |  |  |  |
| Lower quantile | 665 (14.74) | 12 (16.22) | 677 (14.76) | 665 (14.74) | 59 (23.32) | 724 (15.19) |
| Second quantile | 835 (18.50) | 16 (21.62) | 851 (18.55) | 835 (18.50) | 56 (22.13) | 891 (18.69) |
| Third quantile | 938 (20.78) | 12 (16.22) | 950 (20.71) | 938 (20.78) | 59 (23.32) | 997 (20.92) |
| Fourth quantile | 1016 (22.51) | 14 (18.92) | 1030 (22.45) | 1016 (22.51) | 44 (17.39) | 1060 (22.24) |
| Highest quantile | 1037 (22.98) | 20 (27.03) | 1057 (23.04) | 1037 (22.98) | 35 (13.83) | 1072 (22.49) |
| Missing | 22 (0.49) | 0 (0.00) | 22 (0.48) | 22 (0.49) | 0 (0.00) | 22 (0.46) |
| **Income quintile (BU equiv income)** |  |  |  |  |  |  |
| Lower quantile | 776 (17.19) | 14 (18.92) | 790 (17.22) | 776 (17.19) | 64 (25.30) | 840 (17.62) |
| Second quantile | 851 (18.86) | 10 (13.51) | 861 (18.77) | 851 (18.86) | 57 (22.53) | 908 (19.05) |
| Third quantile | 907 (20.10) | 9 (12.16) | 916 (19.97) | 907 (20.10) | 45 (17.79) | 952 (19.97) |
| Fourth quantile | 982 (21.76) | 22 (29.73) | 1004 (21.89) | 982 (21.76) | 51 (20.16) | 1033 (21.67) |
| Highest quantile | 975 (21.60) | 19 (25.68) | 994 (21.67) | 975 (21.60) | 36 (14.23) | 1011 (21.21) |
| Missing | 22 (0.49) | 0 (0.00) | 22 (0.48) | 22 (0.49) | 0 (0.00) | 22 (0.46) |
| **Highest qualification** |  |  |  |  |  |  |
| Missing | 80 (1.77) | 2 (2.70) | 82 (1.79) | 80 (1.77) | 3 (1.19) | 83 (1.74) |
| nvq4/nvq5/degree or equiv | 860 (19.06) | 17 (22.97) | 877 (19.12) | 860 (19.06) | 27 (10.67) | 887 (18.61) |
| higher ed below degree | 654 (14.49) | 11 (14.86) | 665 (14.50) | 654 (14.49) | 24 (9.49) | 678 (14.23) |
| nvq3/gce a level equiv | 421 (9.33) | 10 (13.51) | 431 (9.40) | 421 (9.33) | 20 (7.91) | 441 (9.25) |
| nvq2/gce o level equiv | 901 (19.96) | 11 (14.86) | 912 (19.88) | 901 (19.96) | 38 (15.02) | 939 (19.70) |
| nvq1/cse other grade equiv | 148 (3.28) | 1 (1.35) | 149 (3.25) | 148 (3.28) | 7 (2.77) | 155 (3.25) |
| foreign/other | 658 (14.58) | 13 (17.57) | 671 (14.63) | 658 (14.58) | 38 (15.02) | 696 (14.60) |
| no qualification | 791 (17.53) | 9 (12.16) | 800 (17.44) | 791 (17.53) | 96 (37.94) | 887 (18.61) |
| **Government Office Region** |  |  |  |  |  |  |
| North East | 319 (7.07) | 3 (4.05) | 322 (7.02) | 319 (7.07) | 12 (4.74) | 331 (6.95) |
| North West | 503 (11.15) | 4 (5.41) | 507 (11.05) | 503 (11.15) | 25 (9.88) | 528 (11.08) |
| Yorkshire and the Humber | 442 (9.79) | 4 (5.41) | 446 (9.72) | 442 (9.79) | 24 (9.49) | 466 (9.78) |
| East Midlands | 500 (11.08) | 8 (10.81) | 508 (11.07) | 500 (11.08) | 36 (14.23) | 536 (11.25) |
| West Midlands | 480 (10.64) | 10 (13.51) | 490 (10.68) | 480 (10.64) | 32 (12.65) | 512 (10.74) |
| East of England | 611 (13.54) | 10 (13.51) | 621 (13.54) | 611 (13.54) | 32 (12.65) | 643 (13.49) |
| London | 341 (7.56) | 14 (18.92) | 355 (7.74) | 341 (7.56) | 30 (11.86) | 371 (7.78) |
| South East | 772 (17.11) | 11 (14.86) | 783 (17.07) | 772 (17.11) | 39 (15.42) | 811 (17.02) |
| South West | 531 (11.77) | 10 (13.51) | 541 (11.79) | 531 (11.77) | 23 (9.09) | 554 (11.62) |
| Missing | 4513 | 74 | 4587 | 14 (0.31) | 0 (0.00) | 14 (0.29) |

Table S20: Demographic characteristics by sexual attraction recoding strategy in older adults

|  | Heterosexual | SM | Total | Heterosexual | SM | Total |
| --- | --- | --- | --- | --- | --- | --- |
| **Recoding strategy** | **R** | **R** | **R** | **B** | **B** | **B** |
| N= | 5884 | 122 | 6006 | 5666 | 427 | 6093 |
| **Sex** |  |  |  |  |  |  |
| Male | 2657 (45.16) | 50 (40.98) | 2707 (45.07) | 2586 (45.64) | 140 (32.79) | 2726 (44.74) |
| Female | 3227 (54.84) | 72 (59.02) | 3299 (54.93) | 3080 (54.36) | 287 (67.21) | 3367 (55.26) |
| **Age bands** |  |  |  |  |  |  |
| 50-59 | 637 (10.83) | 16 (13.11) | 653 (10.87) | 594 (10.48) | 62 (14.52) | 656 (10.77) |
| 60-69 | 2394 (40.69) | 42 (34.43) | 2436 (40.56) | 2293 (40.47) | 163 (38.17) | 2456 (40.31) |
| 70-79 | 1886 (32.05) | 49 (40.16) | 1935 (32.22) | 1826 (32.23) | 140 (32.79) | 1966 (32.27) |
| 80-89 | 846 (14.38) | 15 (12.30) | 861 (14.34) | 832 (14.68) | 54 (12.65) | 886 (14.54) |
| 90+ | 121 (2.06) | 0 (0.00) | 121 (2.01) | 121 (2.14) | 8 (1.87) | 129 (2.12) |
| **Ethnicity** |  |  |  |  |  |  |
| White | 5751 (97.74) | 120 (98.36) | 5871 (97.75) | 5536 (97.71) | 415 (97.19) | 5951 (97.67) |
| Non-white | 133 (2.26) | 2 (1.64) | 135 (2.25) | 130 (2.29) | 12 (2.81) | 142 (2.33) |
| **Income quintile (BU net worth)** |  |  |  |  |  |  |
| Lower quantile | 896 (15.23) | 25 (20.49) | 921 (15.33) | 864 (15.25) | 87 (20.37) | 951 (15.61) |
| Second quantile | 1128 (19.17) | 19 (15.57) | 1147 (19.10) | 1085 (19.15) | 90 (21.08) | 1175 (19.28) |
| Third quantile | 1223 (20.79) | 30 (24.59) | 1253 (20.86) | 1187 (20.95) | 85 (19.91) | 1272 (20.88) |
| Fourth quantile | 1323 (22.48) | 25 (20.49) | 1348 (22.44) | 1272 (22.45) | 84 (19.67) | 1356 (22.26) |
| Highest quantile | 1273 (21.63) | 23 (18.85) | 1296 (21.58) | 1217 (21.48) | 81 (18.97) | 1298 (21.30) |
| Missing | 41 (0.70) | 0 (0.00) | 41 (0.68) | 41 (0.72) | 0 (0.00) | 41 (0.67) |
| **Income quintile (BU equiv income)** |  |  |  |  |  |  |
| Lower quantile | 1076 (18.29) | 30 (24.59) | 1106 (18.41) | 1030 (18.18) | 110 (25.76) | 1140 (18.71) |
| Second quantile | 1194 (20.29) | 25 (20.49) | 1219 (20.30) | 1163 (20.53) | 80 (18.74) | 1243 (20.40) |
| Third quantile | 1190 (20.22) | 17 (13.93) | 1207 (20.10) | 1155 (20.38) | 69 (16.16) | 1224 (20.09) |
| Fourth quantile | 1263 (21.46) | 25 (20.49) | 1288 (21.45) | 1219 (21.51) | 77 (18.03) | 1296 (21.27) |
| Highest quantile | 1120 (19.03) | 25 (20.49) | 1145 (19.06) | 1058 (18.67) | 91 (21.31) | 1149 (18.86) |
| Missing | 41 (0.70) | 0 (0.00) | 41 (0.68) | 41 (0.72) | 0 (0.00) | 41 (0.67) |
| **Highest qualification** |  |  |  |  |  |  |
| Missing | 19 (0.32) | 1 (0.82) | 20 (0.33) | 18 (0.32) | 2 (0.47) | 20 (0.33) |
| nvq4/nvq5/degree or equiv | 1051 (17.86) | 20 (16.39) | 1071 (17.83) | 998 (17.61) | 81 (18.97) | 1079 (17.71) |
| higher ed below degree | 846 (14.38) | 20 (16.39) | 866 (14.42) | 818 (14.44) | 57 (13.35) | 875 (14.36) |
| nvq3/gce a level equiv | 537 (9.13) | 11 (9.02) | 548 (9.12) | 504 (8.90) | 48 (11.24) | 552 (9.06) |
| nvq2/gce o level equiv | 1163 (19.77) | 17 (13.93) | 1180 (19.65) | 1124 (19.84) | 66 (15.46) | 1190 (19.53) |
| nvq1/cse other grade equiv | 223 (3.79) | 5 (4.10) | 228 (3.80) | 215 (3.79) | 16 (3.75) | 231 (3.79) |
| foreign/other | 818 (13.90) | 18 (14.75) | 836 (13.92) | 783 (13.82) | 62 (14.52) | 845 (13.87) |
| no qualification | 1227 (20.85) | 30 (24.59) | 1257 (20.93) | 1206 (21.28) | 95 (22.25) | 1301 (21.35) |
| **Government Office Region** |  |  |  |  |  |  |
| North East | 325 (5.52) | 5 (4.10) | 330 (5.49) | 314 (5.54) | 21 (4.92) | 335 (5.50) |
| North West | 650 (11.05) | 12 (9.84) | 662 (11.02) | 634 (11.19) | 37 (8.67) | 671 (11.01) |
| Yorkshire and the Humber | 639 (10.86) | 17 (13.93) | 656 (10.92) | 615 (10.85) | 51 (11.94) | 666 (10.93) |
| East Midlands | 650 (11.05) | 10 (8.20) | 660 (10.99) | 631 (11.14) | 40 (9.37) | 671 (11.01) |
| West Midlands | 577 (9.81) | 12 (9.84) | 589 (9.81) | 549 (9.69) | 49 (11.48) | 598 (9.81) |
| East of England | 842 (14.31) | 14 (11.48) | 856 (14.25) | 814 (14.37) | 52 (12.18) | 866 (14.21) |
| London | 464 (7.89) | 19 (15.57) | 483 (8.04) | 440 (7.77) | 54 (12.65) | 494 (8.11) |
| South East | 1052 (17.88) | 16 (13.11) | 1068 (17.78) | 1011 (17.84) | 73 (17.10) | 1084 (17.79) |
| South West | 667 (11.34) | 17 (13.93) | 684 (11.39) | 640 (11.30) | 50 (11.71) | 690 (11.32) |
| Missing | 18 (0.31) | 0 (0.00) | 18 (0.30) | 18 (0.32) | 0 (0.00) | 18 (0.30) |

Table S21: Demographic characteristics by sexual experience recoding strategy in older adults

|  | Heterosexual | SM | Total | Heterosexual | SM | Total |
| --- | --- | --- | --- | --- | --- | --- |
| **Recoding strategy** | **R** | **R** | **R** | **B** | **B** | **B** |
| N= | 5948 | 83 | 6031 | 5790 | 320 | 6110 |
| **Sex** |  |  |  |  |  |  |
| Male | 2659 (44.70) | 39 (46.99) | 2698 (44.74) | 2574 (44.46) | 151 (47.19) | 2725 (44.60) |
| Female | 3289 (55.30) | 44 (53.01) | 3333 (55.26) | 3216 (55.54) | 169 (52.81) | 3385 (55.40) |
| **Age bands** |  |  |  |  |  |  |
| 50-59 | 640 (10.76) | 16 (19.28) | 656 (10.88) | 616 (10.64) | 42 (13.13) | 658 (10.77) |
| 60-69 | 2404 (40.42) | 31 (37.35) | 2435 (40.37) | 2319 (40.05) | 136 (42.50) | 2455 (40.18) |
| 70-79 | 1916 (32.21) | 29 (34.94) | 1945 (32.25) | 1878 (32.44) | 95 (29.69) | 1973 (32.29) |
| 80-89 | 865 (14.54) | 7 (8.43) | 872 (14.46) | 854 (14.75) | 39 (12.19) | 893 (14.62) |
| 90+ | 123 (2.07) | 0 (0.00) | 123 (2.04) | 123 (2.12) | 8 (2.50) | 131 (2.14) |
| **Ethnicity** |  |  |  |  |  |  |
| White | 5810 (97.68) | 82 (98.80) | 5892 (97.70) | 5658 (97.72) | 309 (96.56) | 5967 (97.66) |
| Non-white | 138 (2.32) | 1 (1.20) | 139 (2.30) | 132 (2.28) | 11 (3.44) | 143 (2.34) |
| **Income quintile (BU net worth)** |  |  |  |  |  |  |
| Lower quantile | 915 (15.38) | 12 (14.46) | 927 (15.37) | 887 (15.32) | 76 (23.75) | 963 (15.76) |
| Second quantile | 1146 (19.27) | 14 (16.87) | 1160 (19.23) | 1127 (19.46) | 50 (15.63) | 1177 (19.26) |
| Third quantile | 1237 (20.80) | 15 (18.07) | 1252 (20.76) | 1204 (20.79) | 66 (20.63) | 1270 (20.79) |
| Fourth quantile | 1332 (22.39) | 21 (25.30) | 1353 (22.43) | 1292 (22.31) | 67 (20.94) | 1359 (22.24) |
| Highest quantile | 1277 (21.47) | 21 (25.30) | 1298 (21.52) | 1240 (21.42) | 60 (18.75) | 1300 (21.28) |
| Missing | 41 (0.69) | 0 (0.00) | 41 (0.68) | 40 (0.69) | 1 (0.31) | 41 (0.67) |
| **Income quintile (BU equiv income)** |  |  |  |  |  |  |
| Lower quantile | 1103 (18.54) | 18 (21.69) | 1121 (18.59) | 1070 (18.48) | 80 (25.00) | 1150 (18.82) |
| Second quantile | 1212 (20.38) | 14 (16.87) | 1226 (20.33) | 1188 (20.52) | 56 (17.50) | 1244 (20.36) |
| Third quantile | 1199 (20.16) | 10 (12.05) | 1209 (20.05) | 1173 (20.26) | 56 (17.50) | 1229 (20.11) |
| Fourth quantile | 1269 (21.33) | 18 (21.69) | 1287 (21.34) | 1239 (21.40) | 56 (17.50) | 1295 (21.19) |
| Highest quantile | 1124 (18.90) | 23 (27.71) | 1147 (19.02) | 1080 (18.65) | 71 (22.19) | 1151 (18.84) |
| Missing | 41 (0.69) | 0 (0.00) | 41 (0.68) | 40 (0.69) | 1 (0.31) | 41 (0.67) |
| **Highest qualification** |  |  |  |  |  |  |
| Missing | 19 (0.32) | 1 (1.20) | 20 (0.33) | 18 (0.31) | 2 (0.63) | 20 (0.33) |
| nvq4/nvq5/degree or equiv | 1050 (17.65) | 21 (25.30) | 1071 (17.76) | 1001 (17.29) | 80 (25.00) | 1081 (17.69) |
| higher ed below degree | 856 (14.39) | 11 (13.25) | 867 (14.38) | 833 (14.39) | 41 (12.81) | 874 (14.30) |
| nvq3/gce a level equiv | 542 (9.11) | 7 (8.43) | 549 (9.10) | 516 (8.91) | 36 (11.25) | 552 (9.03) |
| nvq2/gce o level equiv | 1175 (19.75) | 11 (13.25) | 1186 (19.67) | 1148 (19.83) | 45 (14.06) | 1193 (19.53) |
| nvq1/cse other grade equiv | 227 (3.82) | 2 (2.41) | 229 (3.80) | 227 (3.92) | 4 (1.25) | 231 (3.78) |
| foreign/other | 832 (13.99) | 12 (14.46) | 844 (13.99) | 813 (14.04) | 36 (11.25) | 849 (13.90) |
| no qualification | 1247 (20.97) | 18 (21.69) | 1265 (20.97) | 1234 (21.31) | 76 (23.75) | 1310 (21.44) |
| **Government Office Region** |  |  |  |  |  |  |
| North East | 327 (5.50) | 3 (3.61) | 330 (5.47) | 322 (5.56) | 15 (4.69) | 337 (5.52) |
| North West | 655 (11.01) | 9 (10.84) | 664 (11.01) | 639 (11.04) | 35 (10.94) | 674 (11.03) |
| Yorkshire and the Humber | 650 (10.93) | 9 (10.84) | 659 (10.93) | 635 (10.97) | 35 (10.94) | 670 (10.97) |
| East Midlands | 658 (11.06) | 3 (3.61) | 661 (10.96) | 646 (11.16) | 23 (7.19) | 669 (10.95) |
| West Midlands | 582 (9.78) | 8 (9.64) | 590 (9.78) | 565 (9.76) | 33 (10.31) | 598 (9.79) |
| East of England | 854 (14.36) | 7 (8.43) | 861 (14.28) | 830 (14.34) | 38 (11.88) | 868 (14.21) |
| London | 470 (7.90) | 16 (19.28) | 486 (8.06) | 449 (7.75) | 48 (15.00) | 497 (8.13) |
| South East | 1062 (17.85) | 14 (16.87) | 1076 (17.84) | 1032 (17.82) | 55 (17.19) | 1087 (17.79) |
| South West | 672 (11.30) | 14 (16.87) | 686 (11.37) | 654 (11.30) | 38 (11.88) | 692 (11.33) |
| Missing | 18 (0.30) | 0 (0.00) | 18 (0.30) | 18 (0.31) | 0 (0.00) | 18 (0.29) |

Table S22: Demographic characteristics by combined dimension recoding strategy in older adults

|  | Heterosexual | SM | Total | Heterosexual | SM | Total |
| --- | --- | --- | --- | --- | --- | --- |
| **Recoding strategy** | **R** | **R** | **R** | **B** | **B** | **B** |
| N= | 6944 | 84 | 7028 | 6426 | 704 | 7130 |
| **Sex** |  |  |  |  |  |  |
| Male | 3102 (44.67) | 49 (58.33) | 3151 (44.83) | 2908 (45.25) | 276 (39.20) | 3184 (44.66) |
| Female | 3842 (55.33) | 35 (41.67) | 3877 (55.17) | 3518 (54.75) | 428 (60.80) | 3946 (55.34) |
| **Age bands** |  |  |  |  |  |  |
| 50-59 | 889 (12.80) | 20 (23.81) | 909 (12.93) | 824 (12.82) | 92 (13.07) | 916 (12.85) |
| 60-69 | 2767 (39.85) | 31 (36.90) | 2798 (39.81) | 2548 (39.65) | 276 (39.20) | 2824 (39.61) |
| 70-79 | 2179 (31.38) | 27 (32.14) | 2206 (31.39) | 2017 (31.39) | 216 (30.68) | 2233 (31.32) |
| 80-89 | 976 (14.06) | 6 (7.14) | 982 (13.97) | 906 (14.10) | 109 (15.48) | 1015 (14.24) |
| 90+ | 133 (1.92) | 0 (0.00) | 133 (1.89) | 131 (2.04) | 11 (1.56) | 142 (1.99) |
| **Ethnicity** |  |  |  |  |  |  |
| White | 6762 (97.38) | 84 (100.00) | 6846 (97.41) | 6267 (97.53) | 670 (95.17) | 6937 (97.29) |
| Non-white | 182 (2.62) | 0 (0.00) | 182 (2.59) | 159 (2.47) | 34 (4.83) | 193 (2.71) |
| **Income quintile (BU net worth)** |  |  |  |  |  |  |
| Lower quantile | 1118 (16.10) | 14 (16.67) | 1132 (16.11) | 1013 (15.76) | 151 (21.45) | 1164 (16.33) |
| Second quantile | 1360 (19.59) | 19 (22.62) | 1379 (19.62) | 1253 (19.50) | 149 (21.16) | 1402 (19.66) |
| Third quantile | 1433 (20.64) | 13 (15.48) | 1446 (20.57) | 1323 (20.59) | 152 (21.59) | 1475 (20.69) |
| Fourth quantile | 1502 (21.63) | 17 (20.24) | 1519 (21.61) | 1399 (21.77) | 131 (18.61) | 1530 (21.46) |
| Highest quantile | 1478 (21.28) | 21 (25.00) | 1499 (21.33) | 1386 (21.57) | 120 (17.05) | 1506 (21.12) |
| Missing | 53 (0.76) | 0 (0.00) | 53 (0.75) | 52 (0.81) | 1 (0.14) | 53 (0.74) |
| **Income quintile (BU equiv income)** |  |  |  |  |  |  |
| Lower quantile | 1296 (18.66) | 21 (25.00) | 1317 (18.74) | 1167 (18.16) | 180 (25.57) | 1347 (18.89) |
| Second quantile | 1397 (20.12) | 12 (14.29) | 1409 (20.05) | 1297 (20.18) | 140 (19.89) | 1437 (20.15) |
| Third quantile | 1406 (20.25) | 9 (10.71) | 1415 (20.13) | 1317 (20.49) | 118 (16.76) | 1435 (20.13) |
| Fourth quantile | 1466 (21.11) | 23 (27.38) | 1489 (21.19) | 1376 (21.41) | 127 (18.04) | 1503 (21.08) |
| Highest quantile | 1326 (19.10) | 19 (22.62) | 1345 (19.14) | 1217 (18.94) | 138 (19.60) | 1355 (19.00) |
| Missing | 53 (0.76) | 0 (0.00) | 53 (0.75) | 52 (0.81) | 1 (0.14) | 53 (0.74) |
| **Highest qualification** |  |  |  |  |  |  |
| Missing | 91 (1.31) | 1 (1.19) | 92 (1.31) | 90 (1.40) | 3 (0.43) | 93 (1.30) |
| nvq4/nvq5/degree or equiv | 1199 (17.27) | 19 (22.62) | 1218 (17.33) | 1106 (17.21) | 124 (17.61) | 1230 (17.25) |
| higher ed below degree | 978 (14.08) | 12 (14.29) | 990 (14.09) | 911 (14.18) | 84 (11.93) | 995 (13.96) |
| nvq3/gce a level equiv | 623 (8.97) | 11 (13.10) | 634 (9.02) | 571 (8.89) | 69 (9.80) | 640 (8.98) |
| nvq2/gce o level equiv | 1364 (19.64) | 10 (11.90) | 1374 (19.55) | 1279 (19.90) | 108 (15.34) | 1387 (19.45) |
| nvq1/cse other grade equiv | 254 (3.66) | 2 (2.38) | 256 (3.64) | 237 (3.69) | 22 (3.13) | 259 (3.63) |
| foreign/other | 956 (13.77) | 13 (15.48) | 969 (13.79) | 883 (13.74) | 96 (13.64) | 979 (13.73) |
| no qualification | 1479 (21.30) | 16 (19.05) | 1495 (21.27) | 1349 (20.99) | 198 (28.13) | 1547 (21.70) |
| **Government Office Region** |  |  |  |  |  |  |
| North East | 418 (6.02) | 3 (3.57) | 421 (5.99) | 396 (6.16) | 35 (4.97) | 431 (6.04) |
| North West | 787 (11.33) | 9 (10.71) | 796 (11.33) | 736 (11.45) | 75 (10.65) | 811 (11.37) |
| Yorkshire and the Humber | 736 (10.60) | 8 (9.52) | 744 (10.59) | 682 (10.61) | 76 (10.80) | 758 (10.63) |
| East Midlands | 770 (11.09) | 7 (8.33) | 777 (11.06) | 705 (10.97) | 80 (11.36) | 785 (11.01) |
| West Midlands | 719 (10.35) | 12 (14.29) | 731 (10.40) | 664 (10.33) | 81 (11.51) | 745 (10.45) |
| East of England | 937 (13.49) | 4 (4.76) | 941 (13.39) | 868 (13.51) | 84 (11.93) | 952 (13.35) |
| London | 541 (7.79) | 16 (19.05) | 557 (7.93) | 489 (7.61) | 83 (11.79) | 572 (8.02) |
| South East | 1203 (17.32) | 12 (14.29) | 1215 (17.29) | 1109 (17.26) | 116 (16.48) | 1225 (17.18) |
| South West | 811 (11.68) | 13 (15.48) | 824 (11.72) | 755 (11.75) | 74 (10.51) | 829 (11.63) |
| Missing | 22 (0.32) | 0 (0.00) | 22 (0.31) | 22 (0.34) | 0 (0.00) | 22 (0.31) |

Table S23: Demographic characteristics by sexual identity recoding strategy in adolescents

|  | Heterosexual | SM | Total | Heterosexual | SM | Total |
| --- | --- | --- | --- | --- | --- | --- |
| Strategy | **R** | **R** | **R** | **B** | **B** | **B** |
| N | 8989 | 906 | 9895 | 7888 | 2215 | 10103 |
| **Gender Identity (Four category)** |  |  |  |  |  |  |
| Male | 4553 (50.65) | 278 (30.68) | 4831 (48.82) | 4154 (52.66) | 730 (32.96) | 4884 (48.34) |
| Female | 4410 (49.06) | 582 (64.24) | 4992 (50.45) | 3719 (47.15) | 1372 (61.94) | 5091 (50.39) |
| Non-binary/Other | 2 (0.02) | 31 (3.42) | 33 (0.33) | 0 (0.00) | 58 (2.62) | 58 (0.57) |
| Don’t know / PNS | 14 (0.16) | 15 (1.66) | 29 (0.29) | 9 (0.11) | 50 (2.26) | 59 (0.58) |
| Missing | 10 (0.11) | 0 (0.00) | 10 (0.10) | 6 (0.08) | 5 (0.23) | 11 (0.11) |
| **Ethnicity** |  |  |  |  |  |  |
| White | 7162 (79.69) | 814 (89.85) | 7976 (80.62) | 6233 (79.04) | 1907 (86.09) | 8140 (80.59) |
| **OECD Equivalised income quintiles** |  |  |  |  |  |  |
| Lower quantile | 1433 (15.94) | 116 (12.80) | 1549 (15.65) | 1325 (16.80) | 266 (12.01) | 1591 (15.75) |
| Second quantile | 1383 (15.39) | 148 (16.34) | 1531 (15.47) | 1238 (15.69) | 338 (15.26) | 1576 (15.60) |
| Third quantile | 1784 (19.85) | 169 (18.65) | 1953 (19.74) | 1573 (19.94) | 415 (18.74) | 1988 (19.68) |
| Fourth quantile | 2103 (23.40) | 204 (22.52) | 2307 (23.31) | 1832 (23.23) | 514 (23.21) | 2346 (23.22) |
| Highest quantile | 2095 (23.31) | 249 (27.48) | 2344 (23.69) | 1753 (22.22) | 634 (28.62) | 2387 (23.63) |
| Missing | 191 (2.12) | 20 (2.21) | 211 (2.13) | 167 (2.12) | 48 (2.17) | 215 (2.13) |
| **Highest parent/guardian qualification (NVQ Level)** |  |  |  |  |  |  |
| NVQ Level 1 | 349 (3.88) | 31 (3.42) | 380 (3.84) | 328 (4.16) | 60 (2.71) | 388 (3.84) |
| NVQ Level 2 | 1555 (17.30) | 148 (16.34) | 1703 (17.21) | 1388 (17.60) | 354 (15.98) | 1742 (17.24) |
| NVQ Level 3 | 1250 (13.91) | 116 (12.80) | 1366 (13.80) | 1135 (14.39) | 257 (11.60) | 1392 (13.78) |
| NVQ Level 4 | 3308 (36.80) | 346 (38.19) | 3654 (36.93) | 2863 (36.30) | 862 (38.92) | 3725 (36.87) |
| NVQ Level 5 | 1689 (18.79) | 192 (21.19) | 1881 (19.01) | 1403 (17.79) | 510 (23.02) | 1913 (18.93) |
| None | 406 (4.52) | 29 (3.20) | 435 (4.40) | 379 (4.80) | 75 (3.39) | 454 (4.49) |
| Overseas/Other | 180 (2.00) | 20 (2.21) | 200 (2.02) | 168 (2.13) | 38 (1.72) | 206 (2.04) |
| Missing | 252 (2.80) | 24 (2.65) | 276 (2.79) | 224 (2.84) | 59 (2.66) | 283 (2.80) |
| **ONS Urban/Rural Classification 2005** |  |  |  |  |  |  |
| Rural | 2081 (23.15) | 187 (20.64) | 2268 (22.92) | 1783 (22.60) | 527 (23.79) | 2310 (22.86) |
| Urban | 6609 (73.52) | 691 (76.27) | 7300 (73.77) | 5841 (74.05) | 1618 (73.05) | 7459 (73.83) |
| Missing | 299 (3.33) | 28 (3.09) | 327 (3.30) | 264 (3.35) | 70 (3.16) | 334 (3.31) |

Table S24: Demographic characteristics by sexual attraction recoding strategy in adolescents

|  | Heterosexual | SM | Total | Heterosexual | SM | Total |
| --- | --- | --- | --- | --- | --- | --- |
| Strategy | **R** | **R** | **R** | **B** | **B** | **B** |
| N | 9038 | 716 | 9754 | 7656 | 2446 | 10102 |
| **Gender Identity (Four category)** |  |  |  |  |  |  |
| Male | 4554 (50.39) | 223 (31.15) | 4777 (48.97) | 4155 (54.27) | 729 (29.80) | 4884 (48.35) |
| Female | 4445 (49.18) | 439 (61.31) | 4884 (50.07) | 3476 (45.40) | 1615 (66.03) | 5091 (50.40) |
| Non-binary/Other | 10 (0.11) | 37 (5.17) | 47 (0.48) | 2 (0.03) | 55 (2.25) | 57 (0.56) |
| Don’t know / PNS | 19 (0.21) | 17 (2.37) | 36 (0.37) | 15 (0.20) | 44 (1.80) | 59 (0.58) |
| Missing | 10 (0.11) | 0 (0.00) | 10 (0.10) | 8 (0.10) | 3 (0.12) | 11 (0.11) |
| **Ethnicity** |  |  |  |  |  |  |
| White | 7276 (80.52) | 637 (88.97) | 7913 (81.14) | 6072 (79.33) | 2067 (84.51) | 8139 (80.58) |
| **OECD Equivalised income quintiles** |  |  |  |  |  |  |
| Lower quantile | 1391 (15.39) | 94 (13.13) | 1485 (15.22) | 1265 (16.52) | 326 (13.33) | 1591 (15.75) |
| Second quantile | 1389 (15.37) | 124 (17.32) | 1513 (15.51) | 1198 (15.65) | 378 (15.45) | 1576 (15.60) |
| Third quantile | 1805 (19.97) | 131 (18.30) | 1936 (19.85) | 1517 (19.81) | 471 (19.26) | 1988 (19.68) |
| Fourth quantile | 2137 (23.64) | 151 (21.09) | 2288 (23.46) | 1781 (23.26) | 565 (23.10) | 2346 (23.22) |
| Highest quantile | 2127 (23.53) | 197 (27.51) | 2324 (23.83) | 1736 (22.68) | 650 (26.57) | 2386 (23.62) |
| Missing | 189 (2.09) | 19 (2.65) | 208 (2.13) | 159 (2.08) | 56 (2.29) | 215 (2.13) |
| **Highest parent/guardian qualification (NVQ Level)** |  |  |  |  |  |  |
| NVQ Level 1 | 340 (3.76) | 22 (3.07) | 362 (3.71) | 305 (3.98) | 83 (3.39) | 388 (3.84) |
| NVQ Level 2 | 1556 (17.22) | 117 (16.34) | 1673 (17.15) | 1345 (17.57) | 397 (16.23) | 1742 (17.24) |
| NVQ Level 3 | 1264 (13.99) | 88 (12.29) | 1352 (13.86) | 1091 (14.25) | 301 (12.31) | 1392 (13.78) |
| NVQ Level 4 | 3337 (36.92) | 267 (37.29) | 3604 (36.95) | 2774 (36.23) | 951 (38.88) | 3725 (36.87) |
| NVQ Level 5 | 1716 (18.99) | 158 (22.07) | 1874 (19.21) | 1394 (18.21) | 518 (21.18) | 1912 (18.93) |
| None | 392 (4.34) | 27 (3.77) | 419 (4.30) | 361 (4.72) | 93 (3.80) | 454 (4.49) |
| Overseas/Other | 181 (2.00) | 17 (2.37) | 198 (2.03) | 172 (2.25) | 34 (1.39) | 206 (2.04) |
| Missing | 252 (2.79) | 20 (2.79) | 272 (2.79) | 214 (2.80) | 69 (2.82) | 283 (2.80) |
| **ONS Urban/Rural Classification 2005** |  |  |  |  |  |  |
| Rural | 2119 (23.45) | 134 (18.72) | 2253 (23.10) | 1747 (22.82) | 562 (22.98) | 2309 (22.86) |
| Urban | 6620 (73.25) | 558 (77.93) | 7178 (73.59) | 5653 (73.84) | 1806 (73.83) | 7459 (73.84) |
| Missing | 299 (3.31) | 24 (3.35) | 323 (3.31) | 256 (3.34) | 78 (3.19) | 334 (3.31) |

Table S25: Demographic characteristics by combined dimension recoding strategy in adolescents

|  | Heterosexual | SM | Total | Heterosexual | SM | Total |
| --- | --- | --- | --- | --- | --- | --- |
| Strategy | **R** | **R** | **R** | **B** | **B** | **B** |
| N | 9291 | 480 | 9771 | 7335 | 2746 | 10081 |
| **Gender Identity (Four category)** |  |  |  |  |  |  |
| Male | 4636 (49.90) | 121 (25.21) | 4757 (48.68) | 4011 (54.68) | 867 (31.57) | 4878 (48.39) |
| Female | 4614 (49.66) | 333 (69.38) | 4947 (50.63) | 3310 (45.13) | 1765 (64.28) | 5075 (50.34) |
| Non-binary/Other | 10 (0.11) | 16 (3.33) | 26 (0.27) | 0 (0.00) | 58 (2.11) | 58 (0.58) |
| Don’t know / PNS | 21 (0.23) | 10 (2.08) | 31 (0.32) | 8 (0.11) | 51 (1.86) | 59 (0.59) |
| Missing | 10 (0.11) | 0 (0.00) | 10 (0.10) | 6 (0.08) | 5 (0.18) | 11 (0.11) |
| **Ethnicity** |  |  |  |  |  |  |
| White | 7427 (79.95) | 427 (88.96) | 7854 (80.40) | 5820 (79.37) | 2308 (84.05) | 8128 (80.64) |
| **OECD Equivalised income quintiles** |  |  |  |  |  |  |
| Lower quantile | 1472 (15.84) | 62 (12.92) | 1534 (15.70) | 1213 (16.54) | 367 (13.36) | 1580 (15.67) |
| Second quantile | 1429 (15.38) | 71 (14.79) | 1500 (15.35) | 1155 (15.75) | 418 (15.22) | 1573 (15.60) |
| Third quantile | 1846 (19.87) | 89 (18.54) | 1935 (19.80) | 1459 (19.89) | 524 (19.08) | 1983 (19.67) |
| Fourth quantile | 2177 (23.43) | 107 (22.29) | 2284 (23.38) | 1708 (23.29) | 636 (23.16) | 2344 (23.25) |
| Highest quantile | 2171 (23.37) | 138 (28.75) | 2309 (23.63) | 1649 (22.48) | 737 (26.84) | 2386 (23.67) |
| Missing | 196 (2.11) | 13 (2.71) | 209 (2.14) | 151 (2.06) | 64 (2.33) | 215 (2.13) |
| **Highest parent/guardian qualification (NVQ Level)** |  |  |  |  |  |  |
| NVQ Level 1 | 359 (3.86) | 14 (2.92) | 373 (3.82) | 298 (4.06) | 89 (3.24) | 387 (3.84) |
| NVQ Level 2 | 1605 (17.27) | 75 (15.63) | 1680 (17.19) | 1283 (17.49) | 452 (16.46) | 1735 (17.21) |
| NVQ Level 3 | 1288 (13.86) | 54 (11.25) | 1342 (13.73) | 1060 (14.45) | 330 (12.02) | 1390 (13.79) |
| NVQ Level 4 | 3427 (36.89) | 186 (38.75) | 3613 (36.98) | 2664 (36.32) | 1058 (38.53) | 3722 (36.92) |
| NVQ Level 5 | 1749 (18.82) | 113 (23.54) | 1862 (19.06) | 1317 (17.96) | 594 (21.63) | 1911 (18.96) |
| None | 418 (4.50) | 12 (2.50) | 430 (4.40) | 344 (4.69) | 103 (3.75) | 447 (4.43) |
| Overseas/Other | 185 (1.99) | 12 (2.50) | 197 (2.02) | 164 (2.24) | 42 (1.53) | 206 (2.04) |
| Missing | 260 (2.80) | 14 (2.92) | 274 (2.80) | 205 (2.79) | 78 (2.84) | 283 (2.81) |
| **ONS Urban/Rural Classification 2005** |  |  |  |  |  |  |
| Rural | 2160 (23.25) | 99 (20.63) | 2259 (23.12) | 1682 (22.93) | 627 (22.83) | 2309 (22.90) |
| Urban | 6823 (73.44) | 363 (75.63) | 7186 (73.54) | 5410 (73.76) | 2028 (73.85) | 7438 (73.78) |
| Missing | 308 (3.32) | 18 (3.75) | 326 (3.34) | 243 (3.31) | 91 (3.31) | 334 (3.31) |

Table S26: Health outcomes by sexual identity recoding strategy in older adults

|  | Heterosexual | SM | Total | Heterosexual | SM | Total |
| --- | --- | --- | --- | --- | --- | --- |
| **Recoding strategy** | **R** | **R** | **R** | **B** | **B** | **B** |
| N= | 4513 | 74 | 4587 | 4513 | 253 | 4766 |
| **Life Satisfaction** |  |  |  |  |  |  |
| Missing | 9 (0.20) | 0 (0.00) | 9 (0.20) | 9 (0.20) | 0 (0.00) | 9 (0.19) |
| Strongly agree | 693 (15.36) | 13 (17.57) | 706 (15.39) | 693 (15.36) | 47 (18.58) | 740 (15.53) |
| Agree | 2221 (49.21) | 34 (45.95) | 2255 (49.16) | 2221 (49.21) | 116 (45.85) | 2337 (49.03) |
| Slightly agree | 710 (15.73) | 9 (12.16) | 719 (15.67) | 710 (15.73) | 33 (13.04) | 743 (15.59) |
| Neither agree nor disagree | 353 (7.82) | 7 (9.46) | 360 (7.85) | 353 (7.82) | 27 (10.67) | 380 (7.97) |
| Slightly disagree | 283 (6.27) | 5 (6.76) | 288 (6.28) | 283 (6.27) | 16 (6.32) | 299 (6.27) |
| Disagree | 188 (4.17) | 4 (5.41) | 192 (4.19) | 188 (4.17) | 12 (4.74) | 200 (4.20) |
| Strongly disagree | 56 (1.24) | 2 (2.70) | 58 (1.26) | 56 (1.24) | 2 (0.79) | 58 (1.22) |
| **Self-rated General Health** |  |  |  |  |  |  |
| Missing | 566 (12.54) | 11 (14.86) | 577 (12.58) | 566 (12.54) | 21 (8.30) | 587 (12.32) |
| excellent | 1362 (30.18) | 17 (22.97) | 1379 (30.06) | 1362 (30.18) | 55 (21.74) | 1417 (29.73) |
| very good | 1487 (32.95) | 25 (33.78) | 1512 (32.96) | 1487 (32.95) | 79 (31.23) | 1566 (32.86) |
| Good | 797 (17.66) | 10 (13.51) | 807 (17.59) | 797 (17.66) | 67 (26.48) | 864 (18.13) |
| Fair | 301 (6.67) | 11 (14.86) | 312 (6.80) | 301 (6.67) | 31 (12.25) | 332 (6.97) |
| Poor |  |  |  |  |  |  |
| **CES-D** |  |  |  |  |  |  |
| **Mean CES-D (95% CI)** | 2.91 (2.87, 2.95) | 2.99 (2.69, 3.29) | 2.91 (2.87, 2.95) | 2.91 (2.87, 2.95) | 3.20 (3.02, 3.37) | 2.93 (2.89, 2.96) |

Table S27: Health outcomes by sexual attraction recoding strategy in older adults

|  | Heterosexual | SM | Total | Heterosexual | SM | Total |
| --- | --- | --- | --- | --- | --- | --- |
| **Recoding strategy** | **R** | **R** | **R** | **B** | **B** | **B** |
| N= | 5884 | 122 | 6006 | 5666 | 427 | 6093 |
| **Life Satisfaction** |  |  |  |  |  |  |
| Missing | 43 (0.73) | 1 (0.82) | 44 (0.73) | 43 (0.76) | 2 (0.47) | 45 (0.74) |
| Strongly agree | 875 (14.87) | 27 (22.13) | 902 (15.02) | 849 (14.98) | 70 (16.39) | 919 (15.08) |
| Agree | 2748 (46.70) | 58 (47.54) | 2806 (46.72) | 2640 (46.59) | 204 (47.78) | 2844 (46.68) |
| Slightly agree | 958 (16.28) | 9 (7.38) | 967 (16.10) | 931 (16.43) | 49 (11.48) | 980 (16.08) |
| Neither agree nor disagree | 499 (8.48) | 6 (4.92) | 505 (8.41) | 478 (8.44) | 31 (7.26) | 509 (8.35) |
| Slightly disagree | 378 (6.42) | 9 (7.38) | 387 (6.44) | 357 (6.30) | 38 (8.90) | 395 (6.48) |
| Disagree | 274 (4.66) | 8 (6.56) | 282 (4.70) | 264 (4.66) | 21 (4.92) | 285 (4.68) |
| Strongly disagree | 109 (1.85) | 4 (3.28) | 113 (1.88) | 104 (1.84) | 12 (2.81) | 116 (1.90) |
| **Self-rated General Health** |  |  |  |  |  |  |
| Missing | 1 (0.02) | 0 (0.00) | 1 (0.02) | 0 (0.00) | 1 (0.23) | 1 (0.02) |
| excellent | 629 (10.69) | 15 (12.30) | 644 (10.72) | 603 (10.64) | 50 (11.71) | 653 (10.72) |
| very good | 1676 (28.48) | 25 (20.49) | 1701 (28.32) | 1605 (28.33) | 105 (24.59) | 1710 (28.06) |
| Good | 1929 (32.78) | 41 (33.61) | 1970 (32.80) | 1857 (32.77) | 138 (32.32) | 1995 (32.74) |
| Fair | 1153 (19.60) | 26 (21.31) | 1179 (19.63) | 1118 (19.73) | 81 (18.97) | 1199 (19.68) |
| Poor | 496 (8.43) | 15 (12.30) | 511 (8.51) | 483 (8.52) | 52 (12.18) | 535 (8.78) |
| **CES-D** |  |  |  |  |  |  |
| **Mean CES-D (95% CI)** | 2.98 (2.94, 3.01) | 3.03 (2.79, 3.28) | 2.98 (2.94, 3.01) | 2.98 (2.94, 3.01) | 3.12 (2.98, 3.26) | 2.99 (2.95, 3.02) |

Table S28: Health outcomes by sexual experience recoding strategy in older adults

|  | Heterosexual | SM | Total | Heterosexual | SM | Total |
| --- | --- | --- | --- | --- | --- | --- |
| **Recoding strategy** | **R** | **R** | **R** | **B** | **B** | **B** |
| N= | 5948 | 83 | 6031 | 5790 | 320 | 6110 |
| **Life Satisfaction** |  |  |  |  |  |  |
| Missing | 41 (0.69) | 4 (4.82) | 45 (0.75) | 41 (0.71) | 6 (1.88) | 47 (0.77) |
| Strongly agree | 888 (14.93) | 15 (18.07) | 903 (14.97) | 870 (15.03) | 54 (16.88) | 924 (15.12) |
| Agree | 2785 (46.82) | 41 (49.40) | 2826 (46.86) | 2702 (46.67) | 149 (46.56) | 2851 (46.66) |
| Slightly agree | 966 (16.24) | 6 (7.23) | 972 (16.12) | 944 (16.30) | 37 (11.56) | 981 (16.06) |
| Neither agree nor disagree | 496 (8.34) | 4 (4.82) | 500 (8.29) | 488 (8.43) | 22 (6.88) | 510 (8.35) |
| Slightly disagree | 385 (6.47) | 4 (4.82) | 389 (6.45) | 375 (6.48) | 21 (6.56) | 396 (6.48) |
| Disagree | 278 (4.67) | 6 (7.23) | 284 (4.71) | 269 (4.65) | 16 (5.00) | 285 (4.66) |
| Strongly disagree | 109 (1.83) | 3 (3.61) | 112 (1.86) | 101 (1.74) | 15 (4.69) | 116 (1.90) |
| **Self-rated General Health** |  |  |  |  |  |  |
| Missing | 1 (0.02) | 0 (0.00) | 1 (0.02) | 0 (0.00) | 1 (0.31) | 1 (0.02) |
| excellent | 634 (10.66) | 11 (13.25) | 645 (10.69) | 610 (10.54) | 43 (13.44) | 653 (10.69) |
| very good | 1682 (28.28) | 18 (21.69) | 1700 (28.19) | 1643 (28.38) | 69 (21.56) | 1712 (28.02) |
| Good | 1953 (32.83) | 29 (34.94) | 1982 (32.86) | 1896 (32.75) | 106 (33.13) | 2002 (32.77) |
| Fair | 1170 (19.67) | 14 (16.87) | 1184 (19.63) | 1147 (19.81) | 56 (17.50) | 1203 (19.69) |
| Poor | 508 (8.54) | 11 (13.25) | 519 (8.61) | 494 (8.53) | 45 (14.06) | 539 (8.82) |
| **CES-D** |  |  |  |  |  |  |
| **Mean CES-D (95% CI)** | 2.98 (2.94, 3.01) | 2.94 (2.65, 3.23) | 2.91 (2.87, 2.95) | 2.98 (2.95, 3.02) | 3.03 (2.88, 3.19) | 2.98 (2.95, 3.02) |

Table S29: Health outcomes by combined dimension recoding strategy in older adults

|  | Heterosexual | SM | Total | Heterosexual | SM | Total |
| --- | --- | --- | --- | --- | --- | --- |
| **Recoding strategy** | **R** | **R** | **R** | **B** | **B** | **B** |
| N= | 6944 | 84 | 7028 | 6426 | 704 | 7130 |
| **Life Satisfaction** |  |  |  |  |  |  |
| Missing | 53 (0.76) | 1 (1.19) | 54 (0.77) | 49 (0.76) | 6 (0.85) | 55 (0.77) |
| Strongly agree | 1030 (14.83) | 15 (17.86) | 1045 (14.87) | 956 (14.88) | 113 (16.05) | 1069 (14.99) |
| Agree | 3261 (46.96) | 37 (44.05) | 3298 (46.93) | 3013 (46.89) | 328 (46.59) | 3341 (46.86) |
| Slightly agree | 1128 (16.24) | 10 (11.90) | 1138 (16.19) | 1061 (16.51) | 89 (12.64) | 1150 (16.13) |
| Neither agree nor disagree | 578 (8.32) | 5 (5.95) | 583 (8.30) | 528 (8.22) | 63 (8.95) | 591 (8.29) |
| Slightly disagree | 444 (6.39) | 6 (7.14) | 450 (6.40) | 406 (6.32) | 52 (7.39) | 458 (6.42) |
| Disagree | 323 (4.65) | 7 (8.33) | 330 (4.70) | 301 (4.68) | 34 (4.83) | 335 (4.70) |
| Strongly disagree | 127 (1.83) | 3 (3.57) | 130 (1.85) | 112 (1.74) | 19 (2.70) | 131 (1.84) |
| **Self-rated General Health** |  |  |  |  |  |  |
| Missing | 1 (0.01) | 0 (0.00) | 1 (0.01) | 0 (0.00) | 1 (0.14) | 1 (0.01) |
| excellent | 754 (10.86) | 10 (11.90) | 764 (10.87) | 698 (10.86) | 76 (10.80) | 774 (10.86) |
| very good | 1965 (28.30) | 17 (20.24) | 1982 (28.20) | 1835 (28.56) | 163 (23.15) | 1998 (28.02) |
| Good | 2283 (32.88) | 25 (29.76) | 2308 (32.84) | 2118 (32.96) | 226 (32.10) | 2344 (32.88) |
| Fair | 1343 (19.34) | 16 (19.05) | 1359 (19.34) | 1230 (19.14) | 156 (22.16) | 1386 (19.44) |
| Poor | 598 (8.61) | 16 (19.05) | 614 (8.74) | 545 (8.48) | 82 (11.65) | 627 (8.79) |
| **CES-D** |  |  |  |  |  |  |
| **Mean CES-D (95% CI)** | 2.99 (2.96, 3.02) | 3.10 (2.78, 3.41) | 2.99 (2.96, 3.02) | 2.98 (2.95, 3.01) | 3.14 (3.04, 3.25) | 2.99 (2.96, 3.03) |

Table S30: Health outcomes by sexual identity recoding strategy in adolescents

|  | Heterosexual | SM | Total | Heterosexual | SM | Total |
| --- | --- | --- | --- | --- | --- | --- |
| **Recoding strategy** | **R** | **R** | **R** | **B** | **B** | **B** |
| N= | 8989 | 906 | 9895 | 7888 | 2215 | 10103 |
| **On the whole, I am satisfied with myself** |  |  |  |  |  |  |
| Strongly agree | 1911 (21.26) | 87 (9.60) | 1998 (20.19) | 1787 (22.65) | 233 (10.52) | 2020 (19.99) |
| Agree | 5085 (56.57) | 403 (44.48) | 5488 (55.46) | 4506 (57.12) | 1072 (48.40) | 5578 (55.21) |
| Disagree | 1485 (16.52) | 301 (33.22) | 1786 (18.05) | 1181 (14.97) | 655 (29.57) | 1836 (18.17) |
| Strongly disagree | 493 (5.48) | 114 (12.58) | 607 (6.13) | 401 (5.08) | 242 (10.93) | 643 (6.36) |
| Do Not Know | 7 (0.08) | 1 (0.11) | 8 (0.08) | 5 (0.06) | 9 (0.41) | 14 (0.14) |
| I do not wish to answer | 8 (0.09) | 0 (0.00) | 8 (0.08) | 8 (0.10) | 4 (0.18) | 12 (0.12) |
| **How would you describe your health generally?** |  |  |  |  |  |  |
| Excellent | 2660 (29.59) | 167 (18.43) | 2827 (28.57) | 2395 (30.36) | 471 (21.26) | 2866 (28.37) |
| Very good | 3551 (39.50) | 328 (36.20) | 3879 (39.20) | 3124 (39.60) | 813 (36.70) | 3937 (38.97) |
| Good | 2115 (23.53) | 283 (31.24) | 2398 (24.23) | 1822 (23.10) | 652 (29.44) | 2474 (24.49) |
| Fair | 476 (5.30) | 88 (9.71) | 564 (5.70) | 391 (4.96) | 199 (8.98) | 590 (5.84) |
| Poor | 92 (1.02) | 25 (2.76) | 117 (1.18) | 75 (0.95) | 47 (2.12) | 122 (1.21) |
| No answer | 95 (1.06) | 15 (1.66) | 110 (1.11) | 81 (1.03) | 33 (1.49) | 114 (1.13) |
| **KESSLER** |  |  |  |  |  |  |
| Mean KESSLER (95% CI) | 6.85 (6.75, 6.94) | 10.95 (10.61, 11.29) | 7.22 (7.13, 7.32) | 6.50 (6.40, 6.60) | 10.05 (9.84, 10.27) | 7.27 (7.18, 7.37) |

Table S31: Health outcomes by sexual attraction recoding strategy in adolescents

|  | Heterosexual | SM | Total | Heterosexual | SM | Total |
| --- | --- | --- | --- | --- | --- | --- |
| **Recoding strategy** | **R** | **R** | **R** | **B** | **B** | **B** |
| N= | 9038 | 716 | 9754 | 7656 | 2446 | 10102 |
| **On the whole, I am satisfied with myself** |  |  |  |  |  |  |
| Strongly agree | 1884 (20.85) | 64 (8.94) | 1948 (19.97) | 1734 (22.65) | 286 (11.69) | 2020 (20.00) |
| Agree | 5076 (56.16) | 326 (45.53) | 5402 (55.38) | 4412 (57.63) | 1166 (47.67) | 5578 (55.22) |
| Disagree | 1549 (17.14) | 223 (31.15) | 1772 (18.17) | 1127 (14.72) | 709 (28.99) | 1836 (18.17) |
| Strongly disagree | 516 (5.71) | 102 (14.25) | 618 (6.34) | 373 (4.87) | 269 (11.00) | 642 (6.36) |
| Do Not Know | 7 (0.08) | 1 (0.14) | 8 (0.08) | 5 (0.07) | 9 (0.37) | 14 (0.14) |
| I do not wish to answer | 6 (0.07) | 0 (0.00) | 6 (0.06) | 5 (0.07) | 7 (0.29) | 12 (0.12) |
| **How would you describe your health generally?** |  |  |  |  |  |  |
| Excellent | 2655 (29.38) | 130 (18.16) | 2785 (28.55) | 2350 (30.69) | 516 (21.10) | 2866 (28.37) |
| Very good | 3556 (39.34) | 267 (37.29) | 3823 (39.19) | 3047 (39.80) | 890 (36.39) | 3937 (38.97) |
| Good | 2142 (23.70) | 228 (31.84) | 2370 (24.30) | 1747 (22.82) | 727 (29.72) | 2474 (24.49) |
| Fair | 489 (5.41) | 60 (8.38) | 549 (5.63) | 363 (4.74) | 226 (9.24) | 589 (5.83) |
| Poor | 100 (1.11) | 15 (2.09) | 115 (1.18) | 73 (0.95) | 49 (2.00) | 122 (1.21) |
| No answer | 96 (1.06) | 16 (2.23) | 112 (1.15) | 76 (0.99) | 38 (1.55) | 114 (1.13) |
| **KESSLER** |  |  |  |  |  |  |
| Mean KESSLER (95% CI) | 6.98 (6.88, 7.08) | 11.07 (10.68, 11.46) | 7.28 (7.18, 7.38) | 6.43 (6.33, 6.53) | 9.93 (9.72, 10.13) | 7.27 (7.18, 7.37) |

Table S32: Health outcomes by combined dimension recoding strategy in adolescents

|  | Heterosexual | SM | Total | Heterosexual | SM | Total |
| --- | --- | --- | --- | --- | --- | --- |
| **Recoding strategy** | **R** | **R** | **R** | **B** | **B** | **B** |
| N= | 9291 | 480 | 9771 | 7335 | 2746 | 10081 |
| **On the whole, I am satisfied with myself** |  |  |  |  |  |  |
| Strongly agree | 1942 (20.90) | 30 (6.25) | 1972 (20.18) | 1692 (23.07) | 321 (11.69) | 2013 (19.97) |
| Agree | 5217 (56.15) | 230 (47.92) | 5447 (55.75) | 4223 (57.57) | 1345 (48.98) | 5568 (55.23) |
| Disagree | 1587 (17.08) | 155 (32.29) | 1742 (17.83) | 1060 (14.45) | 773 (28.15) | 1833 (18.18) |
| Strongly disagree | 528 (5.68) | 65 (13.54) | 593 (6.07) | 351 (4.79) | 292 (10.63) | 643 (6.38) |
| Do Not Know | 9 (0.10) | 0 (0.00) | 9 (0.09) | 4 (0.05) | 9 (0.33) | 13 (0.13) |
| I do not wish to answer | 8 (0.09) | 0 (0.00) | 8 (0.08) | 5 (0.07) | 6 (0.22) | 11 (0.11) |
| **How would you describe your health generally?** |  |  |  |  |  |  |
| Excellent | 2721 (29.29) | 83 (17.29) | 2804 (28.70) | 2265 (30.88) | 597 (21.74) | 2862 (28.39) |
| Very good | 3652 (39.31) | 183 (38.13) | 3835 (39.25) | 2926 (39.89) | 1003 (36.53) | 3929 (38.97) |
| Good | 2206 (23.74) | 161 (33.54) | 2367 (24.22) | 1663 (22.67) | 804 (29.28) | 2467 (24.47) |
| Fair | 510 (5.49) | 36 (7.50) | 546 (5.59) | 341 (4.65) | 247 (8.99) | 588 (5.83) |
| Poor | 105 (1.13) | 10 (2.08) | 115 (1.18) | 65 (0.89) | 56 (2.04) | 121 (1.20) |
| No answer | 97 (1.04) | 7 (1.46) | 104 (1.06) | 75 (1.02) | 39 (1.42) | 114 (1.13) |
| **KESSLER** |  |  |  |  |  |  |
| Mean KESSLER (95% CI) | 6.97 (6.87, 7.07) | 11.32 (10.88, 11.76) | 7.18 (7.09, 7.28) | 6.35 (6.25, 6.46) | 9.76 (9.56, 9.95) | 7.28 (7.18, 7.37) |
